# Effect of Ventilator Mode on Ventilator-Free Days in Critically Ill Adults: A Randomized Trial

**DOI:** 10.1101/2024.10.08.24314961

**Authors:** Kevin P. Seitz, Bradley D. Lloyd, Li Wang, Matthew S. Shotwell, Edward T. Qian, Amelia L. Muhs, Roger K. Richardson, J. Craig Rooks, Vanessa Hennings-Williams, Claire E. Sandoval, Whitney D. Richardson, Tracy L. Morgan, Amber N. Thompson, Pamela G. Hastings, Terry P. Ring, Joanna L. Stollings, Erica M. Talbot, David J. Krasinski, Bailey R. DeCoursey, Tanya K. Marvi, Stephanie C. DeMasi, Kevin W. Gibbs, Wesley H. Self, Amanda S. Mixon, Todd W. Rice, Matthew W. Semler, Jonathan D. Casey, the Pragmatic Critical Care Research Group

**Affiliations:** Vanderbilt University Medical Center, Department of Medicine, Division of Allergy, Pulmonary and Critical Care Medicine, Nashville, TN; Vanderbilt University Medical Center, Department of Emergency Medicine, Nashville, TN; Vanderbilt University Medical Center, Department of Biostatistics, Nashville, TN; Vanderbilt University Medical Center, Department of Anesthesiology, Nashville, TN; Vanderbilt University Medical Center, Department of Respiratory Care, Nashville, TN; Vanderbilt University Medical Center, Department of Pharmaceutical Services, Nashville, TN; Vanderbilt University Medical Center, Department of Medicine, Nashville, TN; University of Colorado School of Medicine, Department of Medicine, Division of Pulmonary Sciences and Critical Care Medicine, Aurora, CO; Wake Forest School of Medicine, Department of Medicine, Section of Pulmonary, Critical Care, Allergy, and Immunologic Disease, Winston-Salem, NC; Vanderbilt University Medical Center, Vanderbilt Institute for Clinical and Translational Research, Nashville, TN; Vanderbilt University Medical Center, Department of Medicine, Division of General Internal Medicine and Public Health, Nashville, TN; VA Tennessee Valley Healthcare System, Geriatric Research, Education, and Clinical Center, Nashville, TN

**Keywords:** Critical Illness, Respiratory Failure, Respiration, Artificial, Clinical Trial

## Abstract

**Rationale:** For critically ill adults receiving invasive mechanical ventilation, the ventilator mode determines how breaths are delivered. Whether the choice of ventilator mode affects outcomes for critically ill patients is unknown. To compare the effects of three common ventilator modes (volume control, pressure control, and adaptive pressure control) on death and duration of mechanical ventilation.

**Methods:** We conducted a pragmatic, cluster-randomized, crossover trial among adults receiving invasive mechanical ventilation in a medical ICU between November 1, 2022 and July 31, 2023. Each month, patients in the participating unit were assigned to receive volume control, pressure control, or adaptive pressure control during continuous mandatory ventilation. The primary outcome was ventilator-free days through 28 days.

**Results:** Among 566 patients included in the primary analysis, the median number of ventilator-free days was 23 [IQR, 0-26] in the volume control group, 22 [0-26] in the pressure control group, and 24 [0-26] in the adaptive pressure control group (P=0.60). The median tidal volume was similar in the three groups, but the percentage of breaths larger than 8mL/kg of predicted body weight differed between volume control (median, 4.0%; IQR, 0.0-14.1), pressure control (10.6%; 0.0-31.5), and adaptive pressure control (4.7%; 0.0-19.2). Incidences of hypoxemia, acidemia, and barotrauma were similar in the three groups.

**Conclusions:** Among critically ill adults receiving invasive mechanical ventilation, the use of volume control, pressure control, or adaptive pressure control did not affect the number of ventilator-free days, however, confidence intervals included differences that may be clinically meaningful.

## INTRODUCTION

Millions of critically ill adults receive invasive mechanical ventilation each year.^1,2^ While potentially life-saving, mechanical ventilation itself may injure the lungs, diaphragm, and other organs.^3–5^ In-hospital mortality among critically ill adults receiving mechanical ventilation in the United States remains greater than 30%.^6–8^

To provide invasive mechanical ventilation, clinicians must select a ventilator mode, which determines how the ventilator delivers each breath.^9,10^ The ventilator can administer a set volume and measure the change in pressure that occurs with the breath (e.g., “volume control” mode), administer a set pressure and measure the change in volume that occurs with the breath (e.g., “pressure control” mode), or use an algorithm to titrate the pressure administered breath-by-breath to maintain a target volume set by the clinician (e.g., “adaptive pressure control” mode). While previous studies have shown that avoiding large volumes, high pressures, and deep sedation may all improve patient outcomes,^11–14^ each ventilator mode delivers breaths in a way that prioritizes these variables differently. Volume control can facilitate tighter control of the volume administered but lead to higher pressures.^15,16^ Pressure control can facilitate tighter control of the pressure administered but lead to higher volumes.^17^ As a dual-control mode, adaptive pressure control could provide the benefits of both volume control and pressure control.^18,19^

Although volume control, pressure control, and adaptive pressure control are the three most commonly used ventilator modes in current clinical care,^20–23^ they have never been directly compared in a randomized trial, and whether the choice of ventilator modes affects outcomes for critically ill adults is unknown. To address this gap, the Mode of Ventilation During Critical Illness (MODE) trial compared the use of volume control, pressure control, and adaptive pressure control among critically ill adults receiving mechanical ventilation.

## METHODS

### Trial Design and Oversight

Between November 1, 2022 and July 31, 2023, we conducted a pragmatic, unblinded, cluster-randomized, cluster-crossover trial comparing the use of volume control, pressure control, and adaptive pressure control ventilator modes among critically ill adults receiving invasive mechanical ventilation in an academic medical intensive care unit (ICU). The trial was initiated by the investigators, approved by the local institutional review board with a waiver of informed consent, registered before enrollment commenced (NCT05563779), and overseen by an independent data and safety monitoring board (additional details appear in the Supplemental Methods of the online data supplement). The trial protocol and statistical analysis plan were published before enrollment concluded.^24,25^

### Patient Population

The trial was conducted in the medical ICU at Vanderbilt University Medical Center in Nashville, Tennessee. All adults (≥18 years of age) located in the study unit were enrolled at the time of the first receipt of invasive mechanical ventilation in the unit. Patients were excluded if they were pregnant, incarcerated, receiving extracorporeal membrane oxygenation, or had received invasive mechanical ventilation at their place of residence prior to hospital admission.

### Randomization and Treatment Allocation

All eligible patients in the study ICU were assigned together as a cluster to a single ventilator mode (cluster-level randomization). Each month, the ICU switched between the use of volume control, pressure control, or adaptive pressure control in a randomly generated sequence (cluster-level crossover). The order of study group assignments for the 9 months of the trial was generated using randomization with permuted blocks of three to minimize the impact of seasonal variation and temporal changes. Patients were analyzed in the group to which they were assigned at enrollment even if they remained in the study ICU during a transition from one month to the next. To reduce the number of patients who experienced a crossover of the study unit from one assigned mode to another, the last 3 days of each month were considered an analytic washout period during which the study ICU continued to use the assigned ventilator mode, but new patients were not included in the primary analysis. Patients, clinicians, and investigators were not blinded to group assignment.

### Study Interventions

In each of the three trial groups (volume control group, pressure control group, and adaptive pressure control group), trial protocol specified that patients should receive the assigned ventilator mode during continuous mandatory ventilation. The trial protocol instructed respiratory therapists to use the assigned mode beginning at the first receipt of invasive mechanical ventilation in the study ICU and ending at extubation from mechanical ventilation, transfer out of the study ICU, or the end of the one-month study block, whichever occurred first. The trial protocol did not determine the ventilator mode during time-periods in which the patient was not receiving a continuous mandatory mode of ventilation (e.g., while receiving pressure support ventilation during a spontaneous breathing trial), was not physically located in the study ICU (e.g., during transport), or was undergoing an invasive procedure (e.g., bronchoscopy). For patients who continued to receive mechanical ventilation in a trial location after the end of a study block, the ventilator mode was selected by treating clinicians. If, at any time, a treating clinician determined that a ventilator mode other than that assigned by the trial might be best for the treatment of the patient, the ventilator mode for that patient was modified and the reason for modifying the ventilator mode was recorded.

The trial protocol controlled only the ventilator mode. Regardless of group assignment, existing ventilator protocols in the study ICU recommended the use of lung protective ventilation with a target tidal volume of 6 mL/kg of predicted body weight (PBW) and a plateau pressure less than 30 cm H_2_O. Other aspects of mechanical ventilation, including the set respiratory rate, positive end-expiratory pressure, gas flow rate, inspiratory and expiratory time, timing of extubation, and administration of analgesia and sedation were determined by institutional protocols and treating clinicians (see Supplemental Methods in the online data supplement).

### Data Collection

Trial personnel collected data from the electronic health record on baseline characteristics, on-study management, and clinical outcomes using a standardized case-report form. Data on ventilator mode and oxygen saturation as measured by pulse oximetry (SpO_2_) were automatically extracted from the bedside monitor at a frequency of every 1 minute with the use of a previously validated approach.^26,27^ Values for tidal volume and peak inspiratory pressure were automatically extracted every hour (additional details are in the online data supplement).

### Study Outcomes

The primary outcome was the number of days alive and free of invasive mechanical ventilation (ventilator-free days) through day 28 after enrollment, defined as the number of calendar days alive and free of invasive mechanical ventilation from the final receipt of invasive mechanical ventilation through day 28 after enrollment.^28,29^ Outcome ascertainment ceased at hospital discharge or after day 28, whichever occurred first. Patients were assigned a value of 0 ventilator-free days if they died on or before day 28, continued to receive mechanical ventilation on day 28, or continued to receive mechanical ventilation at the time of hospital discharge prior to day 28. Additional outcomes are described in the Supplemental Methods in the online data supplement.

### Statistical Analysis

Details of the sample-size calculation have been reported previously.^25^ Using data from a previous trial conducted in the same setting,^27^ we estimated that, during the 9 months of the pilot trial, 606 patients would be enrolled and included in the primary analysis. With a median number of ventilator-free days of 22 (interquartile range [IQR], 0 to 25), and an intracluster intraperiod correlation of 0.01, we calculated that enrollment of 606 patients would provide 80% power at a two-sided alpha level of 0.05 to detect an absolute difference between groups of 3 ventilator-free days, comparable to the difference considered to be clinically meaningful in the design of prior trials.^30^

The primary analysis population included all enrolled patients except those enrolled during the 3-day washout periods at the end of each month. In the primary analysis, the number of ventilator-free days was compared between patients assigned to the volume control, pressure control, and adaptive pressure control groups with the use of a proportional-odds model with independent variables of group assignment and time. To account for seasonality and secular trends, time from the start of the trial (in days) was included as a continuous variable with values ranging from 1 (first day of enrollment) to 272 (final day of enrollment). The variable of time was modeled using restricted cubic splines with five knots to allow for non-linearity. In addition to assessing for an overall group effect within the model, we estimated the differences between each pair of ventilator modes by extracting 95% confidence intervals from the model.

In sensitivity analyses, we used alternative definitions of the trial population and alternative statistical methods for analyzing the primary outcome, including an analysis adjusting for baseline covariates and an analysis among all patients enrolled in the trial including those enrolled during the washout periods. Effect modification was assessed in a proportional odds model by including an interaction term between trial-group assignment and a prespecified baseline variable. Prespecified potential effect modifiers included age, chronic obstructive pulmonary disease, duration of invasive mechanical ventilation prior to enrollment, pre-enrollment fraction of inspired oxygen, Sequential Organ Failure Assessment (SOFA) score at enrollment,^31^ presence of shock requiring vasopressors at enrollment, and the indication for mechanical ventilation (hypoxemic respiratory failure, hypercarbic respiratory failure, and altered mental status).

The data and safety monitoring board reviewed a single, planned interim analysis for safety after the first 3 months of enrollment (see Supplemental Methods in the online data supplement). There was no interim analysis for efficacy. For the final analysis of the primary outcome, a two-sided P value of less than 0.05 was considered to indicate statistical significance. All analyses other than the primary analysis of the primary outcome were considered hypothesis-generating, and no statistical corrections were made for multiple comparisons. All analyses were performed using R 4.3.0 (R Foundation for Statistical Computing, Vienna, Austria). Some of the results of this trial have been previously reported in the form of an abstract.^32^

## RESULTS

### Trial Population

Of the 656 patients who received invasive mechanical ventilation in the study ICU during the study period, 37 (5.6%) met exclusion criteria and 619 (94.4%) were enrolled in the trial (Figure 1 and E1 in the online data supplement). Of those enrolled, 53 (8.6%) were enrolled during an analytic washout period, and the remaining 566 (91.4%) were included in the primary analysis. Among the 566 patients in the primary analysis, 181 (32.0%) were assigned to the volume control group, 198 (35.0%) to the pressure control group, and 187 (33.0%) to the adaptive pressure control group. The trial groups had similar baseline characteristics (Table 1). The median age was 58 years, 41.3% were female, 39.8% had hypoxemic respiratory failure as an indication for mechanical ventilation, and 210 (37.1%) had acute respiratory distress syndrome. Complete information on baseline patient characteristics is available in Tables E1 to E6 in the online data supplement.

**Figure 1.**
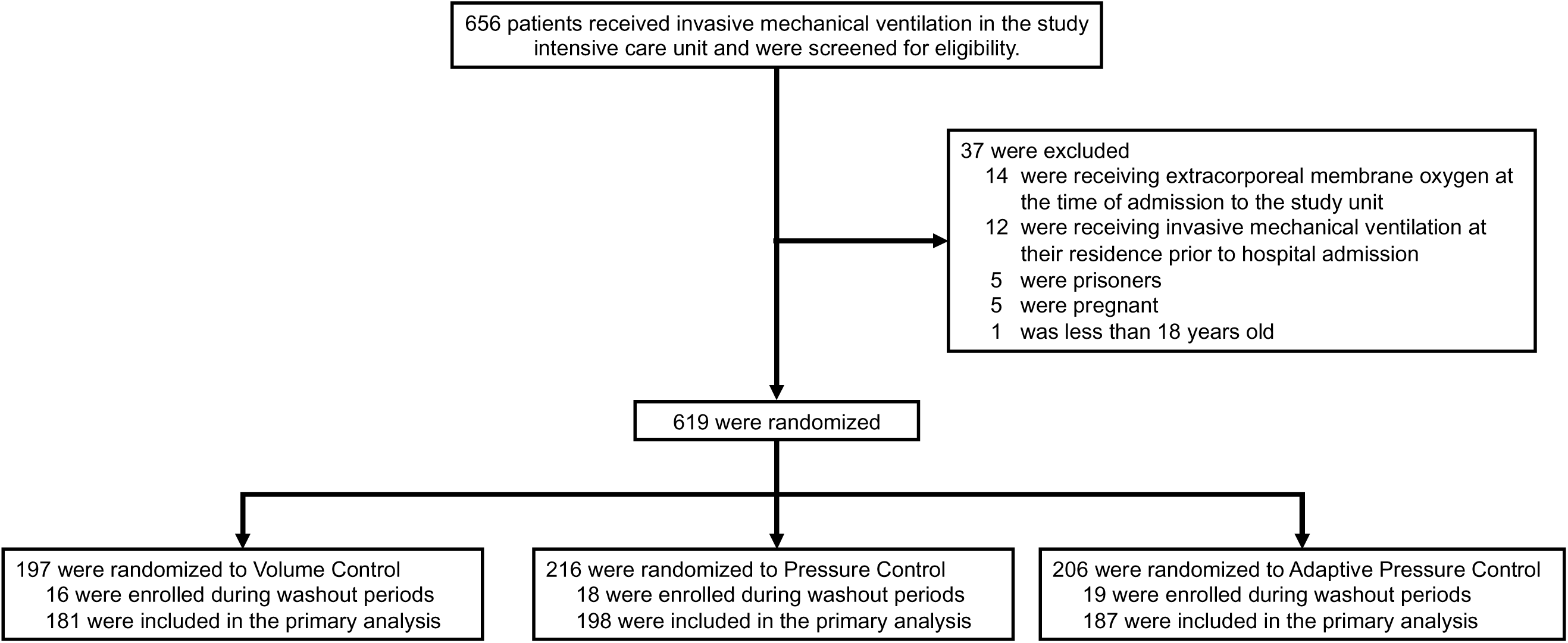
Flow of participants through the trial.

**Table 1.**
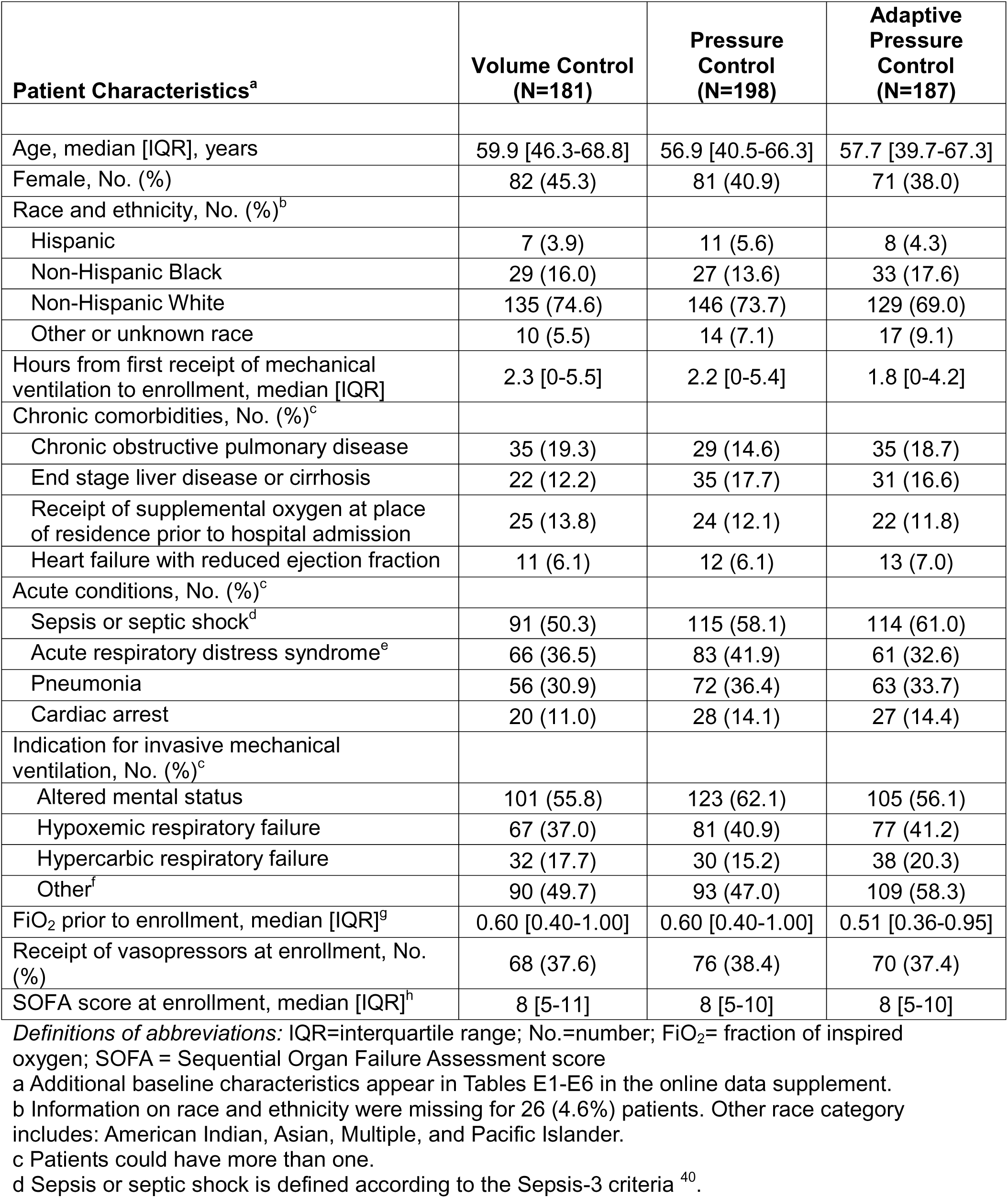
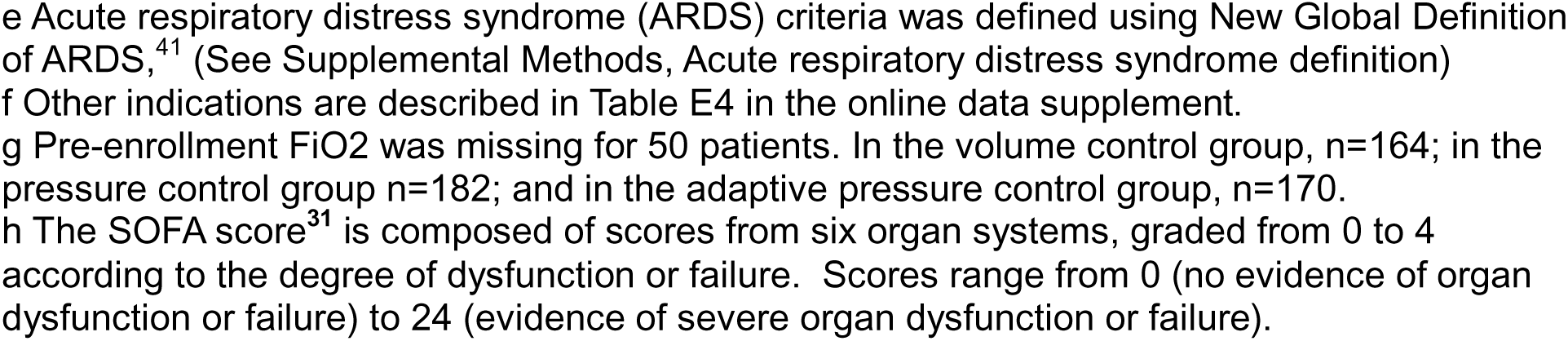
Patient Characteristics at Baseline.

| **Patient Characteristics<sup>a</sup>** | **Volume Control**<br>**(N=181)** | **Pressure**<br>**Control**<br>**(N=198)** | **Adaptive**<br>**Pressure**<br>**Control**<br>**(N=187)** |
| --- | --- | --- | --- |
| Age, median [IQR], years | 59.9 [46.3-68.8] | 56.9 [40.5-66.3] | 57.7 [39.7-67.3] |
| Female, No. (%) | 82 (45.3) | 81 (40.9) | 71 (38.0) |
| Race and ethnicity, No. (%)<sup>b</sup> |  |  |  |
| Hispanic | 7 (3.9) | 11 (5.6) | 8 (4.3) |
| Non-Hispanic Black | 29 (16.0) | 27 (13.6) | 33 (17.6) |
| Non-Hispanic White | 135 (74.6) | 146 (73.7) | 129 (69.0) |
| Other or unknown race | 10 (5.5) | 14 (7.1) | 17 (9.1) |
| Hours from first receipt of mechanical ventilation to enrollment, median [IQR] | 2.3 [0-5.5] | 2.2 [0-5.4] | 1.8 [0-4.2] |
| Chronic comorbidities, No. (%)<sup>c</sup> |  |  |  |
| Chronic obstructive pulmonary disease | 35 (19.3) | 29 (14.6) | 35 (18.7) |
| End stage liver disease or cirrhosis | 22 (12.2) | 35 (17.7) | 31 (16.6) |
| Receipt of supplemental oxygen at place of residence prior to hospital admission | 25 (13.8) | 24 (12.1) | 22 (11.8) |
| Heart failure with reduced ejection fraction | 11 (6.1) | 12 (6.1) | 13 (7.0) |
| Acute conditions, No. (%)<sup>c</sup> |  |  |  |
| Sepsis or septic shock<sup>d</sup> | 91 (50.3) | 115 (58.1) | 114 (61.0) |
| Acute respiratory distress syndrome<sup>e</sup> | 66 (36.5) | 83 (41.9) | 61 (32.6) |
| Pneumonia | 56 (30.9) | 72 (36.4) | 63 (33.7) |
| Cardiac arrest | 20 (11.0) | 28 (14.1) | 27 (14.4) |
| Indication for invasive mechanical ventilation, No. (%)<sup>c</sup> |  |  |  |
| Altered mental status | 101 (55.8) | 123 (62.1) | 105 (56.1) |
| Hypoxemic respiratory failure | 67 (37.0) | 81 (40.9) | 77 (41.2) |
| Hypercarbic respiratory failure | 32 (17.7) | 30 (15.2) | 38 (20.3) |
| Other<sup>f</sup> | 90 (49.7) | 93 (47.0) | 109 (58.3) |
| FiO<sub>2</sub> prior to enrollment, median [IQR]<sup>g</sup> | 0.60 [0.40-1.00] | 0.60 [0.40-1.00] | 0.51 [0.36-0.95] |
| Receipt of vasopressors at enrollment, No. (%) | 68 (37.6) | 76 (38.4) | 70 (37.4) |
| SOFA score at enrollment, median [IQR]<sup>h</sup> | 8 [5-11] | 8 [5-10] | 8 [5-10] |

*Definitions of abbreviations:* IQR=interquartile range; No.=number; FiO<sub>2</sub>= fraction of inspired oxygen; SOFA = Sequential Organ Failure Assessment score
a Additional baseline characteristics appear in Tables E1-E6 in the online data supplement.
b Information on race and ethnicity were missing for 26 (4.6%) patients. Other race category includes: American Indian, Asian, Multiple, and Pacific Islander.
c Patients could have more than one.
d Sepsis or septic shock is defined according to the Sepsis-3 criteria <sup>40</sup>.
e Acute respiratory distress syndrome (ARDS) criteria was defined using New Global Definition of ARDS,<sup>41</sup> (See Supplemental Methods, Acute respiratory distress syndrome definition)
f Other indications are described in Table E4 in the online data supplement.
g Pre-enrollment FiO<sub>2</sub> was missing for 50 patients. In the volume control group, n=164; in the pressure control group n=182; and in the adaptive pressure control group, n=170.
h The SOFA score<sup>31</sup> is composed of scores from six organ systems, graded from 0 to 4 according to the degree of dysfunction or failure. Scores range from 0 (no evidence of organ dysfunction or failure) to 24 (evidence of severe organ dysfunction or failure).

### Invasive Mechanical Ventilation and ICU Interventions

Which ventilator mode the patient was receiving was recorded a median of every 1 minute (interquartile range [IQR], 1 to 1) between enrollment and cessation of invasive mechanical ventilation, resulting in 1,405,218 assessments of ventilator mode among the 566 patients (Supplemental Methods in the online data supplement). In the 72 hours after enrollment, when patients were receiving continuous mandatory ventilation, the median proportion of ventilator mode assessments that were in the assigned mode was 100% (IQR, 100% to 100%) in the volume control group, 100% (IQR, 98.9% to 100%) in the pressure control group, and 100% (IQR, 100% to 100%) in the adaptive pressure control group (Figure 2, and Figures E2 to E4 and Tables E7 to E10 in the online data supplement). A total of 61 (10.8%) patients in the primary analysis experienced a crossover, after spending a median of 8 days (IQR, 5 to 14) in their assigned group, and had their ventilator mode selected by treating clinicians for the remainder of their time on mechanical ventilation (Table E9).

**Figure 2.**
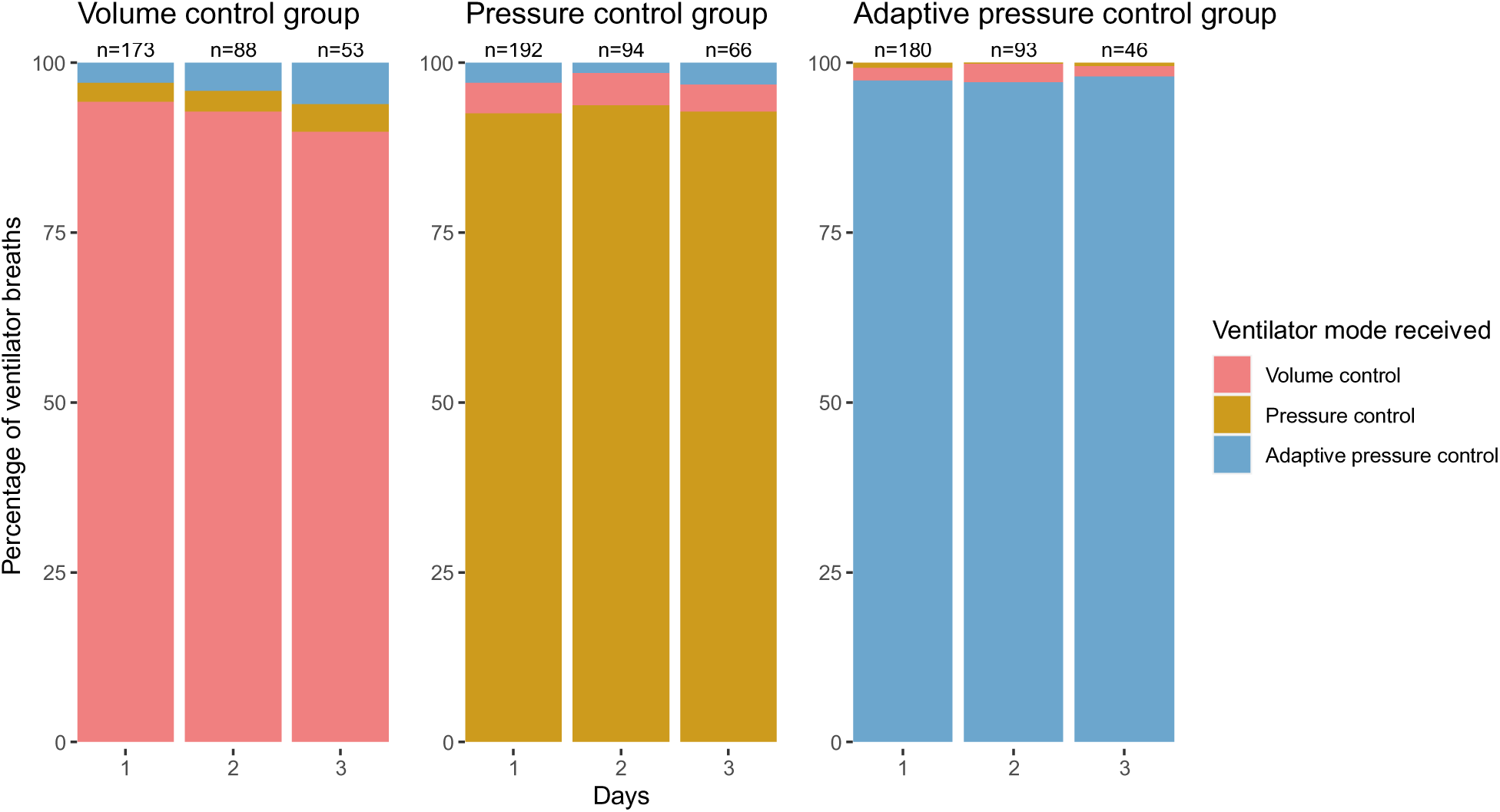
Ventilator mode received by each group. Shown are the percentages of ventilator breaths in each trial group that were in volume control mode (red), pressure control mode (yellow), and adaptive pressure control mode (blue) for the 72 hours following enrollment. Ventilator mode was assessed approximately every 1 minute. This figure displays data on breaths for which the patient was receiving a continuous mandatory mode of ventilation. Also shown is the number of patients who were alive and receiving continuous mandatory ventilation in each group during each time interval. The percentage of breaths in a continuous mandatory mode and in a spontaneous mode (e.g., pressure support) were 86.4% and 13.6% on day 1, 78.7% and 21.3% on day 2, and 75.7% and 24.3% on day 3. Ventilator mode is reported for 545 of 566 patients on study day 1. A total of 18 patients did not receive a continuous mandatory mode (i.e., they received only spontaneous modes) on day 1, of whom, 5 received a continuous mandatory mode on study days 2 or 3. Three patients were missing data on the ventilator modes received. Additional data on ventilator modes are in Figures E2-E4 and Table E7 in the online data supplement.

The mean exhaled tidal volume between enrollment and cessation of mechanical ventilation for each patient (in mL/kg of predicted body weight) did not differ between groups: median of 6.4 (IQR, 5.9 to 7.1) in the volume control group; 6.5 (IQR, 5.7 to 7.4) in the pressure control group, and 6.3 (IQR, 6.0 to 7.0) in the adaptive pressure control group (P=0.23) (Table 2, Figure E5 and Table E11 in the online supplement). The percentage of each patient’s tidal volumes that were greater than 8 mL/kg of predicted body weight differed significantly between groups: median of 4.0 percent (IQR, 0.0 to 14.1) in the volume control group, 10.6 percent (IQR, 0.0 to 31.5) in the pressure control group, and 4.7 percent (IQR, 0.0 to 19.2) in the adaptive pressure control group (P<0.001) (Table 2, Figure E6 and Table E11 in the online data supplement). The mean peak inspiratory pressure for each patient differed significantly between groups: median of 21.6 cmH_2_O (IQR, 18.1 to 27.0) in the volume control group, 19.3 cmH_2_O (IQR, 15.7 to 25.1) in the pressure control group, and 20.2 cmH_2_O (IQR, 16.1 to 24.2) in the adaptive pressure control group (P=0.006) (Table 2, Figure E7 and Table E11 in the online data supplement). The depth of sedation on each study day differed significantly between the groups: median Richmond Agitation and Sedation Scale score of −1.0 (IQR, −2.2 to −0.5) in the volume control group, −0.9 (IQR, −2.7 to −0.5) in the pressure control group, and −0.8 (IQR, −2.3 to −0.4) in the adaptive pressure control group (P=0.04) (Figure E8 and Table E12 in the online data supplement). The receipt of analgesic or sedating medications did not differ between groups on any study day (Table E12 in the online data supplement). Information on FiO_2_, positive end expiratory pressures, coma, delirium, sedative receipt, and blood gas measurements are provided in Table 2, Figure E9 and E10, and Tables E12 to E14 in the online data supplement.

**Table 2.**
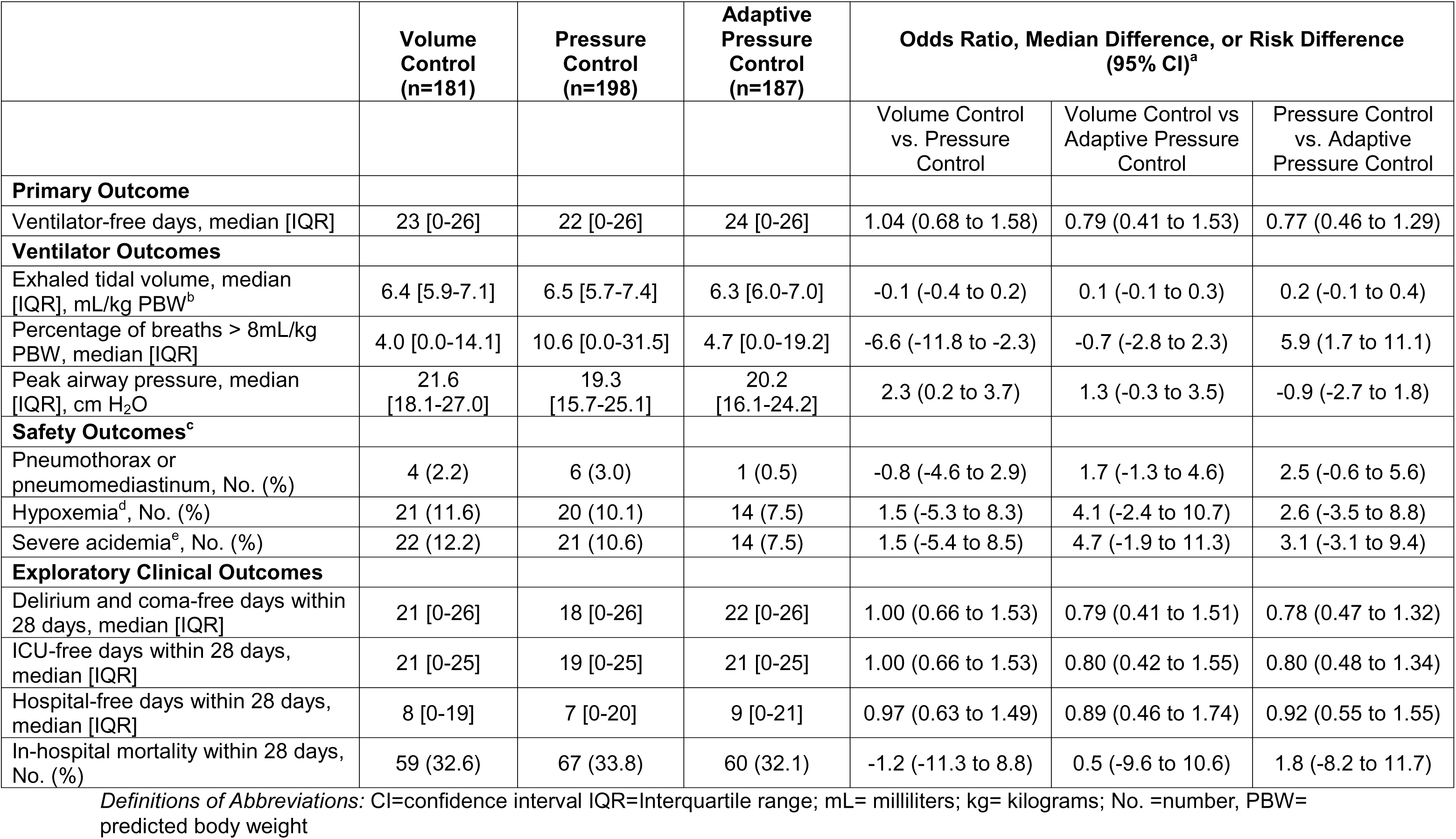
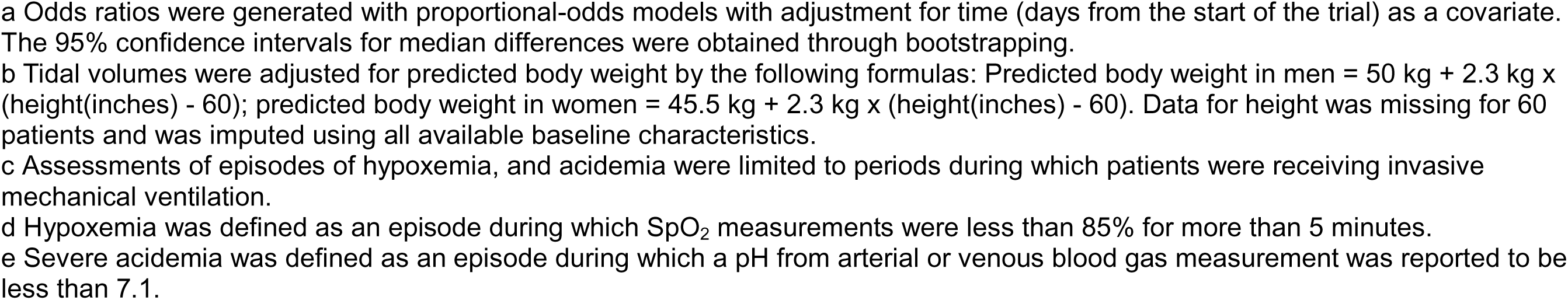
Outcomes.

|  | Volume Control<br>(n=181) | Pressure Control<br>(n=198) | Adaptive Pressure Control<br>(n=187) | Odds Ratio, Median Difference, or Risk Difference<br>(95% CI)<sup>a</sup> |  |  |
| --- | --- | --- | --- | --- | --- | --- |
|  |  |  |  | Volume Control vs. Pressure Control | Volume Control vs Adaptive Pressure Control | Pressure Control vs. Adaptive Pressure Control |
| **Primary Outcome** |  |  |  |  |  |  |
| Ventilator-free days, median [IQR] | 23 [0-26] | 22 [0-26] | 24 [0-26] | 1.04 (0.68 to 1.58) | 0.79 (0.41 to 1.53) | 0.77 (0.46 to 1.29) |
| **Ventilator Outcomes** |  |  |  |  |  |  |
| Exhaled tidal volume, median [IQR], mL/kg PBW<sup>b</sup> | 6.4 [5.9-7.1] | 6.5 [5.7-7.4] | 6.3 [6.0-7.0] | -0.1 (-0.4 to 0.2) | 0.1 (-0.1 to 0.3) | 0.2 (-0.1 to 0.4) |
| Percentage of breaths > 8mL/kg PBW, median [IQR] | 4.0 [0.0-14.1] | 10.6 [0.0-31.5] | 4.7 [0.0-19.2] | -6.6 (-11.8 to -2.3) | -0.7 (-2.8 to 2.3) | 5.9 (1.7 to 11.1) |
| Peak airway pressure, median [IQR], cm H<sub>2</sub>O | 21.6 [18.1-27.0] | 19.3 [15.7-25.1] | 20.2 [16.1-24.2] | 2.3 (0.2 to 3.7) | 1.3 (-0.3 to 3.5) | -0.9 (-2.7 to 1.8) |
| **Safety Outcomes<sup>c</sup>** |  |  |  |  |  |  |
| Pneumothorax or pneumomediastinum, No. (%) | 4 (2.2) | 6 (3.0) | 1 (0.5) | -0.8 (-4.6 to 2.9) | 1.7 (-1.3 to 4.6) | 2.5 (-0.6 to 5.6) |
| Hypoxemia<sup>d</sup>, No. (%) | 21 (11.6) | 20 (10.1) | 14 (7.5) | 1.5 (-5.3 to 8.3) | 4.1 (-2.4 to 10.7) | 2.6 (-3.5 to 8.8) |
| Severe acidemia<sup>e</sup>, No. (%) | 22 (12.2) | 21 (10.6) | 14 (7.5) | 1.5 (-5.4 to 8.5) | 4.7 (-1.9 to 11.3) | 3.1 (-3.1 to 9.4) |
| **Exploratory Clinical Outcomes** |  |  |  |  |  |  |
| Delirium and coma-free days within 28 days, median [IQR] | 21 [0-26] | 18 [0-26] | 22 [0-26] | 1.00 (0.66 to 1.53) | 0.79 (0.41 to 1.51) | 0.78 (0.47 to 1.32) |
| ICU-free days within 28 days, median [IQR] | 21 [0-25] | 19 [0-25] | 21 [0-25] | 1.00 (0.66 to 1.53) | 0.80 (0.42 to 1.55) | 0.80 (0.48 to 1.34) |
| Hospital-free days within 28 days, median [IQR] | 8 [0-19] | 7 [0-20] | 9 [0-21] | 0.97 (0.63 to 1.49) | 0.89 (0.46 to 1.74) | 0.92 (0.55 to 1.55) |
| In-hospital mortality within 28 days, No. (%) | 59 (32.6) | 67 (33.8) | 60 (32.1) | -1.2 (-11.3 to 8.8) | 0.5 (-9.6 to 10.6) | 1.8 (-8.2 to 11.7) |

*Definitions of Abbreviations:* CI=confidence interval IQR=Interquartile range; mL= milliliters; kg= kilograms; No. =number, PBW=predicted body weight
- a Odds ratios were generated with proportional-odds models with adjustment for time (days from the start of the trial) as a covariate. The 95% confidence intervals for median differences were obtained through bootstrapping. - b Tidal volumes were adjusted for predicted body weight by the following formulas: Predicted body weight in men = 50 kg + 2.3 kg x (height(inches) - 60); predicted body weight in women = 45.5 kg + 2.3 kg x (height(inches) - 60). Data for height was missing for 60 patients and was imputed using all available baseline characteristics. - c Assessments of episodes of hypoxemia, and acidemia were limited to periods during which patients were receiving invasive mechanical ventilation. - d Hypoxemia was defined as an episode during which SpO<sub>2</sub> measurements were less than 85% for more than 5 minutes. - e Severe acidemia was defined as an episode during which a pH from arterial or venous blood gas measurement was reported to be less than 7.1.

### Primary Outcome

The number of ventilator-free days through day 28 did not differ significantly between the three study groups, with a median of 23 days (IQR, 0 to 26) in the volume control group, 22 days (IQR, 0 to 26) in the pressure control group, and 24 days (IQR, 0 to 26) in the adaptive pressure control group (P=0.60) (Figure 3, Table 2, and Table E15 and E16). Results were similar in analyses that adjusted for baseline covariates and analyses that included patients enrolled during analytic washout periods (Tables E17 and E18 in the online data supplement). None of the prespecified baseline variables modified the effect of trial group assignment on the primary outcome except hypercarbic respiratory failure as an indication for mechanical ventilation, with fewer ventilator-free days in the volume control and pressure control groups than in the adaptive pressure control group (odds ratio for volume control vs adaptive pressure control, 0.30; 95% 0.11 to 0.83 and odds ratio for pressure control vs adaptive pressure control, 0.32; 95% CI, 0.13 to 0.83). In a post hoc analysis, the presence of acute respiratory distress syndrome criteria at enrollment did not modify the effect of trial group assignment on the primary outcome (Figures E11 to E14 in the online data supplement). (Figures E11 to E14 in the online data supplement).

**Figure 3.**
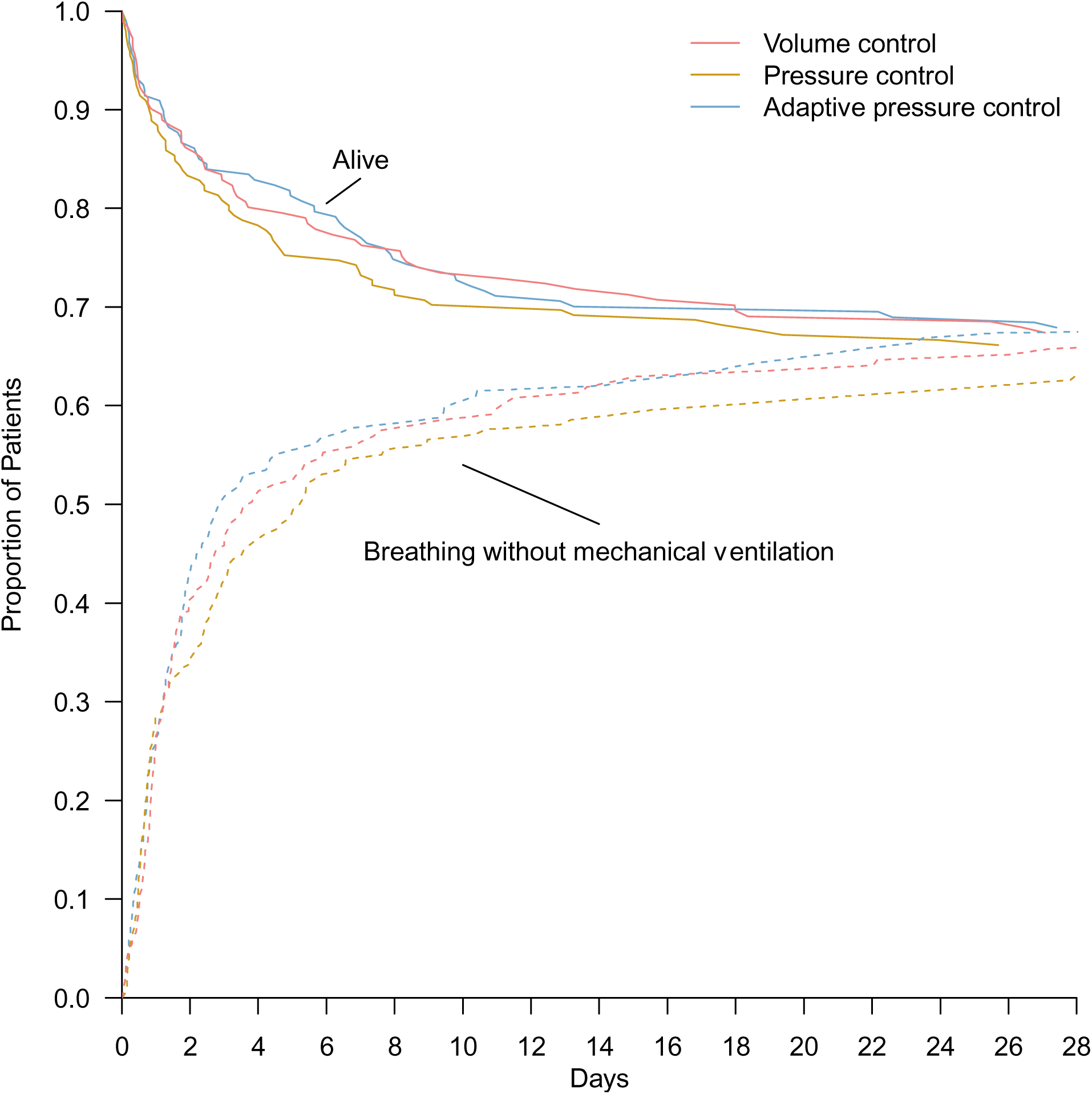
Proportion of patients alive and not receiving invasive mechanical ventilation. The proportions of patients who were alive (solid lines) and breathing without invasive mechanical ventilation (dotted lines) during the 28 days after enrollment in each ventilator mode group are shown. In a proportional-odds model, the number of days that patients were alive and free of invasive mechanical ventilation through day 28 did not differ significantly among the three study groups (P=0.60).

### Exploratory Outcomes

At 28 days, 59 patients (32.6%) in the volume control group, 67 patients (33.8%) in the pressure control group, and 60 patients (32.1%) in the adaptive pressure control group had died before hospital discharge (Table 2). Other pre-specified exploratory outcomes, including daily SOFA score, delirium and coma-free days, ICU-free days, and hospital-free days were similar between the three groups (Table 2, Figure E15 and Tables E11 and E12 in the online data supplement). The safety outcome of pneumothorax or pneumomediastinum occurred in 4 patients (2.2%) in the volume control group, 6 patients (3.0%) in the pressure control group, and 1 (0.5%) patient in the adaptive pressure control group. The safety outcomes of hypoxemia and of severe acidemia were similar between groups (Table 2).

## DISCUSSION

Among critically ill adults receiving invasive mechanical ventilation in this clinical trial, the number of ventilator-free days did not differ between patients treated with volume control, pressure control, or adaptive pressure control. Measures of protective mechanical ventilation differed between groups, with patients in the volume control group experiencing higher peak inspiratory pressures, patients in the pressure control group experiencing a higher percentage of breaths with a tidal volume greater than 8 mL/kg of predicted body weight, and patients in the adaptive pressure control group experiencing less deep sedation.

For each of the millions of adults who receive invasive mechanical ventilation each year, clinicians must select a ventilator mode. Only two previous randomized trials have compared the effects of commonly-used mandatory ventilator modes on outcomes for critically ill adults.^33,34^ These trials enrolled a total of 106 patients combined, compared only volume control and pressure control, and were completed before lung-protective ventilation and daily interruption of sedation were established as evidence-based practices.^11,12^ No prior randomized trial has examined the effect of adaptive pressure control on clinical outcomes. Among almost 600 patients in the current trial, the assigned ventilator mode affected the tidal volumes and inspiratory pressures that patients experienced but did not result in a statistically significant difference in the number of ventilator-free days. These findings provide clinicians with reassurance that the choice between ventilator modes does not result in a large overall difference in death or the duration of mechanical ventilation. However, because the numerical differences in ventilator-free days between the adaptive pressure control (24 days), volume control (23 days), and pressure control (22 days) groups and the associated confidence intervals include differences that could be clinically meaningful, the effects of ventilator mode on patient outcomes warrant further evaluation in randomized trials.

Each of the three ventilator modes evaluated in this trial can be used to deliver ventilation that targets the low tidal volumes and low plateau pressures that have been demonstrated to decrease mortality for critically ill adults,^11,14^ however, it has been hypothesized that, in practice, however, a ventilator mode that directly controls the tidal volume, such as volume control, may be superior at ensuring the consistent delivery of low tidal volumes, whereas a ventilator mode that directly controls the inspiratory pressure, such as pressure control or adaptive pressure control, may be superior at limiting inspiratory pressure.^35–37^ The results of the current trial appear to confirm this hypothesis and suggest that the choice of ventilator mode in practice may involve a trade-off between optimizing the tidal volume received and optimizing the inspiratory pressure experienced. Whether the observed differences between ventilator modes in tidal volume, inspiratory pressure, and sedation translate into small but clinically meaningful differences in downstream patient outcomes overall, or larger differences in outcome for specific types of patients, remains uncertain.

The current trial has several strengths. Enrollment of all eligible patients in the study location increased generalizability. Adherence to the assigned ventilator mode was assessed frequently (every 1 minute) and was excellent (> 92% compliance in all 3 trial groups). Cluster-level allocation allowed patients to begin receiving the assigned mode at or within minutes of the first breaths of invasive mechanical ventilation in the study unit. This early delivery of the study intervention is important because ventilator settings early in critical illness may have the greatest effects on clinical outcomes.^38,39^ Finally, all aspects of ventilator management aside from ventilator mode were guided by unit-wide protocols that standardized the administration of evidence-based best-practices in mechanical ventilation of critically ill adults.

### Limitations

This trial has several important limitations. Although conducting the trial in a single ICU increased internal validity by facilitating fidelity to the intervention and the collection of detailed data on separation among the groups, it limits generalizability. Patients and clinicians were aware of the assigned ventilator mode. Despite being the largest trial examining ventilator modes to date, the trial had insufficient statistical power to detect small but potentially clinically meaningful treatment effects, overall or for specific types of patients. All trial data were collected within routine clinical care; dedicated physiological measurements of respiratory system compliance and patient-ventilator synchrony were not performed. The trial protocol did not control other aspects of care such as positive end-expiratory pressure or the approach to ventilator weaning. These treatments, however, were standardized according to institutional protocols and did not differ among the groups.

### Conclusions

Among critically ill adults receiving invasive mechanical ventilation, the use of volume control, pressure control, or adaptive pressure control did not affect the number of days alive and free of invasive mechanical ventilation. However, because confidence intervals around the effect estimates included differences of a magnitude that patients or clinicians may find clinically meaningful, further randomized trials are needed.

## Supporting information

Protocol and Analysis Plan

## Data Availability

Deidentified participant data and a data dictionary will be available to researchers whose research proposal is approved by the principal investigator in addition to approval by an Institutional Review Board and an executed data use agreement. Data will become available 3 months following publication of outcomes and will remain available for at least 5 years.

## Funding/Support

KPS was supported by the NIH (T32HL087738); SCD was supported by the NIH (T32GM108554). WHS was supported in part by the NIH/NCATS (UL1 TR00243). TWR was supported by the NIH/NCATS (UL1 TR002243). MWS was supported by the NIH/NCATS (UL1 TR002243). JDC was supported by the NIH (K23HL153584).

## Disclosures

Dr. Casey reported receiving a travel grant from Fisher & Paykel Healthcare to speak at conference. Dr. Qian reported speaking honoraria from Karl Storz Endoscopy. Dr. Rice reported personal fees from Cumberland Pharmaceuticals, Cytovale, and Sanofi. Dr. Semler reporting consulting fees for Baxter International, Inc. No other disclosures were reported.

## Author Contributions

Study concept and design: KPS, BDL, KWG, WHS, ASM, TWR, MWS, JDC. Acquisition of data: KPS, BDL, LW, DTQ, ALM, CES, WDR, TLM, ANT, PGH, TPR, JLS, EMT, DJK, BRD, TKM. Analysis and interpretation of data: KPS, BDL, LW, MSS, ETQ, ALM, SCD, KWG, WHS,ASM, TWR, MWS, JDC. Drafting of the manuscript - original draft: KPS. Critical revision of the manuscript for important intellectual content: All authors. Statistical analysis: KPS, LW, MSS, MWS, JDC. Study supervision: TWR, MWS, JDC. KPS and LW had full access to all the data and take responsibility for the integrity of the data and accuracy of the data analysis. MWS and JDC contributed equally.

## Notes

### Competing Interest Statement

The authors have declared no competing interest.

### Clinical Trial

NCT05563779

### Funding Statement

This study did not receive any funding. KPS was supported by the NIH (T32HL087738); SCD was supported by the NIH (T32GM108554). WHS was supported in part by the NIH/NCATS (UL1 TR00243). TWR was supported by the NIH/NCATS (UL1 TR002243). MWS was supported by the NIH/NCATS (UL1 TR002243). JDC was supported by the NIH (K23HL153584).

### Author Declarations

The institutional review board of Vanderbilt University Medical Center gave ethical approval for this work to be conducted with a waiver of informed consent.

