## Supplemental materials for "Effect of Ventilator Mode on Ventilator-Free Days in Critically Ill Adults: A Randomized Trial"

Online Data Supplement

Table of Contents

*SUPPLEMENTAL METHODS* *4*

Rationale for cluster-level allocation 4

IRB approval and waiver of informed consent 6

Study intervention 9

Study intervention characteristics 11

Approach to monitoring ventilator mode 12

Feedback on ventilator mode adherence 13

Protocol for ventilator management in the study ICU 14

Ventilator characteristics in study unit 18

Protocols for assessment and management of pain, agitation, and delirium 19

Liberation from mechanical ventilation 20

Treatment decisions determined by treating clinicians during the study 21

Trial outcomes – primary outcome 22

Trial outcomes – exploratory outcomes 23

Interim analysis 27

Sensitivity analyses of primary outcome 28

Corrections for multiple testing 29

Effect modification analyses 30

Analysis of exploratory outcomes 32

Acute respiratory distress syndrome definition 33

Assessment of inspiratory airway pressure 34

Sampling of high frequency measures 36

Handling of missing data 37

*SUPPLEMENTAL FIGURES* *38*

Figure E1. Schedule of randomized treatment group assignment 38

Figure E2. Ventilator mode received by each group in the first 168 hours 39

Figure E3. Separation between groups in ventilator mode received in the first 72 hours 40

Figure E4. Separation between groups in ventilator mode received in the first 168 hours 41

Figure E5. Mean tidal volume 42

Figure E6. Proportion of tidal volumes above 8mL/kg predicted body weight 43

Figure E7. Peak inspiratory pressure 44

Figure E8. Mean depth of sedation 45

Figure E9. Proportion of assessments with coma 46
Figure E10. Number of blood gas tests per day 47
Figure E11. Heterogeneity of treatment effect in the primary outcome 48
Figure E12. Effect modification of the primary outcome by age 49
Figure E13. Effect modification of the primary outcome by pre-enrollment $FiO_2$ 50
Figure E14. Effect modification of the primary outcome by baseline SOFA score 51
Figure E15. Mean daily SOFA score 52

*SUPPLEMENTAL TABLES* *53*

Table E1. Complete baseline demographics 53
Table E2. Acute illnesses 54
Table E3. Chronic comorbidities 55
Table E4. Indications for invasive mechanical ventilation 56
Table E5. Lab values at enrollment 57
Table E6. First ventilator settings recorded after enrollment 58
Table E7. Ventilator mode assessments 59
Table E8. Time from enrollment to documentation of any ventilator mode, of the assigned ventilator mode, and of pressure support 61
Table E9. Compliance with group assignment and separation between groups 62
Table E10. Modifications of the mode: number, timing, and rationale 63
Table E11. Daily exploratory outcomes 64
Table E12. Coma, delirium, and sedation on study days 1 to 7 67
Table E13. Other ventilator parameters on study days 1 to 7 70
Table E14. Blood gas laboratory values on study days 1 to 7 71
Table E15. Primary analysis of the primary outcome 73
Table E16. Receipt of supportive therapies to day 28. 74
Table E17. Sensitivity analyses: adjusted primary analysis 75
Table E18. Sensitivity analyses: full cohort including patients enrolled in washout periods 76

*SUPPLEMENT REFERENCES* *77*

### SUPPLEMENTAL METHODS

#### Rationale for cluster-level allocation

Group assignment in the MODE trial occurred at the level of the ICU (cluster) for several reasons. In routine clinical care in the study ICU, 2 to 4 respiratory therapists set the ventilator mode and adjusted settings to maintain gas exchange and patient comfort for all mechanically ventilated adults, with input from nurses and physicians. The management of mechanical ventilation (selection of fraction of inspired oxygen, titration of positive end-expiratory pressure, screening for and performance of spontaneous breathing trials) is governed by unit-wide protocols implemented by the 2 to 4 respiratory therapists for all patients in the unit. Assigning the entire unit to a single ventilator mode in this trial both emulated the way mechanical ventilation was managed during clinical care and limited contamination that might result from a respiratory therapist managing multiple patients assigned to different modes of mechanical ventilation.

Additionally, mechanical ventilation can damage lungs even after brief periods of ventilation, and the initial settings have a unique significance. For example, during the minutes-to-hours of temporary mechanical ventilation for surgical procedures, harmful ventilator settings can affect organ function and clinical outcomes.[1] In critically ill patients, the initial hours of invasive mechanical ventilation are also the period with the highest risk of worsening lung injury, dyssynchrony, increased work of breathing, and hemodynamic compromise. Prior research has demonstrated that [1] the very first ventilator settings at the time the patient is being initiated on invasive mechanical ventilation in the ICU are associated with differences in mortality[2,3] and [2] the association between ventilator settings and mortality is larger for earlier settings compared to later settings.[4]

Recognizing the importance of initial ventilator settings, three recent clinical trials have examined other ventilator settings (i.e. end-expiratory pressure, tidal volume, and $SpO_2$ target) by attempting to enroll patients soon after the initiation of mechanical ventilation in the ICU.[5–7] In these patient-level, parallel-group trials, however, the logistical challenges of performing screening, enrollment, randomization, and study group assignment resulted in a significant gap between initiation of invasive mechanical ventilation and delivery of the respective trial interventions. Those approaches also led to the exclusion of 60-67% of eligible patients – raising concern for systematic exclusion of important patient groups (e.g., patients with higher acuity of illness). In MODE, group assignment at the cluster level allowed enrollment immediately on initiation of invasive mechanical ventilation in the ICU. This approach emulated the manner in which ventilator settings were managed in practice, decreased pre-study exposure to harmful ventilator settings, facilitated early separation in the receipt of ventilator modes between groups, and precluded systematic exclusion of important patient groups. The schedule for the randomized treatment group assignment at the cluster-level is shown in Figure E1.

**IRB approval and waiver of informed consent**

Volume control, pressure control, and adaptive pressure control are all common approaches to controlled mechanical ventilation for critically ill adults. All represent standard-of-care treatments in current clinical practice. Results from prior clinical trials do not demonstrate superiority of one approach over the other. Current clinical guidelines do not recommend any specific mode of mechanical ventilation.[8–11] As a result, significant variation exists in the use of volume control, pressure control, and adaptive pressure control for patients in routine clinical care with ARDS[12] and those without ARDS.[13] In the study ICU, all three modes are used commonly in routine care. This trial enrolled only patients who would already receive continuous mandatory ventilation with one of these three modes as part of their routine clinical care. Whenever the treating clinicians determined that optimal care for the patient would involve a specific ventilator mode different from what was assigned by the study, the team could modify the ventilator mode for the patient and use the mode that they believed was optimal for the patient. Only patients for whom the treating clinicians determined that the ventilator mode assigned by the study was acceptable continued to receive the assigned ventilator mode. We requested and received a waiver of informed consent from the Vanderbilt University Medical Center Institutional Review Board (IRB 220446) because the study involved minimal risk and obtaining informed consent would have made the research study impracticable.

The investigators believed that participation in this study involved minimal risk because: First, the three interventions compared were commonly used in routine clinical care in the study ICU. Second, all were interventions to which the patient could have been exposed even if not participating in the study (all patients initiated on mechanical ventilation in the study ICU receive one of these modes of continuous mandatory ventilation). Third, no established differences in risk and benefits were known to exist between the studied approaches to mechanical ventilation based on the available data. Finally, the trial only determined the mode of ventilation when treating clinicians felt all three modes would be consistent with optimal care for the individual patient – otherwise treating clinicians selected the mode of ventilation via a Mode Modification Sheet.

The MODE trial was designed as a cluster-randomized multiple-crossover trial to ensure that the mode selected at the time of initiation of mechanical ventilation was consistent with trial group assignment, preventing contamination between groups and capturing the period of mechanical ventilation in which ventilator settings have the strongest association with outcomes. Obtaining informed consent before initiation of mechanical ventilation in the study ICU would have been impracticable because:

1) The expected medical condition of patients at the time of initiation of invasive mechanical ventilation in the ICU was critical. All patients were critically ill and receiving continuous intravenous sedation at the time of enrollment. Thus, all patients eligible for MODE lacked the capacity to provide informed consent. Further, data from prior trials in the same patient population and setting demonstrated that prior to intubation and initiation of mechanical ventilation, approximately 70% of patients eligible for the MODE trial would have been experiencing encephalopathy (altered mental status) due to their illness. The anticipated median Glasgow coma scale score was anticipated to be 11 (equivalent to moderate brain injury). Among the minority of patients whose level of consciousness is not impaired, 45-55% were anticipated to be experiencing acute delirium. Further, family members or legally authorized representatives (LAR) are frequently unavailable when critically ill patients undergo intubation and initiation of invasive mechanical ventilation in the ED or ICU.
2) The intervention is delivered by unit-level protocols. Mechanical ventilator settings and titrations are performed by respiratory therapists using unit-level protocols. In this cluster-randomized trial, the entire unit was assigned to the same mode of mechanical ventilation for each 1-month block. Obtaining informed consent from every eligible patient in the ICU prior to emergency tracheal intubation and initiation of mechanical ventilation would have been impracticable.

Because the study involved minimal risk, the study did not adversely affect the welfare or privacy rights of the participant, and obtaining informed consent would have been impracticable, the investigators requested a waiver of informed consent.

Numerous previous randomized trials comparing two standards of care for interventions such as emergency intubation and methods of respiratory support during critical illness have been completed with waiver of informed consent.[14–22]

**Study intervention**

The MODE trial compared volume control, pressure control, and adaptive pressure control (Table 1). On initiation of invasive mechanical ventilation in an intensive care unit, all patients require a ventilator mode that initiates breaths at a set frequency (mandatory mode of ventilation). The three most commonly used modes of mandatory ventilation are volume control, pressure control, and adaptive pressure control. Each of these modes provides continuous mandatory ventilation, in which every inspiratory effort by a patient triggers a machine-cycled breath delivered by the ventilator, and a set minimum respiratory rate is maintained by machine-triggered breaths as needed.[23] The study interventions for each study group assignment were as follows.

**Volume Control Group:** Patients assigned to the volume control group received volume control as the mode of ventilation whenever they were receiving continuous mandatory invasive mechanical ventilation.

**Pressure Control Group:** Patients assigned to the pressure control group received pressure control as the mode of ventilation whenever they were receiving continuous mandatory invasive mechanical ventilation.

**Adaptive Pressure Control Group:** Patients assigned to the adaptive pressure control group received adaptive pressure control as the mode of ventilation whenever they were receiving continuous mandatory invasive mechanical ventilation.

Characteristics of each of the three modes of ventilation are presented in Figure E2. For patients enrolled in the MODE trial, clinicians were instructed to use the assigned mode beginning at the first receipt of invasive mechanical ventilation in the study ICU and ending at the first of: (1) extubation from mechanical ventilation, (2) transfer out of the study ICU, (3) end of the one-month study block. The trial protocol did not determine the ventilator mode during time-periods in which the patient was not receiving a continuous mandatory mode of ventilation (e.g., while receiving pressure support ventilation during a spontaneous breathing trial), was not physically located in the study ICU (e.g., during transport), or was undergoing an invasive procedure (e.g., bronchoscopy). For patients who continued to receive mechanical ventilation in a trial location after the end of a study block, the ventilator mode was selected by treating clinicians. If, at any time, a treating clinician, patient, or family member determined that a ventilator mode other than that assigned by the trial might be best for the treatment of the patient, the ventilator mode for that patient was modified and the reason for modifying the ventilator mode was recorded. If a patient was enrolled, extubated, and re-intubated during the same study block, the study protocol determined the ventilator mode until they met one of the criteria listed above.

In the study ICU, respiratory therapists typically have primary responsibility for determining the initial settings for invasive mechanical ventilation and titrating the settings to achieve clinical goals (e.g., $SaO_2$, $pCO_2$, target tidal volume, target plateau pressure, etc.). To set and titrate mechanical ventilators, respiratory therapists use clinical protocols jointly developed by respiratory therapy and physician leaders. For patients enrolled in the MODE trial, respiratory therapists employed the same pre-existing clinical protocols regardless of group assignment (volume control, pressure control, or adaptive pressure control).

**Study intervention characteristics**

| Group Assignment | Volume Control | Pressure Control | Adaptive Pressure Control |
|---|---|---|---|
| Ventilator Mode Designation on Servo Ventilators | Volume Control | Pressure Control | Pressure Regulated Volume Control |
| Breath Control Variable[a] | Volume | Pressure | Pressure |
| Breath Sequence[a] | Continuous mandatory ventilation | Continuous mandatory ventilation | Continuous mandatory ventilation |
| Targeting Scheme[a] | Set-point (clinician sets the tidal volume) | Set-point (clinician sets the inspiratory pressure) | Adaptive (clinician sets target tidal volume and ventilator titrates pressure to achieve target tidal volume) |

a Taxonomy of ventilator mode characteristics as organized in Chatburn et al.[23]

### Approach to monitoring ventilator mode

For all mechanically ventilated patients in the study ICU, ventilators displayed settings such as mode of ventilation and measurements such as observed tidal volume on a digital display at the patient's bedside in real time. For critical measurements such as respiratory rate, minute ventilation, and peak pressures, audible alarms from the ventilator alerted clinicians[21,22] to potentially dangerous situations.

These settings and measurements were automatically uploaded into the electronic medical record and stored in a data warehouse. This allowed for highly representative assessments of the proportion of time that patients received each ventilator mode over the course of the day compared to prior trials which documented ventilator mode once or twice daily.[24,25]

**Feedback on ventilator mode adherence**

During the study, study personnel monitored compliance with the assigned ventilator mode. Study personnel remotely monitored ventilator modes up to four times daily from 5AM through 11PM during weekdays and twice daily during weekends to provide feedback to treating clinicians and explore reasons why patients were not receiving the assigned mode of mechanical ventilation. Study personnel also interacted with physicians, nurses, and respiratory therapists at least once daily during weekdays to solicit safety concerns and adverse events and to identify and address barriers to ventilator mode assignment compliance.

In addition, study personnel attended respiratory therapy group meetings, nursing unit board meetings, and ICU physician meetings to educate staff about the study and receive input on conduct of the trial.

**Protocol for ventilator management in the study ICU**

SUMMARY OF VUMC MICU VENTILATOR PROTOCOL GUIDELINES:

Ventilator Set Up and Adjustment:

1. Calculate Predicted Body Weight (PBW):
    a. In males = 50 + 2.3 (height (inches) - 60)
    b. In females = 45.5 + 2.3 (height (inches) - 60)

2. Select Mode and initial default settings:

    a. **Volume Control.** Set Tidal Volume (Vt) to target 6 mL/kg PBW. Set Flow Pattern: 100 or [square-wave icon with flow adaptation feature]; Set T pause (s): 0.

    b. **Pressure Regulated Volume Control**. Set Vt to target 6 mL/kg PBW.

    c. **Pressure Control.** Set PC above PEEP to 15 $cmH_2O$, set inspiratory time to 0.9 s and confirm delivered Vt is appropriate for target 6 mL/kg PBW.

3. Set initial respiratory rate (RR) to approximate baseline Minute Ventilation (12-16 bpm, not > 35 bpm).

4. Set alarms:

    For **VC and PRVC**: High Peak Pressure alarm at 40 cm $H_2O$

    For **PC**: Low Minute Ventilation alarm at 2 L/min less than the current Minute Ventilation

5. Adjust Vt and RR to achieve pH and plateau pressure goals below.

    Lung Protective Ventilation Goals – Vt ≤ 6 mL/kg PBW, Pplat < 30 $cmH_2O$

        *For Pressure Control, where Vt cannot be directly adjusted, adjust PC over PEEP in increments of 2 $cmH_2O$ and monitor resulting Vt to achieve the changes outlined below.

        If Vt > 6 mL/kg PBW: Reduce Vt by 1 mL/kg at intervals ≤ 2 hours until Vt = 6 mL/kg PBW.

If Pplat > 30 (or PIP > 35): Decrease Vt by 1 mL/kg steps to a minimum of 4mL/kg PBW.

If Pplat < 25 and Vt < 6 mL/kg: Increase Vt by 1 mL/kg until either Pplat > 25 or Vt = 6 mL/kg PBW.

If Pplat <30 and breath stacking occurs: May increase Vt in 1 mL/kg increments to maximum of 8 mL/kg PBW.

Ventilation Goals - pH: 7.30-7.45

If pH 7.15-7.30: Increase RR until pH > 7.30 or $PaCO_2$ < 25 (Max RR = 35).

If RR = 35 and $PaCO_2$ < 25: May give $NaHCO_3$, to be managed by primary team.

If pH < 7.15: Increase RR to 35.

If pH remains < 7.15 and $NaHCO_3$ considered or infused: Vt may be increased until pH >7.15 or Vt = 8 mL/kg PBW (Pplat target may be exceeded).

If pH >7.45: Decrease RR rate.

Oxygenation goals - $SpO_2$: 88-95% or $PaO_2$: 55-80 mmHg.

Use incremental $FiO_2$/PEEP combinations below to provide minimum $FiO_2$ and PEEP necessary to achieve $PaO_2$ or $SpO_2$ goals. Use of a high PEEP table is allowed and may be used in specific cases (e.g., severe ARDS).

| | | | | | | | | | | | | | |
|---|---|---|---|---|---|---|---|---|---|---|---|---|---|
| $FiO_2$ | 0.3-0.4 | 0.4 | 0.5 | 0.5 | 0.6 | 0.7 | 0.7 | 0.7 | 0.8 | 0.9 | 0.9 | 0.9 | 1 |
| PEEP | 5 | 8 | 8 | 10 | 10 | 10 | 12 | 14 | 14 | 14 | 16 | 18 | 18-25 |

Other Settings

**I:E ratio goal:** 1:1.0 – 1:3.0. Adjust Ti to achieve goal.

If $FiO_2$ = 1.0 and PEEP = 24 $cmH_2O$, may adjust I:E to 1:1.

Avoid inverse-ratio ventilation.

**Trigger:** 1.6 L/min by default, can be increased or reduced to improve synchrony

**Tinsp rise (s):** 0.15 s by default, can be increased or decreased to improve ventilation

Monitoring and Documentation:

Measure & record Pplat with inspiratory pause of at least 0.5 s, $SpO_2$, Total RR, Vt and pH (if available) at least every 4 hours AND after each change in PEEP, Vt, or PC above PEEP settings.

Once the patient tolerates $FiO_2 \leq 0.5$ and PEEP ≤ 8 $cmH_2O$, advance to Part II: Spontaneous Breathing Trial Readiness Assessment and Ventilator Discontinuation Guidelines.

Ventilator management guidelines in the study ICU summarized:

| | Volume Control | Pressure Control | Adaptive Pressure Control |
|---|---|---|---|
| Mode option | Volume Control | Pressure Control | Pressure Regulated Volume Control (PRVC) |
| Tidal Volume target (mL/kg PBW) | 6 | 6 | 6 |
| Tidal Volume range (mL/kg PBW) | 4 - 8 | 4 - 8 | 4 - 8 |
| Initial breath settings | Tidal volume to 6 mL/kg PBW | PC over PEEP to 15 cm H2O, Insp Time to 0.9s | Tidal volume to 6 mL/kg PBW |
| Achieving target Tidal Volume | Directly adjust tidal volume | Dependent on adjustment of PC over PEEP | Directly adjust tidal volume target |
| Plat Pressure target (cm $H_2O$) | ≤ 30 | ≤ 30 | ≤ 30 |
| Achieving target Plateau Pressure | Dependent on adjustment of tidal volume | Directly adjust PC over PEEP | Dependent on adjustment of tidal volume |
| Flow | Set directly | Dependent on ventilator and patient factors | Dependent on ventilator and patient factors |
| Flow Pattern | Square waveform | Decelerating, dynamic | Decelerating, dynamic |
| Arterial pH goal | 7.30 - 7.45 | 7.30 - 7.45 | 7.30 - 7.45 |
| Ventilator rate (breaths/min) | ≤ 35 | ≤ 35 | ≤ 35 |
| Oxygenation goal by $PaO_2$ (mmHg) | 55 - 80 | 55 - 80 | 55 - 80 |
| Oxygenation goal by $SpO_2$ (%) | 88 - 95 | 88 - 95 | 88 - 95 |
| Positive end-expiratory pressure | Set with $FiO_2$-PEEP table | Set with $FiO_2$-PEEP table | Set with $FiO_2$-PEEP table |
| Inspiratory: Expiratory ratio | 1:1 - 1:3 | 1:1 - 1:3 | 1:1 - 1:3 |
| Extubation evaluation | MICU SBT Readiness and Extubation Guidelines | MICU SBT Readiness and Extubation Guidelines | MICU SBT Readiness and Extubation Guidelines |

### Ventilator characteristics in study unit

Patients in this study received mechanical ventilation using the mechanical ventilators that they would have received as part of routine clinical care. All mechanical ventilators were either Getinge Servo-U or Servo-I ventilators. The adaptive pressure control intervention was implemented with Pressure Regulated Volume Control (PRVC) mode.

**Protocols for assessment and management of pain, agitation, and delirium**

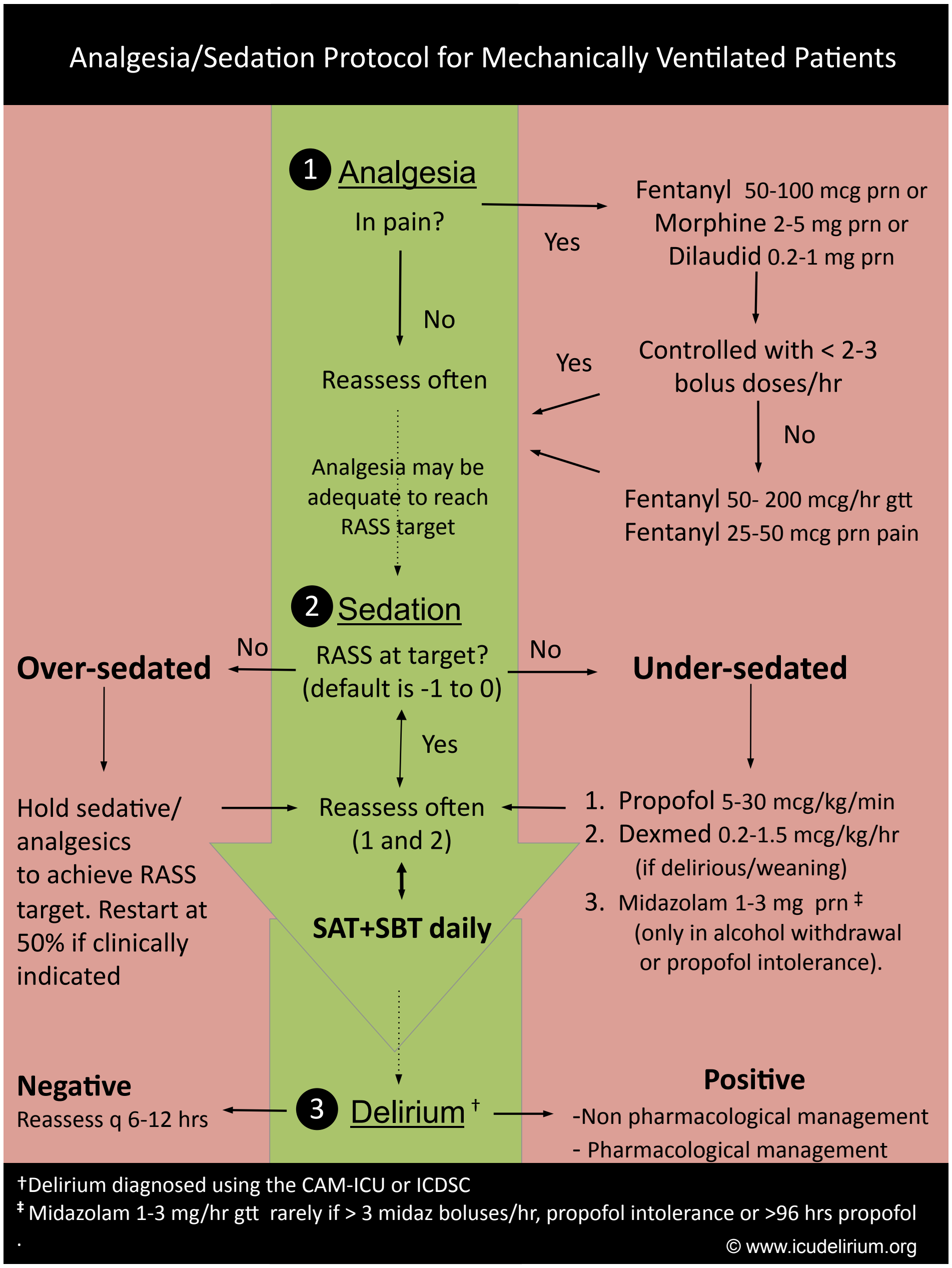

Reproduced with permission from: https://www.icudelirium.org/medical-professionals/choice-of-analgesia-and-sedation

#### Liberation from mechanical ventilation

Each day of mechanical ventilation, all patients in the study ICU are assessed for safety of a spontaneous awakening trial (SAT) and spontaneous breathing trial (SBT)[26] using the SAT and SBT safety criteria from the Awakening and Breathing Controlled (ABC) trial.[27] These criteria relate to the non-respiratory clinical status of the patient and the PEEP and $FiO_2$ settings which were managed similarly across groups. As such, the SAT and SBT procedures were handled in the same way for each patient, regardless of study group assignment. Definitions of SAT and SBT failure and the ventilator settings and duration of the SBT were those used in the ABC trial and used in routine clinical care in the study ICU.  For patients who passed an SAT and SBT, the decision to discontinue invasive mechanical ventilation was made by the treating clinicians.

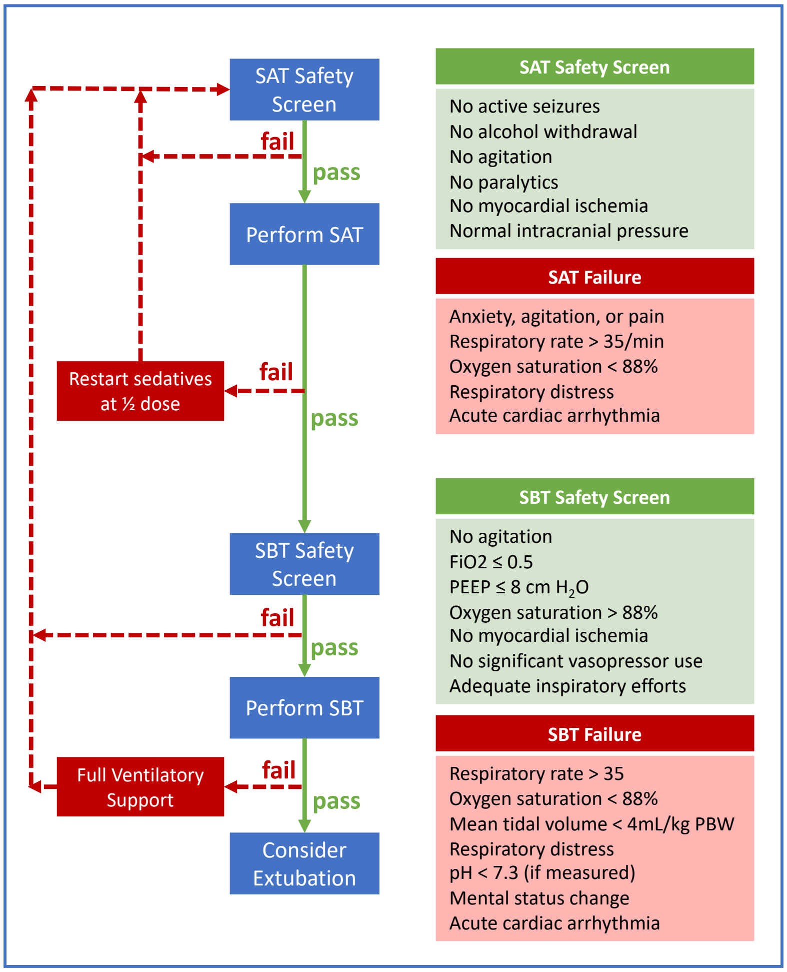

Adapted by the study ICU from the "Wake up and Breathe Flowchart" found at: https://www.icudelirium.org/medical-professionals/both-sat-and-sbt

**Treatment decisions determined by treating clinicians during the study**

All treatment decisions except choice of mode for continuous mandatory ventilation during invasive mechanical ventilation were made by treating clinicians, including: approach to supplemental oxygen therapy and non-invasive positive pressure ventilation prior to invasive mechanical ventilation, choice of oxygen saturation targets, use of spontaneous modes of ventilation such as pressure support, approach to positive end-expiratory pressure during invasive mechanical ventilation, prone positioning, and extracorporeal membrane oxygenation; administration of analgesia and sedation, neuromuscular blocking agents, inhaled epoprostenol, vasopressors, inotropes, antimicrobial medications, diuretics, intravenous fluids, and blood products; measurement of lactate and arterial or venous blood gas.

**Trial outcomes – primary outcome**

Ventilator-free days was defined as the number of whole calendar days alive and free of invasive mechanical ventilation from the final receipt of invasive mechanical ventilation through day 28 after enrollment.[28,29] Outcome ascertainment ceased at the time of hospital discharge or 28 days after enrollment, whichever occurred first. Receipt of invasive mechanical ventilation was considered to end when patients underwent the final tracheal extubation or disconnection of the ventilator from the endotracheal tube or tracheostomy tube between enrollment (day 1) and day 28 day after enrollment. Patients who died prior to discharge on or before day 28 received 0 VFDs. Patients whose final receipt of invasive mechanical ventilation occurred on the day of enrollment and survived to day 28 received 27 VFDs. Patients who continued to receive invasive mechanical ventilation 28 days after enrollment received 0 VFDs. Patients who were discharged from the hospital prior to day 28 and were receiving invasive mechanical ventilation at the time of discharge received 0 VFDs. Patients who were removed from invasive mechanical ventilation and were discharged from the hospital without invasive mechanical ventilation prior to 28 days were assumed to remain alive and free of invasive mechanical ventilation between hospital discharge and day 28. For patients who were removed from invasive mechanical ventilation, returned to invasive mechanical ventilation, and were subsequently removed again from invasive mechanical ventilation prior to day 28, VFDs were counted from the final receipt of invasive mechanical ventilation prior to day 28.

**Trial outcomes – exploratory outcomes**

Safety outcomes:

- Hypoxemia during mechanical ventilation, defined as having an $SpO_2$ < 85% for more than 5 minutes
- Severe acidemia during mechanical ventilation, defined as having a pH < 7.1 on blood gas (arterial or venous)
- Pneumomediastinum or pneumothorax during course of mechanical ventilation

Exploratory on-study outcomes:

- Median exhaled tidal volume (mL/kg predicted body weight) on each study day
- Exhaled tidal volumes above target range: proportion of recorded breaths with exhaled tidal volume values above the target range (>8mL/kg predicted body weight) on each study day
- Number of blood gas laboratory tests per day while receiving mechanical ventilation
- SOFA score daily on the first 7 study days

Exploratory feasibility outcomes:

- Exposure to assigned study mode in the first 3 days: proportion of time in the assigned mode while receiving invasive mechanical ventilation in the study ICU between enrollment and 72 hours after enrollment (including time spent in spontaneous modes such as during a spontaneous breathing trial)
- Adherence to study mode in the first 3 days: proportion of time in the assigned mode while receiving invasive mechanical ventilation in the study ICU with a mandatory mode between enrollment and 72 hours after enrollment (excluding time spent in spontaneous modes such as during a spontaneous breathing trial)
- Time from enrollment to initiation of assigned mode of mechanical ventilation
- Receipt of a “mode modification sheet” completed by treating clinicians at any time after enrollment when the ventilator mode was determined by group assignment.

Exploratory clinical outcomes:

- Delirium and coma-free days to day 28
- ICU-free days to study day 28
- Hospital-free days to study day 28
- In-hospital mortality to study day 28

**Exploratory outcome definitions**

Delirium and coma-free days through study day 28: Delirium and coma-free days was defined as the number of calendar days the patient was alive and free of delirium and coma from enrollment through day 28 after enrollment. Delirium was defined as a positive assessment of the CAM-ICU.[30] Coma was defined as a RASS of -4 or -5.[31,32] The number of days alive and free of delirium and coma was defined as the number of calendar days alive and free of both delirium and coma through day 28 using the "last off method." The day of enrollment was considered to be day 1. Outcome ascertainment ceased at the time of hospital discharge or 28 days, whichever came first. Delirium and coma-free days were considered to begin the day following the patient's final recorded positive assessment of the CAM-ICU or RASS of -4 or -5 between enrollment and day 28. Patients who continued to have delirium or coma at day 28 after enrollment received a value of 0. Patients who died on or before day 28 after enrollment received a value of 0. Patients discharged from the hospital prior to day 28 after enrollment while still having delirium or coma received a value of 0. Patients who were discharged from the hospital prior to day 28 after enrollment without delirium or coma were assumed to remain free of delirium and coma between hospital discharge and day 28 after enrollment. For patients who had a period without delirium and coma and subsequently developed or re-developed delirium or coma, days alive and free of delirium and coma were counted from the final day of delirium or coma through day 28 after enrollment.

ICU-free days through study day 28: Number of days alive and free of the ICU were defined as the number of calendar days in which the patient was alive between the patient's final transfer or discharge from an ICU service and day 28 after enrollment. Patients who were never discharged from the intensive care unit received a value of 0. Patients who died before or on day 28 received a value of 0. Patients who returned to an intensive care unit service and were subsequently discharged prior to day 28, ICU-free days were considered to begin the day following the date of final ICU discharge. All data were censored at hospital discharge or 28 days, whichever came first.

Hospital-free days to study day 28: Number of days alive and free of the hospital were defined as the number of calendar days between enrollment and 28 days after enrollment, in which the patient was alive and not admitted to the hospital. Patients who were never discharged from the hospital received a value of 0. Patients who died before day 28 received a value of 0. All data were censored at hospital discharge from the hospitalization in which they were enrolled in the trial or 28 days, whichever came first.

Median exhaled tidal volume on each study day: Tidal volumes were reported in mL/kg of Predicted Body Weight (PBW), calculated using the following equations: PBW for Males = 50 + 2.3 * [height(inches)-60] and PBW for Females = 45.5 +2.3 * [height(inches) -60].

**Interim analysis**

The DSMB conducted a single, planned interim analysis for safety on March 22, 2023 to review data from patients enrolled after the ICU had been assigned to each of the three study modes, that is, after enrollment of patients for 3 months. According to criteria specified in the trial protocol, the stopping boundary for safety would have been met if the p-value for a difference between the groups in any of the three safety outcomes met the threshold of 0.016 or less. There was no stopping boundary for efficacy or futility.

The DSMB evaluated the incidence of the following safety outcomes across the study groups (volume control, pressure control, and adaptive pressure control): pneumomediastinum or pneumothorax during the course of mechanical ventilation, episodes of hypoxemia while receiving invasive mechanical ventilation: $SpO_2$ < 85% for more than 5 minutes, and in-hospital mortality to study day 28.

After conducting the planned interim analysis, the DSMB recommended the study continue without modification. The DSMB reserved the right to stop the trial at any point, request additional data or interim analyses, or request modifications of the study protocol as required to protect safety.

**Sensitivity analyses of primary outcome**

Adjusted analysis: We repeated the primary analysis with adjustment for pre-specified baseline covariates of: age (continuous), sex (male, female), race and ethnicity (Hispanic, Non-Hispanic Black, Non-Hispanic White, Other), source of ICU admission (ED, hospital ward, another ICU in the study hospital, operating room, outside hospital), vasopressor receipt (yes, no), and acute diagnoses at enrollment (cardiac arrest, sepsis or septic shock, pneumonia, COPD exacerbation, asthma exacerbation), and severity of illness as assessed by SOFA score. To account for non-linear relationships, continuous variables were analyzed using restricted cubic splines with between 3 and 5 knots.

Analysis including patients from washout periods: We repeated the analysis of the primary outcome among all patients enrolled in the trial including those initiated on invasive mechanical ventilation in the study ICU during one of the pre-specified 3-day washout periods.

### Corrections for multiple testing

We pre-specified a single primary clinical outcome, which provided a test of statistical significance for difference between groups. Consistent with recommendations of the Food and Drug Administration and the European Medicines Association, the clinical outcome was tested using a two-sided p-value with a significance level of 0.05. For all other analyses, emphasis was placed on the estimate of effect size with 95% confidence intervals, as recommended by the International Committee of Medical Journal Editors, and no corrections for multiple comparisons will be performed.

**Effect modification analyses**

We prespecified the following baseline variables as potential modifiers of the effect of study group on the primary outcome and hypothesized the direction of the effect modification for each to follow the Instrument to assess the Credibility of Effect Modification Analyses (ICEMAN) criteria.[33]:

1. Age (continuous variable). We hypothesized that age would not modify the effect of ventilator mode on the number of ventilator free days.

2. Duration of invasive mechanical ventilation prior to enrollment (0 minutes; 1 to 360 minutes; >360 minutes). We hypothesized that duration of invasive mechanical ventilation would not modify the effect of ventilator mode on the number of ventilator free days.

3. Pre-enrollment fraction of inspired oxygen ($FiO_2$) (continuous variable). We hypothesized that the $FiO_2$ received pre-enrollment would modify the effect of study mode on the outcome of ventilator free days with a greater increase in the number of ventilator free days in the volume control group among patients with a higher $FiO_2$ compared to patients with lower $FiO_2$. This hypothesis was supported by the rationale that evidence-based practices for lung protective ventilation were established in prior trials that placed a priority on limiting tidal volumes, for which an effect was most pronounced among cohorts with severe hypoxemic respiratory failure.[5,34]

4. SOFA score at enrollment (continuous variable). We hypothesized that baseline SOFA score would not modify the effect of ventilator mode on the number of ventilator free days.

5. Shock receiving vasopressors (yes, no); We hypothesized that shock would not modify the effect of ventilator mode on the number of ventilator free days.

6. Indications for intubation (categories are not mutually exclusive)

   a) Hypoxemic respiratory failure (yes, no); We hypothesized that hypoxemic respiratory failure as a reason for intubation would modify the effect of ventilator mode on the primary outcome among the volume control group with the same justification provided above with $FiO_2$.

b) Hypercarbic respiratory failure (yes, no); We hypothesized that hypercarbic respiratory failure would not modify the effect of ventilator mode on the number of ventilator free days.
c) Altered mental status or airway protection (yes, no); We hypothesized that altered mental status or airway protection as a reason for intubation would modify the effect of study mode on the outcome of ventilator free days with a greater increase in the number of ventilator free days among the pressure control group because the hypothesized effects of improved patient-ventilator synchrony, increased patient comfort, and reduced sedation use with pressure control may be more likely to reduce the duration of mechanical ventilation among patients primarily intubated for altered mental status.[35,36]
d) Chronic Obstructive Pulmonary Disease (yes, no); We hypothesized that chronic obstructive pulmonary disease would not modify the effect of ventilator mode on the number of ventilator free days.

In post hoc analyses, we assessed whether the presence of acute respiratory distress syndrome (ARDS) criteria at enrollment modified the effect of study group on the primary outcome. We hypothesized that the presence of ARDS at enrollment would modify the effect of study group with a greater increase in the number of ventilator free days in the volume control group among patients meeting ARDS criteria. This hypothesis was supported by the rationale that evidence-based practices for lung protective ventilation were established in prior trials that placed a priority on limiting tidal volumes, for which an effect was most pronounced among cohorts with ARDS.[5,34]

**Analysis of exploratory outcomes**

As per the protocol and statistical analysis plan, each of the exploratory outcomes were compared between groups in an intention-to-treat fashion with an approach similar to that used for the primary outcome.[37] A proportional-odds model was used for ordinal and continuous outcomes with independent covariates of group assignment and time (analyzed as the number of days since study initiation). Differences between each pair of trial groups were estimated by extracting odds ratios and 95% confidence intervals from the model.

To summarize repeated measures, a single summary measure was calculated for each patient in the trial (e.g., a mean of all tidal volume values for a patient). These mean values for each patient were summarized as the median and interquartile range among patients in each of the three trial groups (e.g., the median of the mean tidal volume among patients in the volume control group). To test for differences between groups using all of the repeated measures data, generalized estimating equations were used to adjust for within-subject correlation, including independent variables of study group, time (number of days since study initiation), and a term for the interaction of group and study day. The 95% confidence intervals around the difference in medians for continuous variables were calculated using bootstrapping.

**Acute respiratory distress syndrome definition**

We assessed for acute respiratory distress syndrome (ARDS) criteria at enrollment using the New Global Definition of ARDS[38] for intubated patients: [1] Acute onset of hypoxemic respiratory failure as defined as new or worsening respiratory symptoms within 1 week of a predisposing risk factor (e.g., pneumonia, trauma, transfusion), or within one week of new or worsening respiratory symptoms which are not attributable to cardiogenic pulmonary edema, fluid overload, atelectasis, pleural effusion, or pulmonary embolism; [2] Bilateral opacities on chest radiography or computed tomography not fully explained by effusions, atelectasis or nodules/masses; and [3] Hypoxemia as defined by either a $PaO_2/FiO_2$ ratio of ≤ 300 or $SpO_2/FiO_2$ ratio of ≤ 315 when a $PaO_2$ value from arterial blood gas testing was not available and $SpO_2$ values were ≤ 97% while receiving positive end expiratory pressure of at least 5 centimeters of water pressure. Determination of these criteria was completed for each patient by a blinded investigator using data collected during clinical care, prioritizing values at the time of enrollment but utilizing data up to six hours before or after enrollment.

#### Assessment of inspiratory airway pressure

Barotrauma is an important potential mechanism for differences in clinical outcomes between groups in the MODE trial. Lung-distending pressures and the peak inspiratory pressures may both contribute to the risk of barotrauma.[39,40]

Lung-distending pressure is typically measured with an extended end-inspiratory hold, which provides a plateau pressure. During an inspiratory hold, gas pressures equilibrate across the respiratory system from the alveoli to the ventilator circuit and pressure sensor which measures the plateau pressure.[41,42] This static measurement estimates the lung-distending pressure experienced by alveoli, but the measurement is also affected by the chest wall.[43] In critically ill adults, regional heterogeneity of the lung may lead to heterogeneous alveolar pressures, such that different regions of the lung may be experiencing distending pressures higher or lower than suggested by the measured plateau pressure.[44] Maintaining a plateau pressure of less than 30 $cmH_2O$ is specified as a target for the lung protective ventilation strategy used in the study unit, but plateau pressures are not measurable in clinical care for the most of the intervention period of this trial. Valid plateau pressure measurement requires a passive patient, meaning that patients must be deeply sedated or paralyzed. However, trials have shown potential benefits from daily spontaneous awakening trials and light sedation, and most patients receiving mechanical ventilation in the study unit exhibit spontaneous respiratory effort for all or most of their course of mechanical ventilation.[40,42,45] Furthermore, because plateau pressure measurements requires effort from clinicians, measurements are more likely to be obtained for patients who are more severely ill or experiencing clinical deterioration (sampling bias) (see Tables E6 and E13). Finally, measurements of plateau pressure are subject to differences in technique.[46]

Peak inspiratory pressures, in contrast, are automatically measured with every breath for every patient as part of routine clinical care during mechanical ventilation. The peak inspiratory pressure represent the pressure gradient between the ventilator and the alveoli, and it includes both the pressure required for lung distention and the pressure required to overcome the resistive forces of the ventilator tubing and airways. Among critically ill adults experiencing regional heterogeneity of the lung, peak inspiratory pressures represent the maximal, potentially injurious, positive pressure that any part of the lung may be experiencing from the mechanical ventilator.

While some have proposed that the plateau pressure is the inspiratory pressure most directly correlated with risk of barotrauma,[47] it cannot be measured for the majority of patients during the majority of their course of mechanical ventilation. When they are obtained as part of routine clinical care, plateau pressure measurements are infrequent and are subject to sampling bias. Because (1) they are available at all times and in all patients, (2) are sampled frequently as part of routine clinical care, (3) are collected in an unbiased manner, and (4) represent the maximal, potentially injurious, positive pressure that any part of the lung may be experiencing from the mechanical ventilator, peak inspiratory pressures were prioritized as the primary inspiratory pressure measurement in the MODE trial.

**Sampling of high frequency measures**

Ventilator mode and $SpO_2$ were recorded approximately every minute. A total of 1,405,218 assessments of ventilator mode were performed, with a median interval of 1 minute (interquartile range(IQR), 1 to 1) between assessments.

A total of 1,324,960 $SpO_2$ measurements were performed, with a median interval of 1 minute (IQR, 1 to 1) between assessments.

Other mechanical ventilator settings such as tidal volumes and peak inspiratory pressure were recorded approximately every hour. A total of 57,228 tidal volume assessments were recorded with a median interval of 56 minutes (IQR, 34 to 60). A total of 135 (0.2%) tidal volume measurements from 65 patients had a value greater than 1500 mL and were considered to be erroneous and imputed as missing. A total of 54,713 measurements of peak inspiratory pressure were recorded with a median interval of 58 minutes (IQR, 38 to 60).

**Handling of missing data**

The primary outcome of VFDs was not missing for any patients. Missing data were not imputed for any exploratory outcomes. For analyses of tidal volume in mL/kg PBW, height was imputed when missing for calculations of PBW with multiple imputations using all other available baseline characteristics.

None of the covariates pre-specified for the adjusted analyses were missing for any patients.

### SUPPLEMENTAL FIGURES

#### Figure E1. Schedule of randomized treatment group assignment

| Block 1 | | | Block 2 | | | Block 3 | | |
|---|---|---|---|---|---|---|---|---|
| 2022 | | 2023 | | | | | | |
| November | December | January | February | March | April | May | June | July |
| Volume Control | Pressure Control | Adaptive Pressure Control | Adaptive Pressure Control | Pressure Control | Volume Control | Pressure Control | Volume Control | Adaptive Pressure Control |

For each of the nine one-month study periods, the ICU was randomly assigned to a study mode (volume control, pressure control, and adaptive pressure control) for mandatory ventilation.

**Figure E2. Ventilator mode received by each group in the first 168 hours**

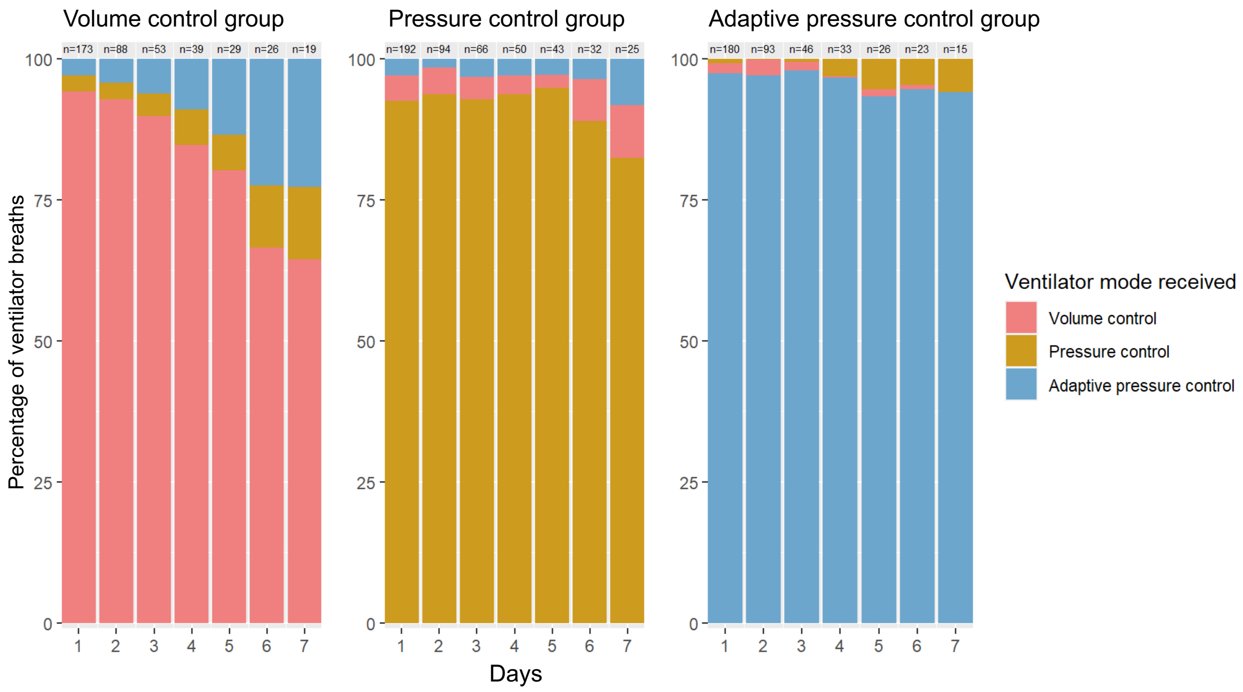

Shown are the percentages of ventilator breaths in each trial group that were in volume control mode (red), pressure control mode (yellow), and adaptive pressure control mode (blue) for the 168 hours following enrollment. Ventilator mode was assessed approximately every 1 minute. This figure displays data on breaths for which the patient was receiving a continuous mandatory mode of ventilation. Also shown is the number of patients who were alive and receiving a continuous mandatory mode of ventilation in each group during each time interval. Data including spontaneous modes are in Figures E3 and E4 and Table E7.

**Figure E3. Separation between groups in ventilator mode received in the first 72 hours**

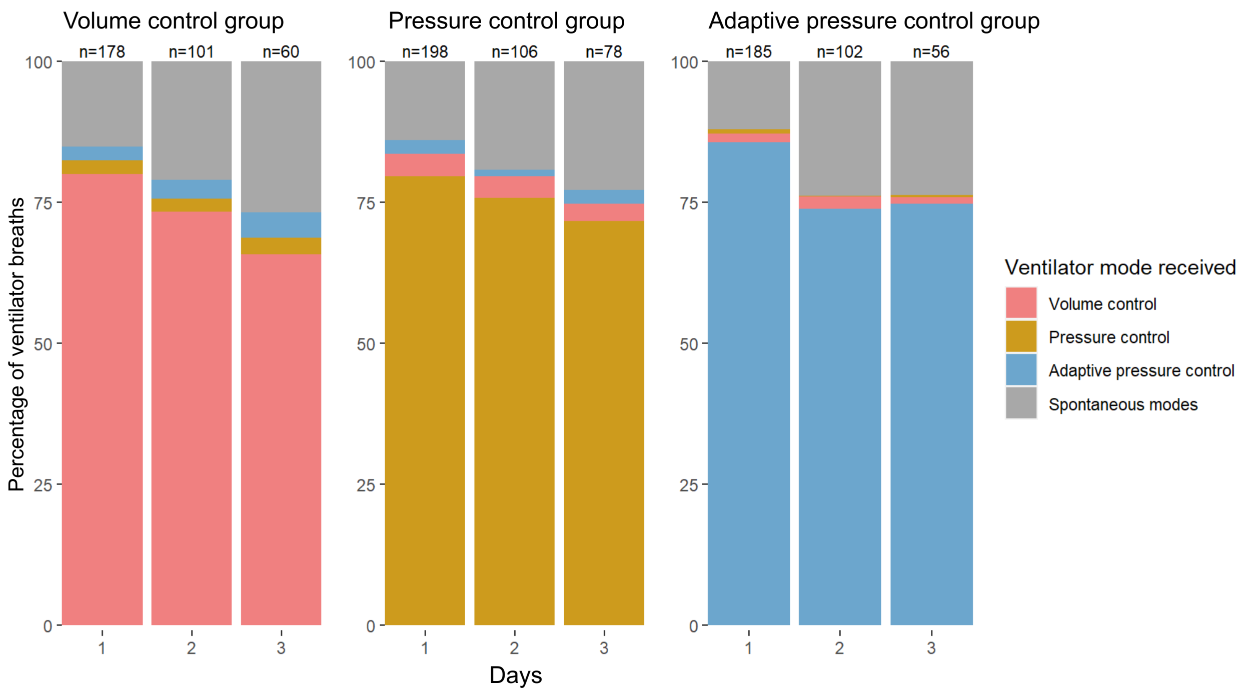

Shown are the percentages of all ventilator breaths assessed in each trial group by ventilator mode: volume control mode (red), pressure control mode (yellow), adaptive pressure control mode (blue), and spontaneous modes (e.g., pressure support) (grey) for the 72 hours following enrollment.  Ventilator mode was assessed approximately every 1 minute. Also shown is the number of patients who were alive and receiving invasive mechanical ventilation in each group during each time interval. Ventilator mode is reported for 561 of 566 patients on day 1. Two patients were missing data on ventilator mode on study day 1 but contributed data on subsequent days, and three patients were missing data on all days.

**Figure E4. Separation between groups in ventilator mode received in the first 168 hours**

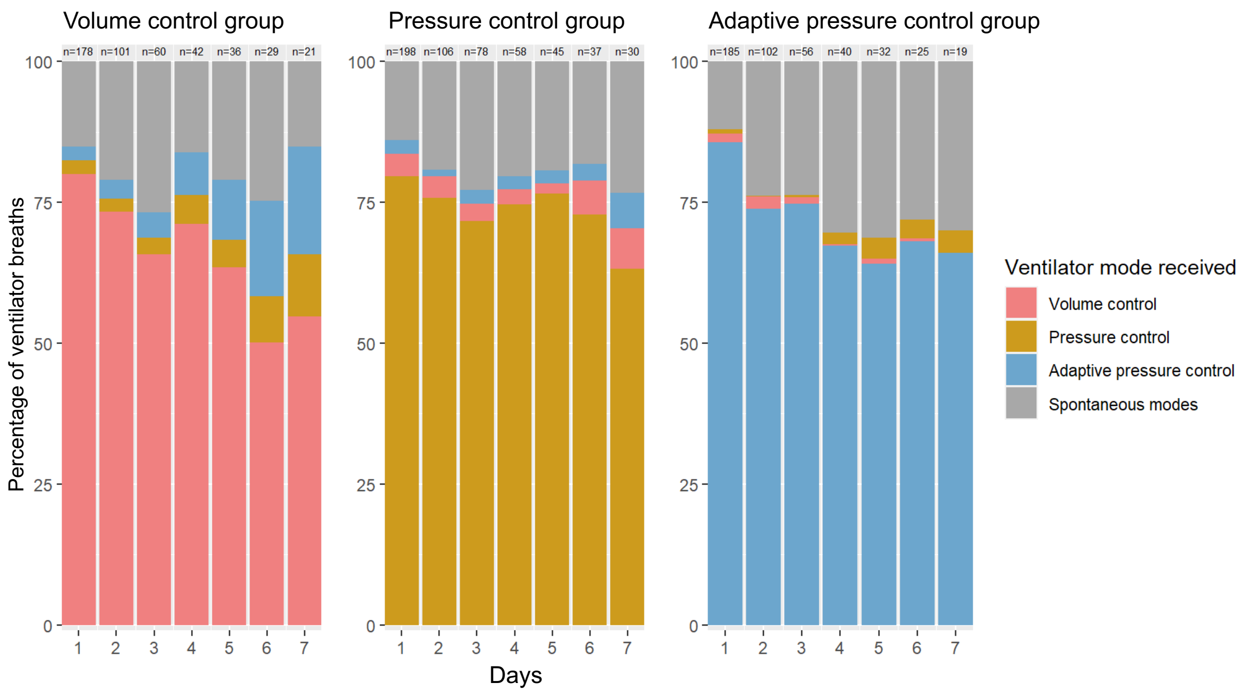

Shown are the percentages of all ventilator breaths assessed in each trial group by ventilator mode: volume control mode (red), pressure control mode (yellow), adaptive pressure control mode (blue), and spontaneous modes (e.g., pressure support) (grey) for the 168 hours following enrollment. Ventilator mode was assessed approximately every 1 minute. Also shown is the number of patients who were alive and receiving invasive mechanical ventilation in each group during each time interval. Ventilator mode is reported for 561 of 566 patients on day 1. Two patients were missing data on ventilator mode on study day 1 but contributed data on subsequent days, and three patients were missing data on all days.

**Figure E5. Mean tidal volume**

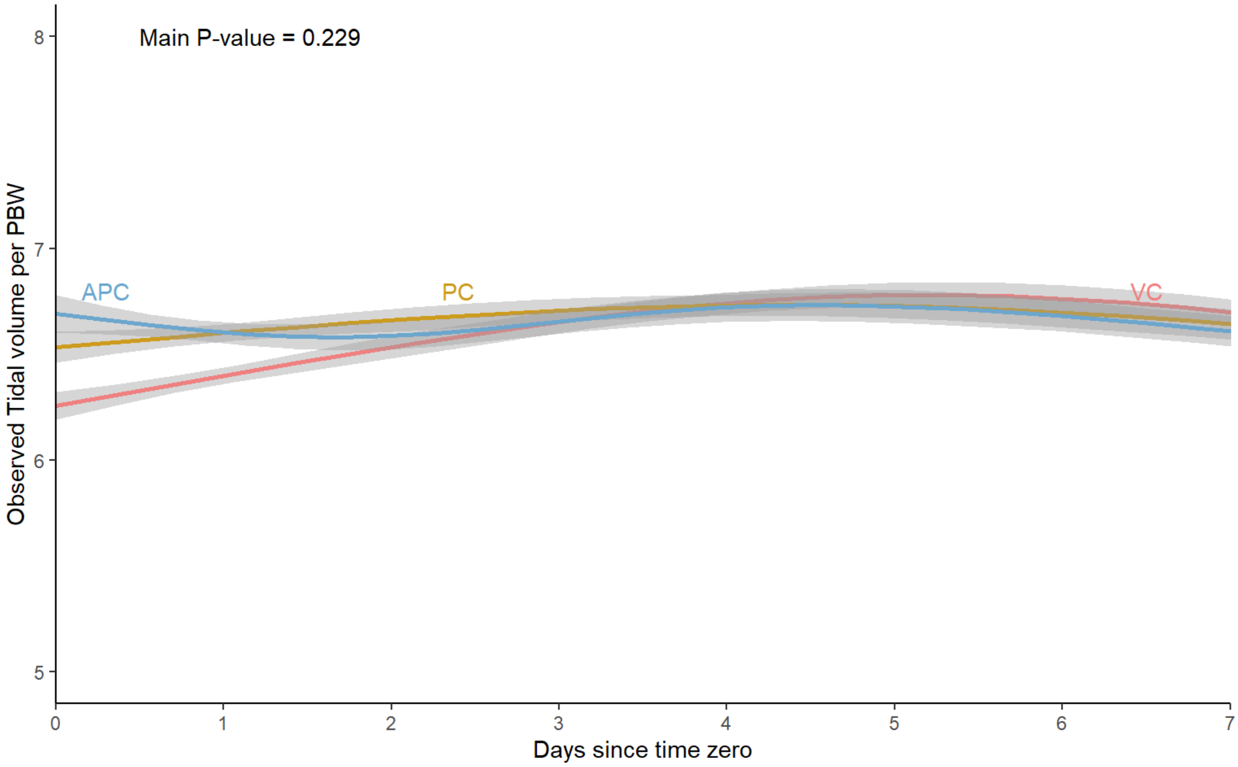

Tidal volume measurements adjusted for predicted body weight (PBW) are shown for the volume control group (red), the pressure control group (yellow), and the adaptive pressure control group (blue) over the first seven days after enrollment. All available datapoints for patients are displayed as mean (95% confidence interval) for each study group. Values on Day 0 represent the first available ventilator measurements after the time of enrollment. A p-value for the difference between groups was generated using a generalized estimating equation adjusting for within subject correlation, including independent variables of study group, study day, and a term for the interaction of group and study day. The number of patients with tidal volume measurements available declined from 565 on day 1 to 77 on day 7. Summary statistics of these values for patients in each study group on each study day are provided in Table E11.

**Figure E6. Proportion of tidal volumes above 8mL/kg predicted body weight**

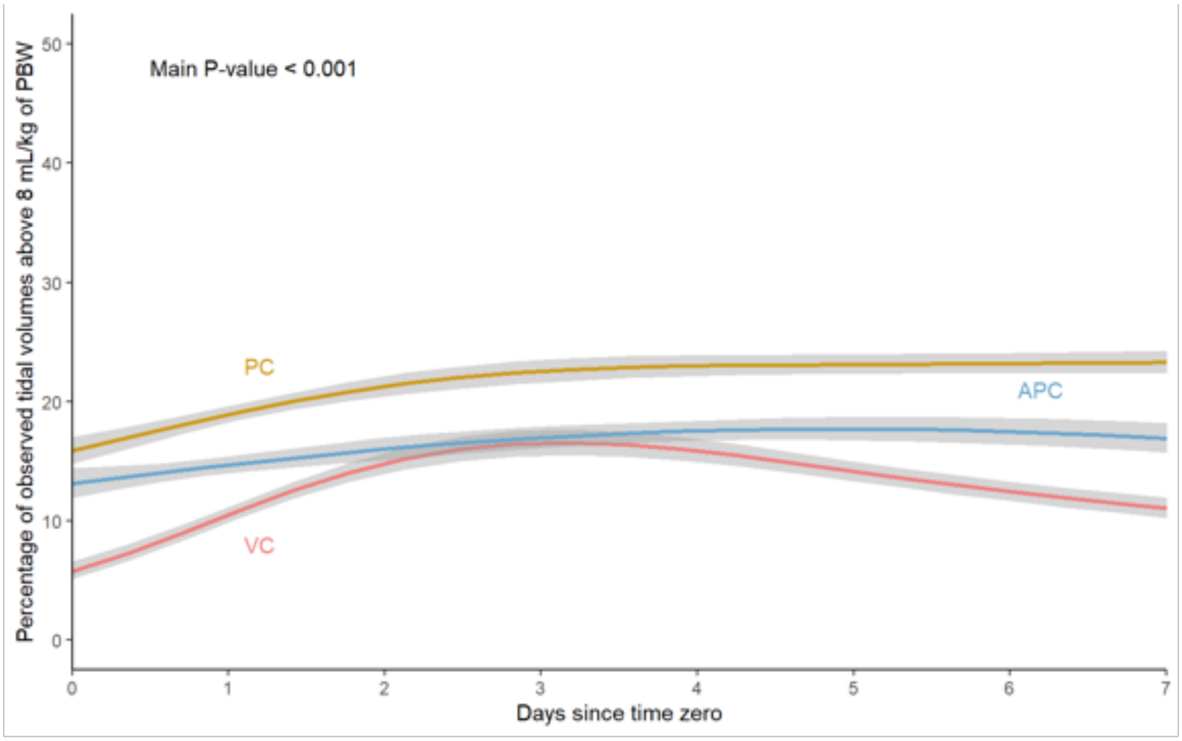

Percentages of tidal volume assessments above 8mL/kg of predicted body weight are shown for volume control group (red), pressure control group (yellow), and adaptive pressure control group (blue) over the first seven days after enrollment. All available datapoints for patients are displayed as mean proportion (95% confidence interval) for each study group. Values on Day 0 represent the first available ventilator measurements after the time of enrollment. A p-value for the difference between groups was generated using a generalized estimating equation adjusting for within subject correlation, including independent variables of study group, study day, and a term for the interaction of group and study day. The number of patients with tidal volume measurements available declined from 565 on day 1 to 77 on day 7. Summary statistics of these values for patients in each study group on each study day are provided in Table E11.

**Figure E7. Peak inspiratory pressure**

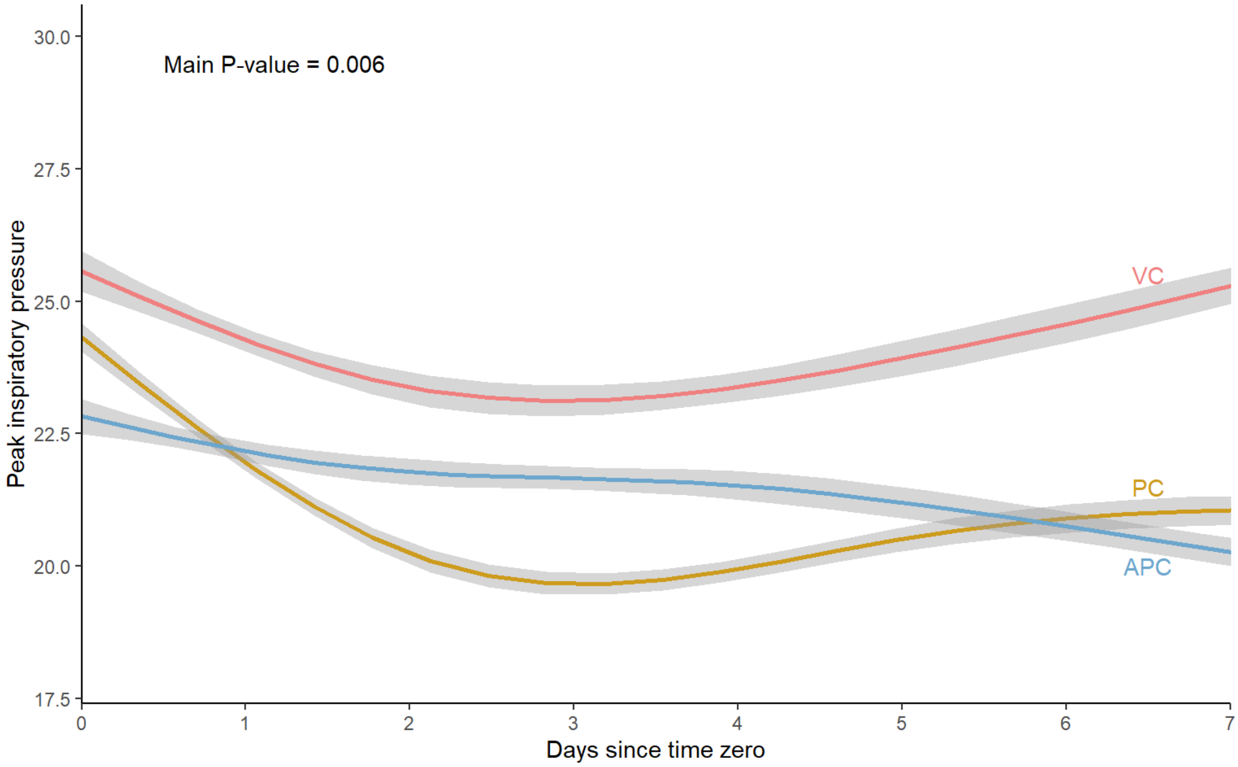

Peak inspiratory pressure measurements are shown for the volume control group (red), the pressure control group (yellow), and the adaptive pressure control group (blue) over the first seven days after enrollment. All available datapoints for patients are displayed as mean (95% confidence interval) for each study group. Values on Day 0 represent the first available ventilator measurements after the time of enrollment. A p-value for the difference between groups was generated using a generalized estimating equation adjusting for within subject correlation, including independent variables of study group, study day, and a term for the interaction of group and study day. The number of patients with peak inspiratory pressure measurements available declined from 565 on day 1 to 77 on day 7. Summary statistics of these values for patients in each study group on each study day are provided in the Table E11.

**Figure E8. Mean depth of sedation**

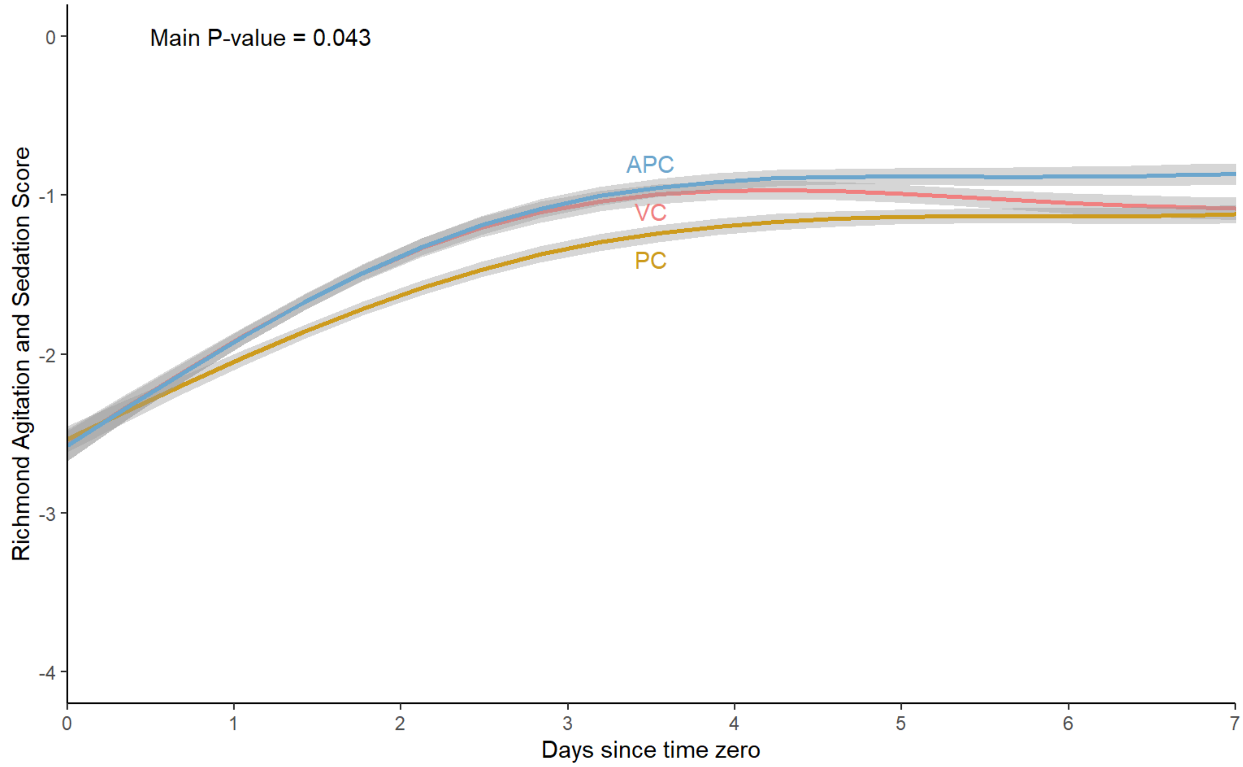

Depth of sedation assessments by the Richmond Agitation and Sedation Score are shown for the volume control group (red), the pressure control group (yellow), and the adaptive pressure control group (blue) over the first seven days after enrollment. All available datapoints for patients are displayed as mean (95% confidence interval) for each study group. Values on Day 0 represent the first available RASS assessments after the time of enrollment. A p-value for the difference between groups was generated using a generalized estimating equation adjusting for within subject correlation, including independent variables of study group, study day, and a term for the interaction of group and study day. The number of patients with RASS assessments available declined from 560 on day 1 to 302 on day 7. Summary statistics of these values for patients in each study group on each study day are provided in the Table E14.

**Figure E9. Proportion of assessments with coma**

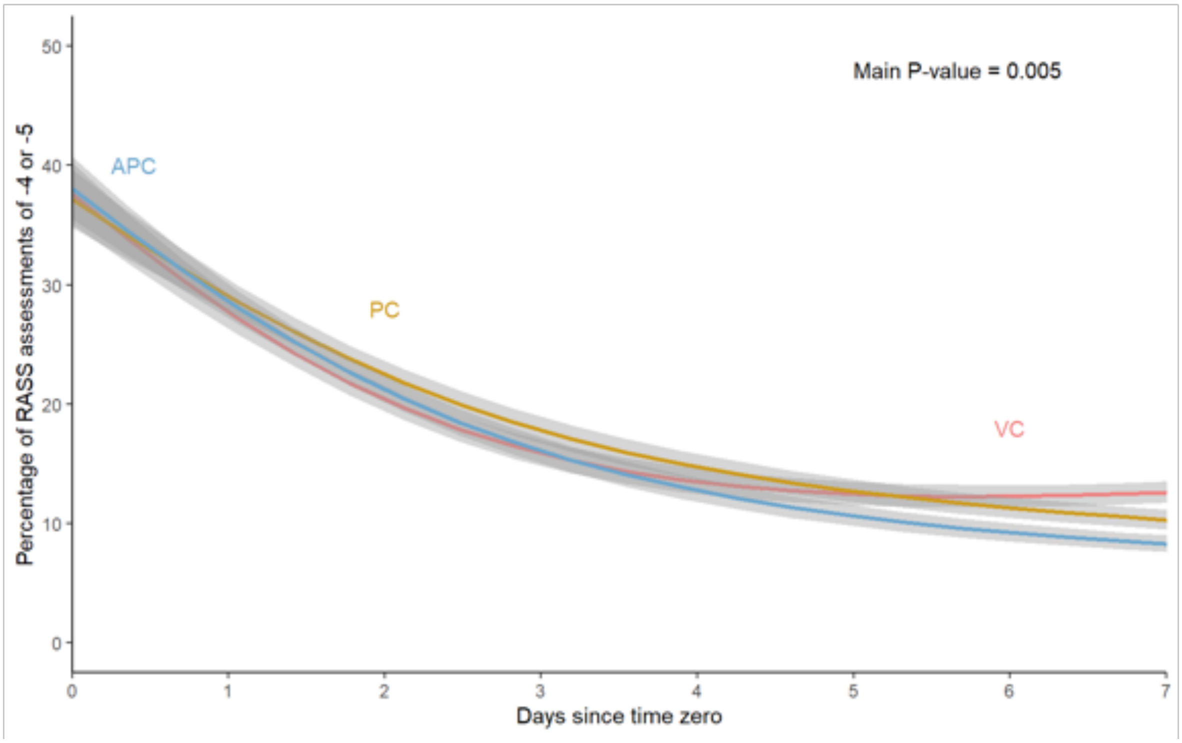

Percentage of assessments with coma defined by the Richmond Agitation and Sedation Score of -4 or -5 are shown for the volume control group (red), the pressure control group (yellow), and the adaptive pressure control group (blue) over the first seven days after enrollment. All available datapoints for patients are displayed as mean (95% confidence interval) for each study group. Values on Day 0 represent the first available assessments after the time of enrollment. A p-value for the difference between groups was generated using a generalized estimating equation adjusting for within subject correlation, including independent variables of study group, study day, and a term for the interaction of group and study day. The number of patients with RASS assessments available declined from 560 on day 1 to 302 on day 7. Summary statistics of these values for patients in each study group on each study day are provided in the Table E14.

**Figure E10. Number of blood gas tests per day**

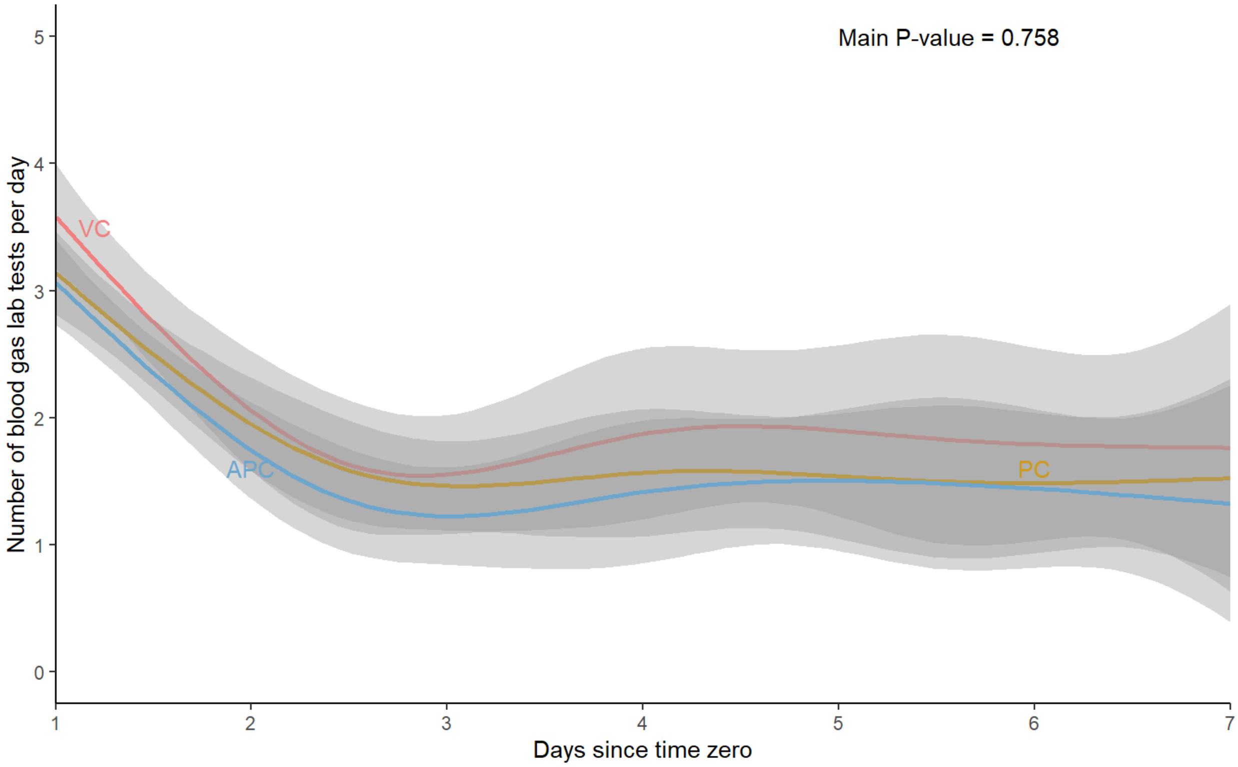

Number of blood gas tests obtained while on invasive mechanical ventilation per patient per day are shown for the volume control group (red), the pressure control group (yellow), and the adaptive pressure control group (blue) over the first seven days after enrollment. The number of venous or arterial blood gas tests were assessed for patients alive and receiving invasive mechanical ventilation. Values are displayed as mean (95% confidence interval) for each study group. A p-value for the difference between groups was generated using a generalized estimating equation adjusting for within subject correlation, including independent variables of study group, study day, and a term for the interaction of group and study day. The number of patients alive, receiving invasive mechanical ventilation and hospitalized declined from 566 on day 1 to 79 day 7. Summary statistics of these values for patients in each study group on each study day are provided in Table E10.

### Figure E11. Heterogeneity of treatment effect in the primary outcome

| Subgroup | No. of Patients | VC | PC | APC |
|---|---|---|---|---|
| | | median ventilator-free days [IQR] | | |
| FiO2 prior to enrollment | | | | |
| <0.3 | 73 | 24 [0 - 26] | 22.5 [0 - 24.5] | 23 [0 - 26.5] |
| 0.3-0.6 | 242 | 24 [0 - 26] | 22 [0 - 26] | 25 [2.5 - 26] |
| >0.6 | 201 | 5 [0 - 25] | 1 [0 - 26] | 0 [0 - 26] |
| SOFA Score at enrollment | | | | |
| <8 | 266 | 25 [11.5 - 26] | 23 [0 - 26] | 24.5 [0 - 26] |
| >=8 | 300 | 14 [0 - 25.75] | 19 [0 - 26] | 23 [0 - 26] |
| Chronic Obstructive Pulmonary Disease | | | | |
| Yes | 99 | 20 [0 - 26] | 24 [0 - 26] | 24 [0 - 26] |
| No | 467 | 24 [0 - 26] | 21 [0 - 26] | 23.5 [0 - 26] |
| Hypoxemic respiratory failure | | | | |
| Yes | 225 | 14 [0 - 24.5] | 0 [0 - 24] | 17 [0 - 26] |
| No | 341 | 25 [0 - 26] | 24 [0 - 26] | 25 [0 - 26] |
| Hypercarbic respriatory failure | | | | |
| Yes | 100 | 6 [0 - 25] | 1 [0 - 24] | 24 [13.75 - 26] |
| No | 466 | 24 [0 - 26] | 22 [0 - 26] | 23 [0 - 26] |
| Altered mental status | | | | |
| Yes | 329 | 23 [0 - 26] | 23 [0 - 26] | 24 [0 - 26] |
| No | 237 | 24 [0 - 26] | 18 [0 - 26] | 23 [0 - 26] |
| Duration of mechanical ventilation prior to enrollment | | | | |
| 0 minutes | 198 | 12 [0 - 25] | 0 [0 - 24] | 3.5 [0 - 25.25] |
| 1-360 minutes | 251 | 25 [0 - 26] | 24.5 [0 - 26] | 25 [0 - 26] |
| >360 minutes | 117 | 23 [0 - 26] | 23 [0 - 26] | 23 [0 - 26] |
| Receipt of vasopressors at enrollment | | | | |
| Yes | 214 | 0 [0 - 24] | 0 [0 - 24.25] | 5.5 [0 - 25] |
| No | 352 | 25 [13 - 26] | 24 [0 - 26] | 25 [0 - 26] |
| ARDS by the New Global Definition at enrollment | | | | |
| Yes | 210 | 0 [0 - 21] | 0 [0 - 22.5] | 0 [0 - 24] |
| No | 356 | 25 [14.5 - 26] | 25 [14 - 26] | 25 [2 - 26] |
| ***Overall*** | ***566*** | ***23 [0-26]*** | ***22 [0-26]*** | ***24 [0-26]*** |

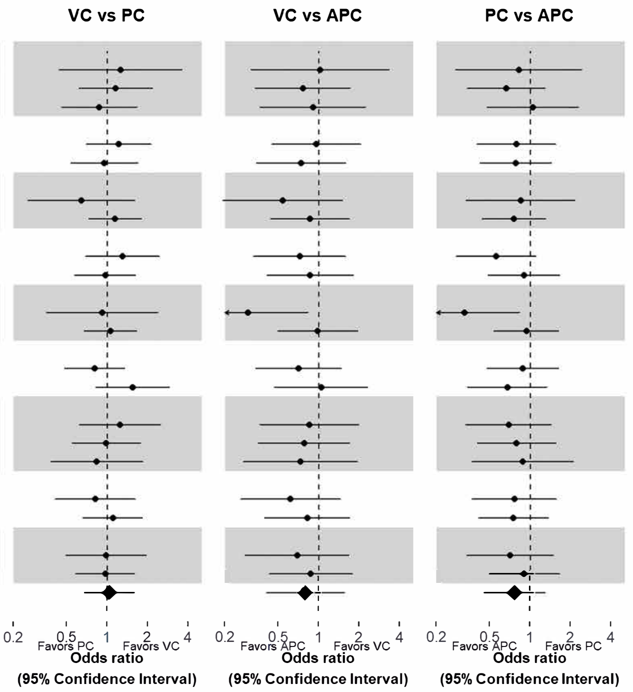

The median number of ventilator-free days through study day 28 (primary outcome) is compared among the three ventilator mode groups (volume control (VC), pressure control (PC), and adaptive pressure control (APC)) in subgroups of patients defined according to prespecified baseline characteristics. Odds ratios greater than 1.0 indicate a greater number of ventilator-free days (i.e., a better outcome). IQR denotes interquartile range. $FiO_2$ is the fraction of inspired oxygen, and SOFA score is the Sequential Organ Failure Assessment score.[48] Pre-enrollment $FiO_2$ was missing for 50 patients, such that in the volume control group, n=164; in the pressure control group n=182; and in the adaptive pressure control group, n=170. Acute respiratory distress syndrome (ARDS) criteria was defined using New Global Definition of ARDS[38] (See Supplemental Methods, Acute respiratory distress syndrome definition).

**Figure E12. Effect modification of the primary outcome by age**

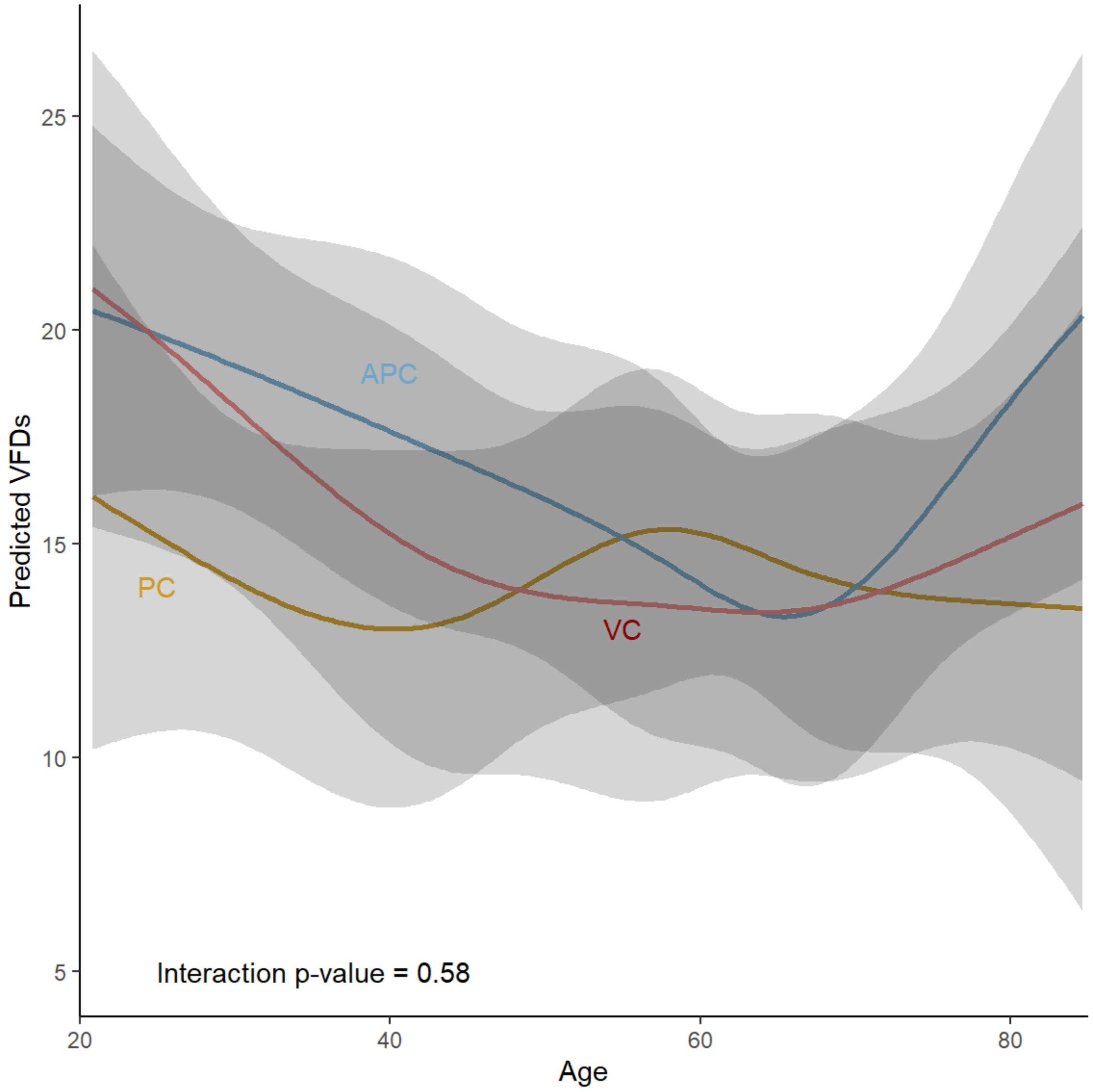

This figure displays the predicted number of ventilator-free days (VFDs, Y axis) in the volume control group (VC, red), pressure control group (PC, yellow), and adaptive pressure control (APC, blue) by the age of the patient, which ranged from 18 to 90 years. Patients' age did not modify the effect of ventilator mode group on ventilator-free days.

**Figure E13. Effect modification of the primary outcome by pre-enrollment $FiO_2$**

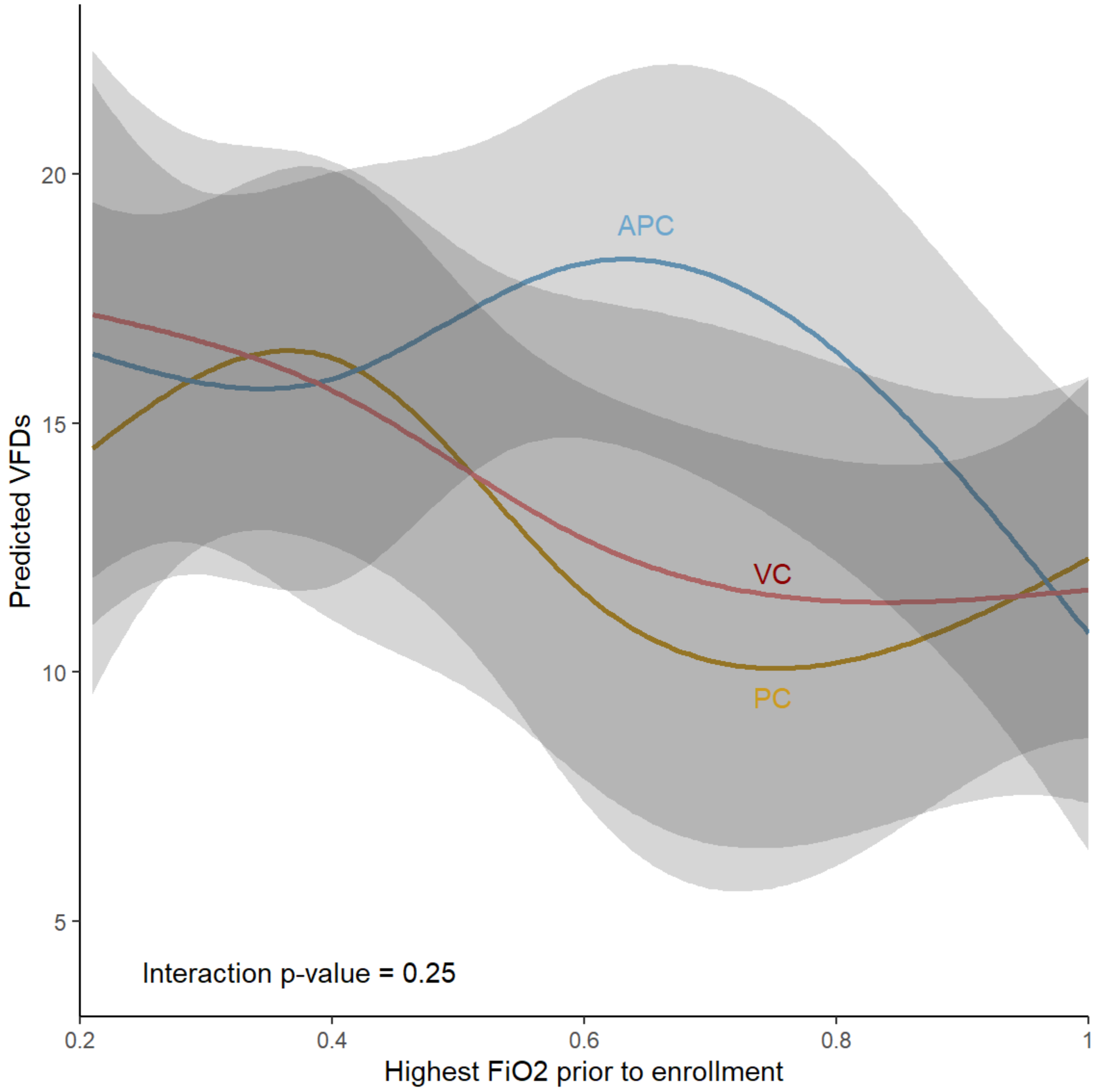

This figure displays the predicted number of ventilator-free days (VFDs, Y axis) in the volume control group (VC, red), pressure control group (PC, yellow), and adaptive pressure control (APC, blue) by the highest documented $FiO_2$ prior to enrollment, which ranged from 0.21 to 1. The $FiO_2$ received by patients prior to enrollment did not modify the effect of ventilator mode group on ventilator-free days.

**Figure E14. Effect modification of the primary outcome by baseline SOFA score**

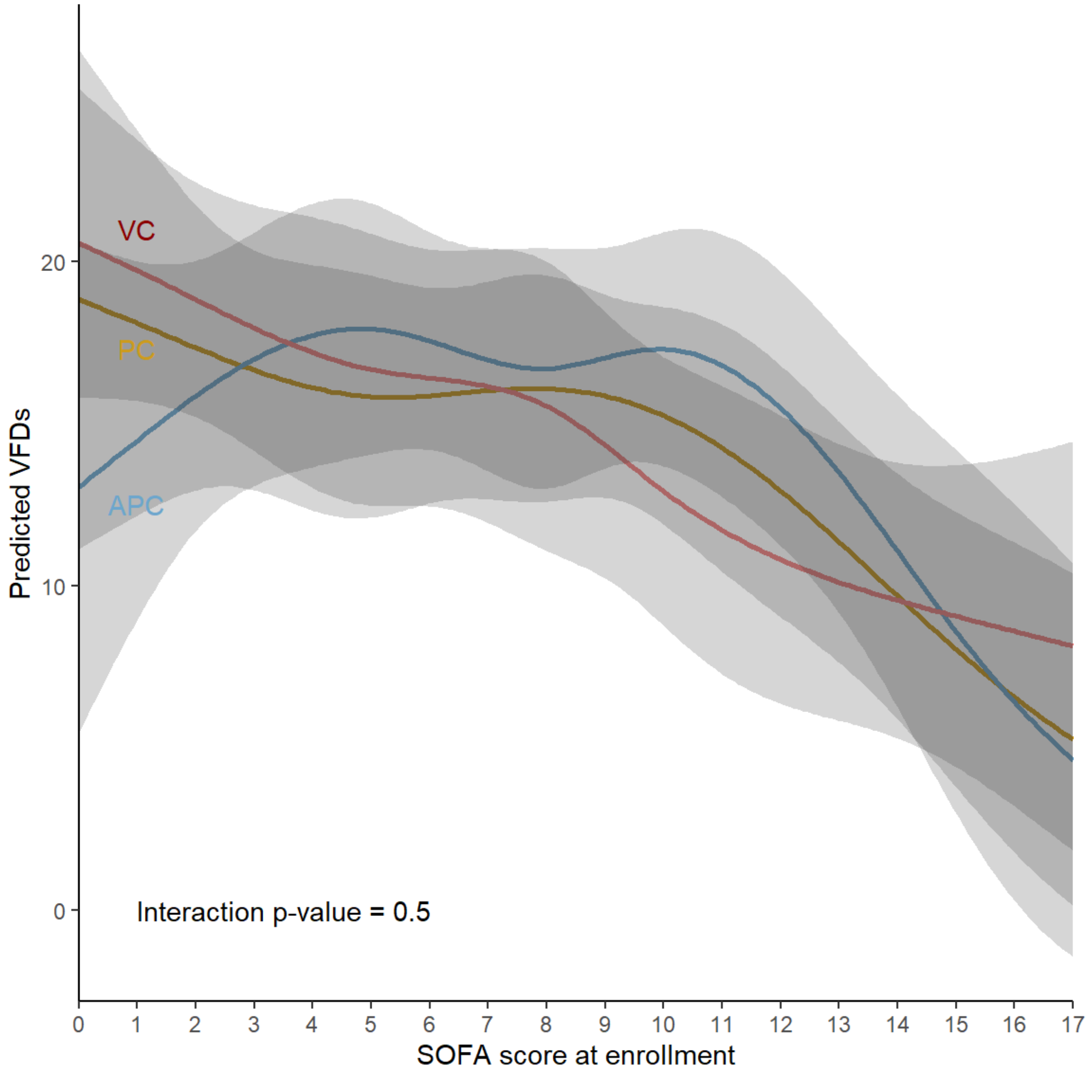

This figure displays the predicted number of ventilator-free days (VFDs, Y axis) in the volume control group (VC, red), pressure control group (PC, yellow), and adaptive pressure control (APC, blue) by patients' sequential organ failure assessment (SOFA) score at enrollment. The SOFA score is composed of scores from six organ systems, each graded on a scale of 0 (no organ dysfunction) to 4 (severe organ dysfunction or failure). Scores range from 0 (no evidence of organ dysfunction or failure in any organ system) to 24 (evidence of severe organ dysfunction or failure in each of the six organ systems assessed). Patients SOFA score at enrollment did not modify the effect of ventilator mode group on ventilator-free days.

**Figure E15. Mean daily SOFA score**

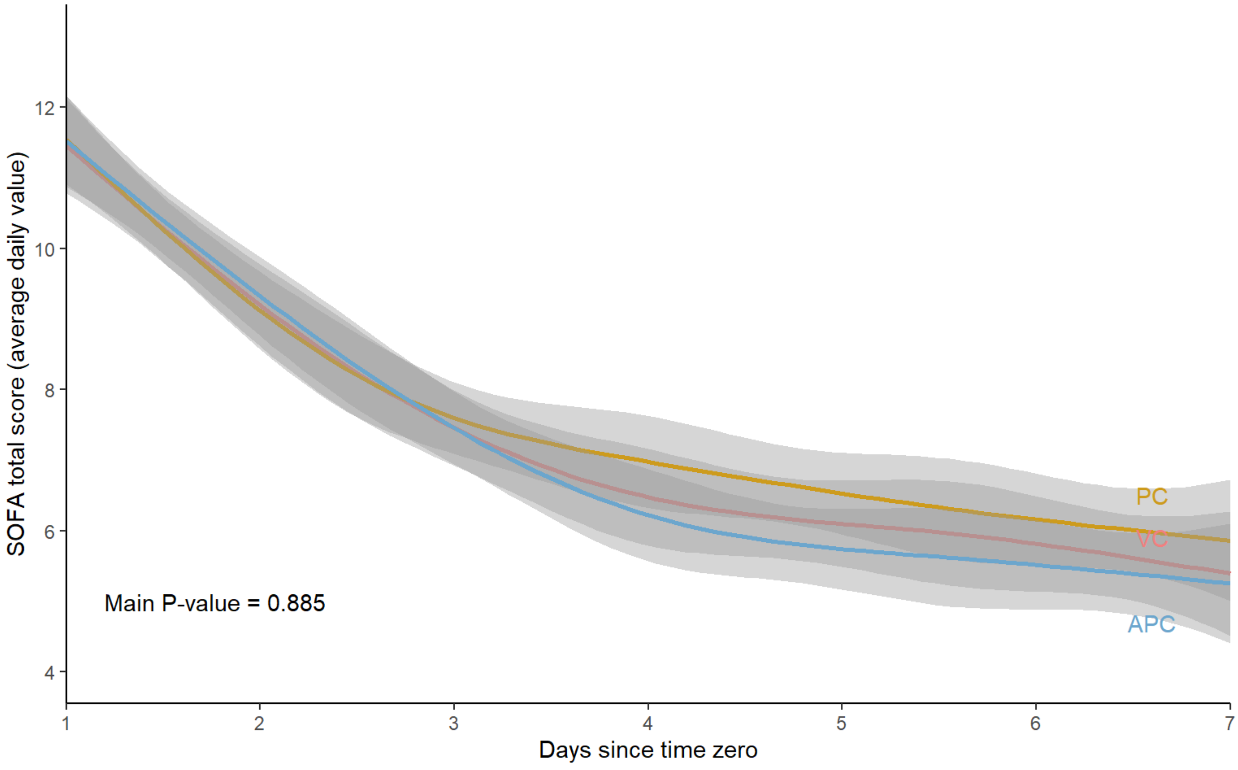

Severity of illness by the Sequential Organ Failure score[48] are shown for the volume control group (red), the pressure control group (yellow), and the adaptive pressure control group (blue) for the first seven days after enrollment. A daily average SOFA score was calculated for each patient alive and still hospitalized, and all of these values are displayed as mean (95% confidence interval) for each study group. Values on Day 0 represent the first available SOFA score assessments after the time of enrollment. A p-value for the difference between groups was generated using a generalized estimating equation adjusting for within subject correlation, including independent variables of study group, study day, and a term for the interaction of group and study day. The number of patients with SOFA assessments available declined from 566 at enrollment to 309 on day 7. Summary statistics of these values for patients in each study group on each study day are provided in Table E9.

### SUPPLEMENTAL TABLES

#### Table E1. Complete baseline demographics

| Characteristic | Volume Control (n=181) | Pressure Control (n=198) | Adaptive Pressure Control (n=187) |
|---|---|---|---|
| Age, median [IQR], years | 59.9 [46.3-68.8] | 56.9 [40.5-66.3] | 57.7 [39.7-67.3] |
| Female sex, no. (%) | 82 (45.3) | 81 (40.9) | 71 (38.0) |
| Race, No. (%)[a] | | | |
| American Indian or Alaska Native | 1 (0.6) | 1 (0.5) | 3 (1.6) |
| Asian | 3 (1.7) | 2 (1.0) | 3 (1.6) |
| Black or African American | 31 (17.1) | 28 (14.1) | 35 (18.7) |
| Native Hawaiian or Pacific Islander | 0 (0.0) | 0 (0.0) | 0 (0.0) |
| White | 138 (76.2) | 153 (77.3) | 133 (71.1) |
| Other or Unknown | 5 (2.8) | 7 (3.5) | 9 (4.8) |
| Ethnicity – Hispanic, no. (%)[b] | 7 (3.9) | 11 (5.9) | 8 (4.5) |
| Height, median [IQR], cm[c] | 172.7 [162.6-180.3] | 172.7 [165.1-180.3] | 172.7 [165.1-182.9] |
| Weight, median [IQR], kg[d] | 80.8 [68.0-100.0] | 80.9 [63.5-103.3] | 80.0 [68.0-98.7] |
| Hours from first receipt of mechanical ventilation to enrollment, median [IQR] | 2.3 [0.0-5.5] | 2.2 [0.0-5.4] | 1.8 [0.0-4.2] |
| Mechanical ventilation received for < 1 minute prior to enrollment, no. (%) | 59 (32.6) | 71 (35.9) | 68 (36.4) |
| Mechanical ventilation received for 1-360 minutes prior to enrollment, no. (%) | 84 (46.4) | 82 (41.4) | 85 (45.5) |
| Mechanical ventilation received for >360 minutes prior to enrollment, no. (%) | 38 (21.0) | 45 (22.7) | 34 (18.2) |
| Source of admission to study ICU, no. (%) | | | |
| ED at study hospital | 103 (56.9) | 114 (57.6) | 98 (52.4) |
| Hospital ward at study hospital | 37 (20.4) | 47 (23.7) | 45 (24.1) |
| Other ICU at study hospital | 10 (5.5) | 10 (5.1) | 7 (3.7) |
| OR or PACU at study hospital | 9 (5.0) | 7 (3.5) | 13 (7.0) |
| Outside hospital | 22 (12.2) | 20 (10.1) | 24 (12.8) |

*Definitions of abbreviations:* IQR=Interquartile range; ED=Emergency department; ICU= Intensive care unit; OR=Operating room; PACU= Post-anesthesia care unit

[a] Information on race was self-reported by patients using pre-specified categories and was missing for 22 patients

[b] Information on ethnicity was self-reported by patients and was missing for 26 patients.

[c] Information on height was not available for 60 patients.

[d] Information on weight was not available for 20 patients.

**Table E2. Acute illnesses**

| Characteristic | Volume Control (n=181) | Pressure Control (n=198) | Adaptive Pressure Control (n=187) |
|---|---|---|---|
| Acute illnesses, no. (%) | | | |
| Altered mental status | 120 (66.3) | 150 (75.8) | 136 (72.7) |
| Sepsis or septic shock[a] | 91 (50.3) | 115 (58.1) | 114 (61.0) |
| Acute respiratory distress syndrome[b] | 66 (36·5) | 83 (41·9) | 61 (32·6) |
| Acute kidney injury, stage II or greater[c] | 62 (34.3) | 72 (36.4) | 62 (33.2) |
| Pneumonia | 56 (30.9) | 72 (36.4) | 63 (33.7) |
| Aspiration pneumonitis or pneumonia | 38 (21.0) | 48 (24.2) | 38 (20.3) |
| Cardiac arrest | 20 (11.0) | 28 (14.1) | 27 (14.4) |
| Hepatic encephalopathy | 13 (7.2) | 24 (12.1) | 19 (10.2) |
| Chronic obstructive pulmonary disease exacerbation | 10 (5.5) | 7 (3.5) | 16 (8.6) |
| COVID-19 diagnosis | 11 (6.1) | 9 (4.5) | 5 (2.7) |
| Asthma exacerbation | 2 (1.1) | 0 (0.0) | 3 (1.6) |
| $FiO_2$ prior to enrollment, median [IQR][d] | 0.60 [0.40-1.00] | 0.60 [0.40-1.00] | 0.51 [0.36-0.95] |
| Receipt of vasopressors at enrollment, no. (%) | 68 (37.6) | 76 (38.4) | 70 (37.4) |
| SOFA score at enrollment, median [IQR][e] | 8 [5-11] | 8 [5-10] | 8 [5-10] |

*Definitions of abbreviations:* COVID-19= Coronavirus disease 2019; $FiO_2$= fraction of inspired oxygen; SOFA= Sequential Organ Failure score;

a Sepsis or septic shock is defined according to the Sepsis-3 criteria.[49]

b Acute respiratory distress syndrome (ARDS) criteria was defined using New Global Definition of ARDS[38] (See Supplemental Methods, Acute respiratory distress syndrome definition).

c Acute kidney injury, stage II or greater is defined according to Kidney Disease Improving Global Outcomes (KDIGO) creatinine criteria.[50] Acute kidney injury is presented for the 28 patients who were did not have End Stage Renal Disease requiring chronic renal replacement therapy at enrollment.

d Pre-enrollment $FiO_2$ was missing for 50 patients. In the volume control group, n=164; in the pressure control group n=182; and in the adaptive pressure control group, n=170.

e The Sequential Organ Failure Assessment (SOFA) score[48] is composed of scores from six organ systems, graded from 0 to 4 according to the degree of dysfunction or failure. Scores range from 0 (no evidence of organ dysfunction or failure) to 24 (evidence of severe organ dysfunction or failure).

**Table E3. Chronic comorbidities**

| Characteristic | Volume Control (n=181) | Pressure Control (n=198) | Adaptive Pressure Control (n=187) |
|---|---|---|---|
| Elixhauser comorbidity index, median [IQR] | 11 [3-17] | 10 [3-17] | 11 [3-18] |
| Charlson comorbidity index, median [IQR] | 3 [2-5] | 3 [2-5] | 3 [2-5] |
| Comorbidities, no. (%) | - | - | - |
| Coronary artery disease | 59 (32.6) | 39 (19.7) | 49 (26.2) |
| Chronic obstructive pulmonary disease | 35 (19.3) | 29 (14.6) | 35 (18.7) |
| End Stage Liver Disease or cirrhosis | 22 (12.2) | 35 (17.7) | 31 (16.6) |
| Congestive heart failure with reduced ejection fraction | 11 (6.1) | 12 (6.1) | 13 (7.0) |
| End-stage kidney disease on kidney replacement therapy | 11 (6.1) | 6 (3.0) | 11 (5.9) |
| Receipt of supplemental oxygen at place of residence prior to hospital admission, no. (%) | 25 (13.8) | 24 (12.1) | 22 (11.8) |

**Table E4. Indications for invasive mechanical ventilation**

| Indication[a] | Volume Control (n=181) | Pressure Control (n=198) | Adaptive Pressure Control (n=187) |
|---|---|---|---|
| Altered mental status, no. (%) | 101 (55.8) | 123 (62.1) | 105 (56.1) |
| Hypoxemic respiratory failure, no. (%) | 67 (37.0) | 81 (40.9) | 77 (41.2) |
| Hypercarbic respiratory failure, no. (%) | 32 (17.7) | 30 (15.2) | 38 (20.3) |
| Procedure, no. (%) | 26 (14.4) | 25 (12.6) | 30 (16.0) |
| Cardiac arrest, no. (%) | 18 (9.9) | 20 (10.1) | 26 (13.9) |
| Hemodynamic instability, no. (%) | 15 (8.3) | 19 (9.6) | 17 (9.1) |
| Seizure, no. (%) | 11 (6.1) | 18 (9.1) | 19 (10.2) |
| Acidosis, no. (%) | 15 (8.3) | 18 (9.1) | 15 (8.0) |
| Upper airway compromise, no. (%) | 22 (12.2) | 12 (6.1) | 13 (7.0) |
| Respiratory arrest, no. (%) | 3 (1.7) | 9 (4.5) | 11 (5.9) |
| Agitation, no. (%) | 3 (1.7) | 9 (4.5) | 3 (1.6) |

[a] Patients could have more than one indication.

**Table E5. Lab values at enrollment**

| Laboratory value[a] | Total N | Volume Control (n=181) | Pressure Control (n=198) | Adaptive Pressure Control (n=187) |
|---|---|---|---|---|
| Cell counts | | | | |
| White blood cells – $10^9$ per liter | 561 | 11.8 [7.4-17.8] | 12.2 [8.3-17.5] | 11.5 [8.1-16.4] |
| Hemoglobin – g/dL | 564 | 10.0 [8.6-12.6] | 10.5 [8.5-13.2] | 10.5 [8.5-12.9] |
| Platelets – $10^9$ per liter | 560 | 210 [109-279] | 202 [109-278] | 218 [120-296] |
| Metabolic panel | | | | |
| Sodium – mmol/L | 565 | 138 [134-141] | 138 [134-141] | 138 [133-141] |
| Potassium - mmol/L | 565 | 4.3 [3.7-4.8] | 4.1 [3.6-4.7] | 4.3 [3.8-4.8] |
| Bicarbonate – mmol/L | 564 | 22 [18-26] | 22 [19-27] | 22 [19-26] |
| Creatinine – mg/dL | 563 | 1.42 [0.84-2.74] | 1.29 [0.85-2.27] | 1.30 [0.82-2.86] |
| Lactate – mmol/L | 365 | 2.4 [1.4-5.0] | 2.4 [1.4-4.0] | 2.1 [1.4-4.4] |
| Liver function | | | | |
| Bilirubin – mg/dL | 473 | 0.7 [0.4-1.6] | 0.7 [0.4-1.9] | 0.8 [0.5-1.5] |
| Aspartate transaminase – IU | 474 | 50.5 [25.8-117.8] | 50.5 [26.0-108.8] | 37.5 [23.0-84.2] |
| Alanine aminotransferase – IU | 464 | 29.0 [18.0-81.0] | 31.0 [18.0-80.0] | 26.0 [16.0-52.0] |
| Arterial blood gas | | | | |
| pH | 241 | 7.28 [7.18-7.37] | 7.32 [7.19-7.40] | 7.32 [7.19-7.41] |
| $PaCO_2$ – mm Hg | 245 | 43.5 [36.0-50.2] | 39.0 [32.0-52.0] | 43.5 [34.8-53.2] |
| $PaO_2$ – mm Hg | 245 | 119.0 [78.5-178.5] | 107.0 [77.0-165.0] | 108.0 [83.8-197.5] |
| $SaO_2$ - % | 246 | 99 [95-100] | 98 [96-99] | 99 [96-100] |
| Venous blood gas | | | | |
| pH | 452 | 7.29 [7.19-7.35] | 7.30 [7.24-7.38] | 7.30 [7.22-7.36] |
| $PCO_2$ – mm Hg | 460 | 50.0 [42.0-60.5] | 47.0 [38.5-58.0] | 47.5 [39.2-59.0] |

[a] All values are presented as median and interquartile range. Trial protocol did not dictate laboratory measurements, all laboratory values represent those measured by treating clinicians as a part of clinical care.

**Table E6. First ventilator settings recorded after enrollment**

| Ventilator parameter[a] | n | Volume Control (n=181) | Pressure Control (n=198) | Adaptive Pressure Control (n=187) |
|---|---|---|---|---|
| Exhaled Tidal Volume, mL/kg predicted body weight | 565 | 6.1 [5.9-6.9] | 6.2 [5.7-7.3] | 6.1 [5.8-6.8] |
| Respiratory Rate (set), per minute | 557 | 20 [16-24] | 20 [16-26] | 20 [16-24] |
| Respiratory Rate (measured), per minute | 566 | 22 [18-26] | 22 [18-28] | 21 [18-26] |
| Peak Inspiratory Pressure, cm $H_2O$ | 565 | 24.4 [19.3-30.8] | 21.4 [17.1-27.6] | 22.0 [17.0-26.1] |
| Plateau Pressure, cm $H_2O$ | 358 | 19 [16-24] | 20 [15-24] | 19 [15-25] |
| Peak End-Expiratory Pressure, cm $H_2O$ | 556 | 5 [5-8] | 5 [5-8] | 5 [5-8] |
| Fraction inspired oxygen | 565 | 0.60 [0.40-0.99] | 0.59 [0.40-0.93] | 0.50 [0.40-0.98] |

[a] All values are presented as median and interquartile range.

**Table E7. Ventilator mode assessments**

| Ventilator parameter | N | Volume Control | Pressure Control | Adaptive Pressure Control |
|---|---|---|---|---|
| Mode, no. (%) | 1,405,218 | | | |
| Day 1 | 385,437 | n=115,755 | n=140,230 | n=129,452 |
| Volume Control | | 92,656 (80.0) | 5,466 (3.9) | 2,063 (1.6) |
| Pressure Control | | 2,734 (2.4) | 111,693 (79.6) | 905 (0.7) |
| Adaptive Pressure Control | | 2,876 (2.5) | 3,561 (2.5) | 110,885 (85.7) |
| SIMV | | 712 (0.6) | 118 (0.1) | 289 (0.2) |
| Pressure support | | 16,777 (14.5) | 19,392 (13.8) | 15,310 (11.8) |
| Other | | 0 (0.0) | 0 (0.0) | 0 (0.0) |
| Day 2 | 225,863 | n=69,822 | n=84,483 | n=71,558 |
| Volume Control | | 51,201 (73.3) | 3,271 (3.9) | 1,499 (2.1) |
| Pressure Control | | 1,644 (2.4) | 63,967 (75.7) | 83 (0.1) |
| Adaptive Pressure Control | | 2,308 (3.3) | 1,000 (1.2) | 52,879 (73.9) |
| SIMV | | 190 (0.3) | 48 (0.1) | 201 (0.3) |
| Pressure support | | 14,479 (20.7) | 16,197 (19.2) | 16,896 (23.6) |
| Other | | 0 (0.0) | 0 (0.0) | 0 (0.0) |
| Day 3 | 158,722 | n=50,781 | n=64,356 | n=43,585 |
| Volume Control | | 33,427 (65.8) | 1,975 (3.1) | 489 (1.1) |
| Pressure Control | | 1,501 (3.0) | 46,108 (71.6) | 174 (0.4) |
| Adaptive Pressure Control | | 2,268 (4.5) | 1,596 (2.5) | 32,568 (74.7) |
| SIMV | | 84 (0.2) | 114 (0.2) | 0 (0.0) |
| Pressure support | | 13,501 (26.6) | 14,563 (22.6) | 10,354 (23.8) |
| Other | | 0 (0.0) | 0 (0.0) | 0 (0.0) |
| Day 4 | 114,676 | n=35,430 | n=47,537 | n=31,709 |
| Volume Control | | 25,195 (71.1) | 1,285 (2.7) | 75 (0.2) |
| Pressure Control | | 1,855 (5.2) | 35,460 (74.6) | 662 (2.1) |
| Adaptive Pressure Control | | 2,672 (7.5) | 1,100 (2.3) | 21,330 (67.3) |
| SIMV | | 4 (0.0) | 0 (0.0) | 0 (0.0) |
| Pressure support | | 5,704 (16.1) | 9,692 (20.4) | 9,642 (30.4) |
| Other | | 0 (0.0) | 0 (0.0) | 0 (0.0) |
| Day 5 | 96,710 | n=31,657 | n=40,187 | n=24,866 |
| Volume Control | | 20,072 (63.4) | 730 (1.8) | 219 (0.9) |
| Pressure Control | | 1,568 (5.0) | 30,744 (76.5) | 927 (3.7) |

| | | | | |
|---|---|---|---|---|
| Adaptive Pressure Control | | 3,347 (10.6) | 917 (2.3) | 15,950 (64.1) |
| SIMV | | 1 (0.0) | 11 (0.0) | 2 (0.0) |
| Pressure support | | 6,669 (21.1) | 7,785 (19.4) | 7,768 (31.2) |
| Other | | 0 (0.0) | 0 (0.0) | 0 (0.0) |
| Day 6 | 77,103 | n=25,038 | n=31,033 | n=21,032 |
| Volume Control | | 12,540 (50.1) | 1,872 (6.0) | 113 (0.5) |
| Pressure Control | | 2,058 (8.2) | 22,581 (72.8) | 691 (3.3) |
| Adaptive Pressure Control | | 4,235 (16.9) | 916 (3.0) | 14,324 (68.1) |
| SIMV | | 0 (0.0) | 29 (0.1) | 0 (0.0) |
| Pressure support | | 6,205 (24.8) | 5,635 (18.2) | 5,904 (28.1) |
| Other | | 0 (0.0) | 0 (0.0) | 0 (0.0) |
| Day 7 | 58,319 | n=18,720 | n=24,213 | n=15,386 |
| Volume Control | | 10,257 (54.8) | 1,741 (7.2) | 0 (0.0) |
| Pressure Control | | 2,044 (10.9) | 15,309 (63.2) | 630 (4.1) |
| Adaptive Pressure Control | | 3,600 (19.2) | 1,520 (6.3) | 10,150 (66.0) |
| SIMV | | 0 (0.0) | 20 (0.1) | 84 (0.5) |
| Pressure support | | 2,819 (15.1) | 5,623 (23.2) | 4,522 (29.4) |
| Other | | 0 (0.0) | 0 (0.0) | 0 (0.0 |

**Table E8. Time from enrollment to documentation of any ventilator mode, of the assigned ventilator mode, and of pressure support**

| Time interval[a] | n | Volume Control (n=181) | Pressure Control (n=198) | Adaptive Pressure Control (n=187) |
|---|---|---|---|---|
| Time from enrollment to first documentation of any ventilator mode, minutes | 564 | 4.2 [0.8-13.8] | 3.9 [0.7-10.6] | 2.9 [0.7-11.5] |
| Time from enrollment to first documentation of the assigned ventilator mode, minutes | 547 | 5.2 [1.7-16.6] | 5.2 [1.4-17.1] | 4.6 [0.8-15.1] |
| Time from enrollment to first documentation of pressure support mode, hours | 444 | 14.7 [5.0-32.5] | 13.1 [6.2-26.1] | 14.5 [6.4-23.6] |

a All values are presented as median and interquartile range.

**Table E9. Compliance with group assignment and separation between groups**

| Outcome | n | Volume Control N=181 | Pressure Control N=198 | Adaptive Pressure Control N=187 | VC vs PC Median or Risk Difference (95% CI) | VC vs APC Median or Risk Difference (95% CI) | PC vs APC Median or Risk Difference (95% CI) |
|---|---|---|---|---|---|---|---|
| Percentage of time on a mandatory mode of ventilation in the first 72 hours receiving the assigned mode, median [IQR] % | 550 | 100.0 [100.0-100.0] | 100.0 [98.9-100.0] | 100.0 [100.0-100.0] | 0.0 (0.0 to 0.0) | 0.0 (0.0 to 0.0) | 0.0 (0.0 to 0.0) |
| Number (%) of patients for whom >80% of breaths in a mandatory mode in the first 72 hours were in the assigned mode | 550 | 162 (92.0) | 172 (88.7) | 175 (97.2) | 3.4 (-3.1 to 9.9) | -5.2 (-10.4 to 0.0) | -8.6 (-14.2 to -3.0) |
| Number (%) of patients for whom 100% of breaths in a mandatory mode in the first 72 hours were in the assigned mode | 550 | 147 (83.5) | 129 (66.5) | 145 (80.6) | 17.0 (7.9 to 26.2) | 3.0 (-5.6 to 11.5) | -14.1 (-23.4 to -4.7) |
| Percentage of time on mechanical ventilation in the first 72 hours receiving the assigned mode (including time on a spontaneous mode), median [IQR] % | 563 | 87.5 [61.0-98.4] | 90.2 [56.6-98.9] | 93.8 [74.2-99.7] | -2.7 (-8.7 to 4.2) | -6.3 (-11.7 to -0.7) | -3.6 (-8.5 to 0.6) |
| Receipt of a mode modification sheet completed by treating clinicians, n (%) | 566 | 19 (10.5) | 25 (12.6) | 4 (2.1) | -2.1 (-9.1 to 4.8) | 8.4 (2.9 to 13.8) | 10.5 (4.9 to 16.1) |
| Number (%) of patients who experienced a crossover (i.e., received mechanical ventilation into a subsequent study month) | 566 | 22 (12.2) | 23 (11.6) | 16 (8.6 | | | |
| Calendar days receiving mechanical ventilation prior to crossover, median [IQR] days | 61 | 9 [5-12] | 11 [5-16] | 7 [4-15] | | | |
| Calendar days receiving mechanical ventilation after crossover, median [IQR] days | 61 | 6 [3-9] | 10 [4-12] | 4 [2-8] | | | |

**Table E10. Modifications of the mode: number, timing, and rationale**

| | Volume Control N=181 | Pressure Control N=198 | Adaptive Pressure Control N=187 |
|---|---|---|---|
| Mode modified, No. (%) | 19 (10.5) | 25 (12.6) | 4 (2.1) |
| Time from enrollment to modification, median [IQR], days | 0.8 [0.1-4.2] | 0.5 [0.1-1.5] | 0.4 [0.1-1.2] |
| Rationale for modifying ventilator mode provided by clinician, No. (%)[a] | | | |
| Asynchrony or dyssynchrony not amenable to changes within the assigned mode | 8 (4.4) | 7 (3.5) | 1 (0.5) |
| Peak pressures persistently high (e.g. > 40 cmH2O) | 13 (7.2) | 2 (1.0) | 1 (0.5) |
| Need to increase minute ventilation in acidosis | 1 (0.6) | 12 (6.1) | 2 (1.1) |
| Excessive work of breathing | 4 (2.2) | 2 (1.0) | 2 (1.1) |
| Refractory hypoxemia | 4 (2.2) | 3 (1.5) | 0 (0) |
| Inability to limit tidal volumes delivered | 2 (1.1) | 3 (1.5) | 0 (0) |
| Need to guarantee minute ventilation in variable physiology | 0 (0) | 3 (1.5) | 1 (0.5) |
| Need to increase tidal volume in low compliance respiratory system | 0 (0) | 4 (2.0) | 0 (0) |
| Clinician preference, during extra corporeal membrane oxygenation | 3 (1.7) | 0 (0) | 0 (0) |
| Clinician preference, not otherwise specified | 1 (0.6) | 2 (1.0) | 0 (0) |
| Intrinsic PEEP | 2 (1.1) | 0 (0) | 1 (0.5) |
| Barotrauma (pneumothorax or pneumomediastinum) | 0 (0) | 1 (0.5) | 0 (0) |
| Need to limit inspiratory pressure | 0 (0) | 1 (0.5) | 0 (0) |
| Transition to home ventilator settings | 0 (0) | 1 (0.5) | 0 (0) |
| Patient preference | 1 (0.6) | 0 (0) | 0 (0) |

a Clinicians provided the rationale for modifying the ventilator mode for all 48 MODE modification sheets. Clinicians could provide one or more rationales using either pre-specified categories or free text. In 31 cases, at least one reason was adjudicated by the research team from free-text responses.

**Table E11. Daily exploratory outcomes**

| | n | Volume Control N=181 | Pressure Control N=198 | Adaptive Pressure Control N=187 | VC vs PC Difference (95% CI) | VC vs APC Difference (95% CI) | PC vs APC Difference (95% CI) |
|---|---|---|---|---|---|---|---|
| Mean exhaled tidal volume, median [IQR], mL[a] | | | | | | | |
| Day 1[b] | 565 | 414 [354-463] | 423 [374-477] | 436 [386-476] | | | |
| Day 2 | 328 | 399 [353-454] | 405 [349-460] | 448 [395-484] | | | |
| Day 3 | 216 | 408 [356-451] | 408 [348-471] | 453 [414-489] | | | |
| Day 4 | 155 | 406 [378-439] | 417 [370-477] | 470 [418-505] | | | |
| Day 5 | 123 | 400 [373-465] | 439 [350-492] | 453 [409-495] | | | |
| Day 6 | 99 | 397 [373-454] | 436 [347-518] | 440 [389-485] | | | |
| Day 7 | 77 | 401 [356-453] | 418 [335-520] | 426 [402-496] | | | |
| Mean exhaled tidal volume, median [IQR], mL/kg PBW[a,c] | | | | | | | |
| Day 1 | 565 | 6.2 [5.9-7.1] | 6.4 [5.8-7.4] | 6.2 [5.9-6.9] | -0.2 (-0.5 to 0.1) | 0.0 (-0.3 to 0.2) | 0.2 (-0.1 to 0.4) |
| Day 2 | 328 | 6.3 [5.8-7.3] | 6.3 [5.5-7.4] | 6.3 [5.9-7.4] | 0.1 (-0.4 to 0.7) | 0.1 (-0.2 to 0.5) | 0.0 (-0.4 to 0.4) |
| Day 3 | 216 | 6.2 [5.7-7.1] | 6.3 [5.6-7.4] | 6.4 [5.9-7.1] | -0.1 (-0.6 to 0.4) | -0.1 (-0.4 to 0.3) | -0.1 (-0.5 to 0.5) |
| Day 4 | 155 | 6.2 [5.8-7.2] | 6.5 [5.7-7.8] | 6.4 [5.9-7.8] | -0.3 (-1.0 to 0.5) | -0.2 (-0.8 to 0.5) | 0.1 (-0.7 to 0.8) |
| Day 5 | 123 | 6.5 [5.7-7.5] | 6.5 [5.5-7.8] | 6.6 [6.0-7.5] | 0.0 (-1.2 to 0.6) | -0.1 (-0.7 to 0.6) | -0.1 (-0.4 to 1.1) |
| Day 6 | 99 | 6.3 [5.7-7.1] | 6.3 [5.7-7.4] | 6.4 [5.9-7.1] | 0.0 (-0.8 to 0.8) | -0.1 (-0.7 to 0.8) | -0.1 (-0.8 to 0.8) |
| Day 7 | 77 | 6.3 [5.7-7.3] | 6.7 [5.4-7.2] | 6.6 [5.5-6.9] | -0.4 (-0.9 to 0.8) | -0.2 (-0.8 to 1.2) | 0.1 (-0.7 to 1.2) |
| Percentage of exhaled tidal volumes > 8mL/kg PBW, median [IQR][a] | | | | | | | |

| | | | | | | | |
|---|---|---|---|---|---|---|---|
| Day 1 | 565 | 0.0 [0.0-8.3] | 6.8 [0.0-31.4] | 3.6 [0.0-14.3] | -6.8 (-11.1 to -0.8) | -3.6 (-5.6 to 0.1) | 3.2 (-2.1 to 8.5) |
| Day 2 | 328 | 3.3 [0.0-17.9] | 3.2 [0.0-23.8] | 3.6 [0.0-29.1] | 0.1 (-5.7 to 5.3) | -0.2 (-5.1 to 3.6) | -0.3 (-5.1 to 5.5) |
| Day 3 | 216 | 3.6 [0.0-13.6] | 5.4 [0.0-32.8] | 3.4 [0.0-19.7] | -1.7 (-13.6 to 2.9) | 0.3 (-6.4 to 5.9) | 2.0 (-5.3 to 13.9) |
| Day 4 | 155 | 1.7 [0.0-17.3] | 10.3 [0.0-46.2] | 8.3 [0.0-26.5] | -8.6 (-20.8 to 2.6) | -6.6 (-15.6 to 2.4) | 2.0 (-9.2 to 14.5) |
| Day 5 | 123 | 6.9 [0.0-19.9] | 6.7 [0.0-48.1] | 7.0 [0.0-27.5] | 0.2 (-28.0 to 8.1) | -0.1 (-7.1 to 8.0) | -0.4 (-6.4 to 32.2) |
| Day 6 | 99 | 3.7 [0.0-21.1] | 3.8 [0.0-28.8] | 6.2 [0.0-19.5] | -0.1 (-11.9 to 10.0) | -2.5 (-12.5 to 8.9) | -2.4 (-11.9 to 10.5) |
| Day 7 | 77 | 3.6 [0.0-9.4] | 11.5 [0.0-24.1] | 4.0 [0.0-24.9] | -8.0 (-18.8 to 3.4) | -0.4 (-8.3 to 7.1) | 7.5 (-5.9 to 18.8) |
| Mean peak airway pressure, median [IQR], cm $H_2O$[a] | | | | | | | |
| Day 1 | 565 | 22.4 [18.6-28.6] | 20.6 [15.9-26.4] | 20.3 [16.5-25.0] | 1.8 (-0.7 to 3.9) | 2.1 (0.4 to 3.9) | 0.3 (-1.8 to 2.8) |
| Day 2 | 328 | 20.8 [16.7-27.9] | 19.9 [15.0-24.5] | 19.8 [15.1-24.9] | 0.9 (-0.9 to 4.1) | 1.0 (-1.6 to 3.5) | 0.1 (-3.6 to 2.0) |
| Day 3 | 216 | 21.6 [17.1-29.8] | 19.9 [14.6-24.4] | 19.8 [14.8-27.4] | 1.7 (-1.0 to 7.0) | 1.7 (-1.8 to 5.9) | 0.1 (-4.5 to 1.9) |
| Day 4 | 155 | 23.0 [15.9-28.9] | 20.1 [15.4-26.2] | 18.4 [14.7-26.9] | 2.8 (-2.6 to 6.1) | 4.5 (-1.6 to 7.4) | 1.7 (-2.6 to 5.1) |
| Day 5 | 123 | 23.7 [15.6-30.0] | 19.0 [16.3-25.0] | 20.2 [14.4-25.0] | 4.7 (-1.9 to 8.4) | 3.5 (-2.1 to 8.8) | -1.2 (-4.4 to 5.0) |
| Day 6 | 99 | 24.2 [19.6-26.7] | 19.3 [15.0-26.5] | 19.7 [16.9-25.9] | 4.9 (0.6 to 7.9) | 4.5 (-0.4 to 7.4) | -0.4 (-4.9 to 3.3) |
| Day 7 | 77 | 23.8 [17.4-26.5] | 19.1 [16.9-24.1] | 19.1 [12.9-23.3] | 4.7 (-2.6 to 7.9) | 4.6 (-3.0 to 9.5) | 0.0 (-3.9 to 5.9) |
| Blood gas laboratory tests performed, median [IQR][d] | | | | | | | |
| Day 1 | 566 | 3 [1-5] | 2 [1-5] | 3 [1-5] | 1 (-1 to 1) | 0 (-1 to 1) | -1 (-1 to 1) |

| | | | | | | | |
|---|---|---|---|---|---|---|---|
| Day 2 | 333 | 1 [0-3] | 1 [0-3] | 1 [0-2] | 0 (0 to 1) | 0 (0 to 1) | 0 (0 to 0) |
| Day 3 | 224 | 1 [0-3] | 1 [0-3] | 0 [0-2] | 0 (-1 to 1) | 1 (0 to 1) | 1 (-1 to 2) |
| Day 4 | 163 | 0 [0-3] | 1 [0-2] | 1 [0-2] | -1 (-1 to 1) | -1 (-1 to 1) | 0 (-1 to 1) |
| Day 5 | 127 | 1 [0-4] | 1 [0-2] | 0 [0-2] | 0 (-1 to 2) | 1 (-1 to 2) | 1 (-1 to 1) |
| Day 6 | 102 | 1 [0-3] | 1 [0-2] | 1 [0-2] | 0 (-1 to 1) | 0 (-1 to 1) | 0 (-1 to 1) |
| Day 7 | 79 | 1 [0-2] | 1 [0-2] | 0 [0-1] | 0 (-1 to 2) | 1 (-1 to 2) | 1 (-1 to 1) |
| Mean daily SOFA Score median [IQR][e] | | | | | | | |
| Day 1 | 562 | 11.0 [8.4, 13.9] | 11.9 [8.5, 14.0] | 11.0 [8.4, 13.9] | -0.9 (-1.8 to 0.5) | 0.0 (-1.2 to 1.2) | 0.9 (-0.8 to 1.9) |
| Day 2 | 505 | 8.9 [6.0, 12.5] | 8.6 [5.9, 12.2] | 9.0 [5.8, 13.5] | 0.2 (-1.1 to 1.0) | -0.1 (-1.5 to 1.5) | -0.4 (-1.4 to 1.2) |
| Day 3 | 460 | 6.2 [3.0, 10.2] | 7.0 [3.8, 10.8] | 6.5 [3.6, 10.8] | -0.8 (-2.2 to 0.5) | -0.2 (-1.8 to 1.0) | 0.5 (-1.0 to 1.9) |
| Day 4 | 415 | 5.2 [2.5, 10.5] | 5.8 [2.8, 10.0] | 5.0 [2.5, 9.0] | -0.5 (-2.8 to 1.0) | 0.2 (-1.5 to 2.0) | 0.8 (-0.8 to 3.0) |
| Day 5 | 380 | 4.8 [2.0, 9.2] | 5.8 [2.5, 9.2] | 4.4 [2.1, 7.8] | -1.0 (-2.8 to 0.8) | 0.4 (-1.0 to 1.9) | 1.4 (-0.1 to 2.8) |
| Day 6 | 340 | 4.0 [2.0, 8.5] | 4.8 [2.9, 8.4] | 4.5 [2.0, 7.4] | -0.8 (-2.2 to 1.5) | -0.5 (-1.2 to 1.8) | 0.2 (-0.8 to 2.2) |
| Day 7 | 309 | 4.0 [2.0, 8.0] | 4.8 [2.5, 7.8] | 4.1 [2.0, 7.0] | -0.8 (-2.0 to 0.5) | -0.1 (-1.2 to 1.2) | 0.6 (-0.2 to 2.0) |

*Definitions of abbreviations:* VC=volume control, PC=pressure control, APC=adaptive pressure control, IQR=interquartile range, PBW=predicted body weight

a Tidal volumes, peak airway pressures, and arterial blood gases are assessed from time of enrollment to first extubation and reported as a mean daily value.

b Study days are sequential 24-hour intervals from time zero. For example, day 1 is the time of enrollment to 24 hours after the time of enrollment.

c Height was missing in 60 patients and was imputed using all available baseline data for calculation of PBW.

d Blood gas tests refer to both arterial and venous blood gas testing

e SOFA score reported as the first SOFA score on each study day.

**Table E12. Coma, delirium, and sedation on study days 1 to 7**

| | n | Volume Control N=181 | Pressure Control N=198 | Adaptive Pressure Control N=187 | VC vs PC Difference (95% CI) | VC vs APC Difference (95% CI) | PC vs APC Difference (95% CI) |
|---|---|---|---|---|---|---|---|
| Mean RASS - median [IQR] | | | | | | | |
| Day 1 | 560 | -2.3 [-3.5 to -1.2] | -2.5 [-3.8 to -1.2] | -2.2 [-3.6 to -1.1] | 0.2 (-0.4 to 0.7) | -0.0 (-0.6 to 0.5) | -0.3 (-0.7 to 0.4) |
| Day 2 | 500 | -0.8 [-2.0 to -0.1] | -1.0 [-2.7 to -0.2] | -0.7 [-2.6 to -0.1] | 0.2 (-0.3 to 0.6) | -0.1 (-0.4 to 0.6) | -0.3 (-0.6 to 0.4) |
| Day 3 | 456 | -0.3 [-1.6 to 0.0] | -0.3 [-2.1 to 0.0] | -0.3 [-1.8 to 0.0] | 0.0 (-0.2 to 0.4) | -0.1 (-0.3 to 0.3) | -0.1 (-0.4 to 0.3) |
| Day 4 | 408 | -0.2 [-1.7 to 0.0] | -0.2 [-2.0 to 0.0] | -0.1 [-1.0 to -0.0] | -0.0 (-0.3 to 0.4) | -0.1 (-0.3 to 0.2) | -0.1 (-0.5 to 0.2) |
| Day 5 | 375 | -0.1 [-1.2-0.0] | -0.1 [-1.5-0.0] | 0.0 [-0.5-0.0] | -0.0 (-0.3 to 0.5) | -0.1 (-0.3 to 0.0) | -0.1 (-0.6 to 0.1) |
| Day 6 | 335 | -0.1 [-1.0-0.0] | -0.2 [-1.1-0.0] | 0.0 [-0.5-0.0] | 0.1 (-0.2 to 0.3) | -0.1 (-0.3 to 0.1) | -0.2 (-0.3 to 0.0) |
| Day 7 | 302 | 0.0 [-0.8-0.0] | -0.1 [-1.0-0.0] | 0.0 [-0.5-0.0] | 0.1 (-0.1 to 0.4) | 0.0 (-0.2 to 0.1) | -0.1 (-0.4 to 0.0) |
| Lowest daily RASS of -4 or -5, no. (%) | | | | | | | |
| Day 1 | 560 | 128 (71.1) | 149 (76.8) | 137 (73.7) | -5.7 (-15.1 to 3.7) | -2.5 (-12.3 to 7.2) | 3.1 (-6.1 to 12.4) |
| Day 2 | 500 | 45 (28.1) | 50 (29.1) | 51 (30.4) | -0.9 (-11.3 to 9.4) | -2.2 (-12.7 to 8.2) | -1.3 (-11.6 to 9.0) |
| Day 3 | 456 | 34 (23) | 32 (20.6) | 31 (20.3) | 2.3 (-7.6 to 12.3) | 2.7 (-7.3 to 12.7) | 0.4 (-9.0 to 9.8) |
| Day 4 | 408 | 26 (20.2) | 32 (23) | 21 (15) | -2.9 (-13.5 to 7.7) | 5.2 (-4.7 to 15.0) | 8.0 (-1.9 to 17.9) |
| Day 5 | 375 | 16 (13.7) | 25 (20) | 20 (15) | -6.3 (-16.5 to 3.9) | -1.4 (-10.9 to 8.1) | 5.0 (-5.1 to 15.0) |
| Day 6 | 335 | 19 (17.6) | 21 (18.9) | 15 (12.9) | -1.3 (-12.5 to 9.8) | 4.7 (-5.7 to 15.0) | 6.0 (-4.4 to 16.4) |
| Day 7 | 302 | 13 (13.1) | 13 (12.9) | 15 (14.7) | 0.3% (-9.3 to 9.8) | -1.6 (-12.1 to 9.0) | -1.8 (-12.3 to 8.6) |
| Mean Glasgow coma scale (GCS) | | | | | | | |

| | | | | | | | |
|---|---|---|---|---|---|---|---|
| score, median [IQR] | | | | | | | |
| Day 1 | 560 | 8.1 [5.4-10.1] | 7.8 [5.4-10.0] | 7.9 [5.5-10.3] | 0.3 (-0.5 to 1.6) | 0.2 (-1.1 to 1.2) | -0.1 (-1.6 to 0.9) |
| Day 2 | 497 | 11.0 [8.3-14.7] | 10.2 [7.6-14.9] | 10.7 [8.0-14.1] | 0.8 (-0.5 to 2.2) | 0.3 (-0.9 to 1.9) | -0.5 (-1.6 to 0.6) |
| Day 3 | 455 | 13.8 [8.3-15.0] | 11.7 [8.6-15.0] | 13.8 [8.6-15.0] | 2.1 (-0.9 to 3.7) | 0.0 (-2.2 to 2.2) | -2.1 (-3.7 to 1.1) |
| Day 4 | 409 | 14.0 [8.9-15.0] | 13.7 [8.3-15.0] | 14.2 [9.6-15.0] | 0.3 (-0.6 to 3.4) | -0.2 (-1.3 to 0.8) | -0.4 (-3.5 to 0.3) |
| Day 5 | 375 | 14.4 [10.8-15.0] | 13.7 [9.0-15.0] | 14.5 [10.5-15.0] | 0.7 (-0.2 to 3.4) | -0.1 (-0.7 to 0.6) | -0.8 (-3.5 to 0.0) |
| Day 6 | 336 | 14.6 [10.3-15.0] | 14.0 [9.5-15.0] | 14.5 [11.0-15.0] | 0.6 (-0.4 to 3.1) | 0.1 (-0.9 to 0.7) | -0.5 (-3.0 to 0.4) |
| Day 7 | 305 | 14.7 [10.9-15.0] | 14.0 [10.1-15.0] | 14.7 [11.0-15.0] | 0.7 (-0.2 to 2.8) | 0.1 (-0.7 to 0.8) | -0.7 (-2.8 to 0.3) |
| Lowest daily GCS < 8, no. (%) | | | | | | | |
| Day 1 | 560 | 140 (77.8) | 155 (80.3) | 154 (82.4) | -2.5 (-11.3 to 6.3) | -4.6 (-13.3 to 4.1) | -2.0 (-10.4 to 6.3) |
| Day 2 | 497 | 53 (33.1) | 65 (38) | 64 (38.6) | -4.9 (-15.8 to 6.0) | -5.4 (-16.4 to 5.6) | -0.5 (-11.5 to 10.4) |
| Day 3 | 455 | 42 (28.2) | 44 (28.4) | 40 (26.5) | -0.2 (-10.5 to 10.1) | 1.7 (-9.1 to 12.5) | 1.9 (-8.8 to 12.5) |
| Day 4 | 409 | 35 (26.9) | 40 (28.8) | 30 (21.4) | -1.9 (-13.3 to 9.6) | 5.5 (-5.5 to 16.5) | 7.3 (-3.5 to 18.2) |
| Day 5 | 375 | 18 (15.4) | 26 (20.8) | 27 (20.3) | -5.4 (-15.9 to 5.1) | -4.9 (-15.2 to 5.3) | 0.5 (-9.9 to 10.9) |
| Day 6 | 336 | 20 (18.2) | 24 (21.8) | 21 (18.1) | -3.6 (-15.1 to 7.8) | 0.1 (-10.1 to 10.2) | 3.7 (-7.6 to 15.0) |
| Day 7 | 305 | 15 (15) | 16 (15.7) | 21 (20.4) | -0.7 (-11.3 to 9.9) | -5.4 (-16.8 to 6.1) | -4.7 (-16.2 to 6.8) |
| CAM-ICU Positive, no. (%) | | | | | | | |
| Day 1 | 426 | 111 (78.2) | 107 (77) | 109 (75.2) | 1.2 (-9.3 to 11.7) | 3.0 (-7.5 to 13.5) | 1.8 (-8.8 to 12.4) |
| Day 2 | 420 | 80 (58.4) | 87 (60.4) | 78 (56.1) | -2.0 (-14.2 to 10.2) | 2.3 (-10.1 to 14.7) | 4.3 (-7.9 to 16.5) |
| Day 3 | 358 | 55 (47.8) | 68 (56.2) | 66 (54.1) | -8.4 (-21.9 to 5.2) | -6.3 (-19.8 to 7.3) | 2.1 (-11.2 to 15.4) |
| Day 4 | 294 | 45 (50.6) | 62 (63.3) | 54 (50.5) | -12.7 (-27.9 to 2.5) | 0.1 (-14.1 to 14.2) | 12.8 (-1.6 to 27.2) |
| Day 5 | 235 | 42 (56) | 48 (60.8) | 43 (53.1) | -4.8 (-21.6 to 12.1) | 2.9 (-14.0 to 19.8) | 7.7 (-8.9 to 24.2) |
| Day 6 | 201 | 34 (55.7) | 43 (58.9) | 34 (50.7) | -3.2 (-21.5 to 15.2) | 5.0 (-13.9 to 23.8) | 8.2 (-9.7 to 26.0) |

| | | | | | | | |
|---|---|---|---|---|---|---|---|
| Day 7 | 165 | 29 (55.8) | 40 (67.8) | 24 (44.4) | -12.0 (-31.8 to 7.8) | 11.3 (-9.5 to 32.1) | 23.4 (3.8 to 43.0) |
| Receipt of intravenous analgesia or sedation, no. (%) | | | | | | | |
| Day 1[a] | 566 | 176 (97.2) | 189 (95.5) | 183 (97.9) | 1.8 (-2.5 to 6.1) | -0.6 (-4.3 to 3.1) | -2.4 (-6.5 to 1.7) |
| Day 2 | 526 | 156 (92.9) | 164 (90.6) | 158 (89.3) | 2.2 (-4.1 to 8.6) | 3.6 (-3.0 to 10.2) | 1.3 (-5.5 to 8.1) |
| Day 3 | 486 | 109 (69.9) | 119 (70.8) | 115 (71.0) | -1.0 (-11.5 to 9.6) | -1.1 (-11.8 to 9.5) | -0.2 (-10.1 to 9.8) |
| Day 4 | 440 | 75 (53.2) | 93 (61.6) | 81 (54.7) | -8.4 (-20.4 to 3.6) | -1.5 (-13.7 to 10.6) | 6.9 (-5.0 to 18.7) |
| Day 5 | 399 | 57 (46) | 74 (54.8) | 69 (49.3) | -8.8 (-21.8 to 4.1) | -3.3 (-16.1 to 9.5) | 5.5 (-7.0 to 18.0) |
| Day 6 | 361 | 55 (47.8) | 69 (57.5) | 56 (44.4) | -9.7 (-23.2 to 3.9) | 3.4 (-10.0 to 16.8) | 13.1 (-0.1 to 26.3) |
| Day 7 | 324 | 41 (39) | 54 (49.5) | 47 (42.7) | -10.5 (-24.7 to 3.7) | -3.7 (-17.7 to 10.4) | 6.8 (-7.3 to 20.9) |

*Definitions of abbreviations:* RASS=Richmond Agitation and Sedation Scale; IQR=Interquartile range; CAM-ICU=Confusion Assessment Method for the ICU

a Days of medication use represent calendar days, with the day of enrollment as Day 1.

### Table E13. Other ventilator parameters on study days 1 to 7

| Ventilator Setting, median [IQR] | n | Volume Control N=181 | Pressure Control N=198 | Adaptive Pressure Control N=187 |
|---|---|---|---|---|
| Mean Peak End Expiratory Pressure – cm $H_2O$ | | | | |
| Day 1 | 556 | 5.1 [5.0-7.3] | 5.0 [5.0-7.5] | 5.0 [5.0-6.3] |
| Day 2 | 312 | 5.0 [5.0-5.7] | 5.0 [5.0-6.2] | 5.0 [5.0-6.1] |
| Day 3 | 198 | 5.0 [5.0-8.0] | 5.0 [5.0-6.5] | 5.0 [5.0-7.9] |
| Day 4 | 138 | 5.0 [5.0-9.1] | 5.8 [5.0-8.0] | 5.0 [5.0-7.8] |
| Day 5 | 108 | 5.0 [5.0-9.1 | 6.1 [5.0-8.0] | 5.0 [5.0-8.0] |
| Day 6 | 86 | 5.4 [5.0-8.5] | 7.0 [5.0-8.0] | 5.0 [5.0-8.0] |
| Day 7 | 65 | 5.0 [5.0-8.0] | 6.4 [5.0-8.0] | 5.0 [5.0-8.0] |
| Mean fraction of inspired oxygen ($FiO_2$) | | | | |
| Day 1 | 565 | 0.42 [0.36-0.53] | 0.42 [0.36-0.55] | 0.40 [0.36-0.48] |
| Day 2 | 322 | 0.40 [0.30-0.50] | 0.40 [0.31-0.48] | 0.39 [0.31-0.43] |
| Day 3 | 208 | 0.40 [0.31-0.56] | 0.40 [0.31-0.51] | 0.40 [0.30-0.45] |
| Day 4 | 148 | 0.40 [0.31-0.58] | 0.44 [0.32-0.54] | 0.37 [0.30-0.47] |
| Day 5 | 115 | 0.42 [0.31-0.53] | 0.41 [0.36-0.49] | 0.36 [0.30-0.54] |
| Day 6 | 92 | 0.44 [0.39-0.58] | 0.42 [0.34-0.52] | 0.37 [0.31-0.51] |
| Day 7 | 70 | 0.42 [0.40-0.60] | 0.41 [0.33-0.55] | 0.36 [0.31-0.58] |
| Mean plateau pressure, cm $H_2O$ | | | | |
| Day 1 | 302 | 19.2 [16.0-23.8] | 18.0 [15.0-23.5] | 19.6 [15.0-23.0] |
| Day 2 | 140 | 19.3 [15.7-24.3] | 19.2 [15.3-23.0] | 18.2 [15.8-21.6] |
| Day 3 | 90 | 18.3 [15.0-24.7] | 19.5 [16.5-25.9] | 19.5 [15.5-25.8] |
| Day 4 | 57 | 18.0 [16.0-24.3] | 17.0 [14.3-24.1] | 20.0 [19.2-26.5] |
| Day 5 | 50 | 21.8 [18.2-26.1] | 20.0 [14.5-27.0] | 22.5 [18.0-26.0] |
| Day 6 | 40 | 20.8 [17.0-25.1] | 18.5 [15.0-27.3] | 21.0 [17.2-25.8] |
| Day 7 | 31 | 23.0 [20.3-24.5] | 24.0 [15.2-29.0] | 23.0 [20.0-27.0] |
| Mean measured respiratory rate, per minute | | | | |
| Day 1 | 566 | 21.9 [19.4-26.7] | 22.1 [18.0-26.8] | 21.1 [18.5-25.8] |
| Day 2 | 331 | 21.8 [18.8-26.8] | 21.9 [17.9-26.3] | 20.6 [17.0-24.3] |
| Day 3 | 220 | 22.1 [19.6-27.4] | 22.9 [19.9-26.9] | 22.6 [17.2-25.5] |
| Day 4 | 161 | 22.2 [19.1-26.7] | 23.0 [19.2-26.9] | 21.6 [17.2-25.6] |
| Day 5 | 126 | 22.4 [19.2-29.3] | 21.1 [18.7-29.0] | 20.2 [15.9-25.5] |
| Day 6 | 101 | 24.4 [18.3-29.2] | 21.1 [17.7-27.0] | 22.9 [17.3-26.6] |
| Day 7 | 78 | 25.5 [21.3-29.9] | 23.1 [18.3-27.1] | 23.2 [18.1-28.2] |

**Table E14. Blood gas laboratory values on study days 1 to 7**

| Laboratory value[a], median [IQR] | n | Volume Control N=181 | Pressure Control N=198 | Adaptive Pressure Control N=187 |
|---|---|---|---|---|
| Arterial pH | | | | |
| Day 1 | 227 | 7.28 [7.18-7.35] | 7.28 [7.19-7.37] | 7.32 [7.20-7.40] |
| Day 2 | 117 | 7.36 [7.31-7.41] | 7.37 [7.32-7.43] | 7.38 [7.32-7.43] |
| Day 3 | 72 | 7.38 [7.34-7.45] | 7.36 [7.31-7.44] | 7.38 [7.35-7.43] |
| Day 4 | 58 | 7.36 [7.29-7.43] | 7.39 [7.33-7.43] | 7.40 [7.36-7.51] |
| Day 5 | 54 | 7.34 [7.30-7.39] | 7.40 [7.29-7.45] | 7.46 [7.29-7.48] |
| Day 6 | 48 | 7.34 [7.32-7.37] | 7.39 [7.32-7.43] | 7.35 [7.29-7.47] |
| Day 7 | 36 | 7.38 [7.32-7.41] | 7.38 [7.35-7.43] | 7.24 [7.18-7.39] |
| Arterial $PaCO_2$, mmHg | | | | |
| Day 1 | 232 | 43 [36-50] | 42 [34-51] | 43 [35-52] |
| Day 2 | 114 | 39 [36-46] | 42 [35-47] | 43 [34-48] |
| Day 3 | 72 | 40 [32-45] | 36 [32-50] | 40 [36-48] |
| Day 4 | 57 | 42 [37-46] | 42 [34-56] | 41 [38-47] |
| Day 5 | 53 | 39 [36-51] | 41 [37-48] | 46 [35-51] |
| Day 6 | 47 | 43 [42-50] | 41 [37-51] | 47 [41-51] |
| Day 7 | 35 | 43 [36-48] | 46 [38-60] | 51 [47-56] |
| Arterial $PaO_2$, mmHg | | | | |
| Day 1 | 232 | 107 [81-160] | 107 [73-165] | 108 [89-172] |
| Day 2 | 113 | 100 [86-129] | 92 [71-116] | 92 [76-116] |
| Day 3 | 72 | 94 [81-121] | 99 [85-139] | 104 [74-126] |
| Day 4 | 57 | 98 [82-123] | 88 [71-102] | 98 [80-149] |
| Day 5 | 53 | 95 [78-127] | 87 [79-100] | 86 [79-132] |
| Day 6 | 47 | 96 [70-116] | 89 [79-115] | 86 [79-90] |
| Day 7 | 35 | 105 [76-125] | 86 [76-129] | 71 [68-97] |
| Venous pH | | | | |
| Day 1 | 380 | 7.30 [7.20-7.36] | 7.34 [7.26-7.40] | 7.32 [7.24-7.39] |
| Day 2 | 163 | 7.36 [7.31-7.41] | 7.37 [7.32-7.43] | 7.37 [7.30-7.43] |
| Day 3 | 118 | 7.37 [7.30-7.42] | 7.41 [7.31-7.45] | 7.39 [7.32-7.44] |
| Day 4 | 84 | 7.34 [7.28-7.44] | 7.36 [7.31-7.43] | 7.39 [7.36-7.44] |
| Day 5 | 76 | 7.36 [7.32-7.43] | 7.40 [7.33-7.42] | 7.40 [7.34-7.44] |
| Day 6 | 63 | 7.42 [7.37-7.48] | 7.40 [7.34-7.44] | 7.36 [7.33-7.38] |
| Day 7 | 57 | 7.38 [7.35-7.41] | 7.36 [7.34-7.42] | 7.36 [7.33-7.43] |
| Venous $pCO_2$, mmHg | | | | |
| Day 1 | 384 | 49 [44-60] | 47 [40-55] | 48 [40-56] |
| Day 2 | 157 | 43 [37-50] | 46 [39-51] | 44 [38-55] |

| | | | | |
|---|---|---|---|---|
| Day 3 | 111 | 45 [40-58] | 49 [38-56] | 46 [38-56] |
| Day 4 | 78 | 51 [46-58] | 51 [42-60] | 48 [39-60] |
| Day 5 | 69 | 54 [48-61] | 53 [47-62] | 47 [39-63] |
| Day 6 | 59 | 48 [40-57] | 54 [45-62] | 45 [37-66] |
| Day 7 | 52 | 50 [43-61] | 53 [48-68] | 48 [40-66] |

a Laboratory values shown represent the first result on each study day.

**Table E15. Primary analysis of the primary outcome**

| Variable | Odds Ratio | 95% CI | P-value |
|---|---|---|---|
| Study group | | | 0.60 |
| Volume control | 0.79 | 0.41 - 1.53 | |
| Pressure control | 0.77 | 0.46 - 1.29 | |
| Adaptive pressure control | referent | | |
| Time - days | 1.62 | 0.83 – 3.16 | 0.13 |

For the primary outcome, the intraclass correlation calculated with the use of an analysis-of-variance method was 0.01.

**Table E16. Receipt of supportive therapies to day 28.**

| | n | Volume Control N=181 | Pressure Control N=198 | Adaptive Pressure Control N=187 |
|---|---|---|---|---|
| Mechanical ventilation, no. (%) | | 181 (100.0) | 198 (100.0) | 187 (100.0) |
| Duration, median [IQR], days | 566 | 3 [2-6] | 3 [2-6] | 3 [2-6] |
| Duration among survivors, median [IQR], days | 380 | 3 [2-5] | 3 [2-6] | 2 [2-4] |
| Receipt of vasopressors | | 112 (61.9) | 126 (63.6) | 119 (63.6) |
| Duration, median [IQR], days | 566 | 2 [0-3] | 1 [0-3] | 1 [0-3] |
| Duration among survivors, median [IQR], days | 380 | 1 [0-2] | 1 [0-2] | 1 [0-2] |
| ICU admission | | 181 (100.0) | 198 (100.0) | 187 (100.0) |
| Duration, median [IQR], days | 566 | 4 [3-8] | 4 [2-8] | 4 [3-8] |
| Duration among survivors, median [IQR], days | 380 | 4 [3-7] | 4 [3-9] | 4 [3-8] |
| Hospitalization | | 181 (100.0) | 198 (100.0) | 187 (100.0) |
| Duration, median [IQR], days | 566 | 9 [4-17] | 8 [4-18] | 8 [5-14] |
| Duration among survivors, median [IQR], days | 380 | 12 [6-21] | 10 [6-21] | 10 [6-20] |

All outcomes censored at 28 days after enrollment. Duration of receipt of life-support is determined from the first receipt at or after enrollment until the last receipt prior to death, hospital discharge, or 28 days after enrollment, whichever occurs first.

**Table E17. Sensitivity analyses: adjusted primary analysis**

| **Adjusted primary analysis:** Primary analysis adjusted for prespecified baseline covariates of age, sex, race and ethnicity, source of ICU admission, vasopressor receipt, acute diagnoses at enrollment, and SOFA score. | | | |
|---|---|---|---|
| **Variable** | **Odds Ratio** | **95% CI** | **P-value** |
| Study group | | | 0.64 |
| Volume control | 0.82 | ( 0.41, 1.65 ) | |
| Pressure control | 0.77 | ( 0.45, 1.34 ) | |
| Adaptive pressure control | Referent | ( 0.75, 2.24 ) | |
| Time – days (200:63) | 1.27 | ( 0.63, 2.56 ) | 0.50 |
| Age – years (67.5:41.8) | 0.75 | ( 0.47, 1.19 ) | 0.23 |
| Gender (male) | 1.62 | ( 1.17, 2.25 ) | 0.004 |
| Race and ethnicity | | | 0.18 |
| Hispanic | referent | | |
| Non-Hispanic Black | 0.41 | ( 0.17, 0.98 ) | |
| Non-Hispanic White | 0.59 | ( 0.27, 1.32 ) | |
| Other or unknown race | 0.52 | ( 0.19, 1.36 ) | |
| Source of admission to study ICU | | | <0.001 |
| Other ICU at study hospital | referent | | |
| ED at study hospital | 1.95 | ( 0.92, 4.13 ) | |
| Hospital ward at study hospital | 0.76 | ( 0.34, 1.71 ) | |
| OR or PACU at study hospital | 2.73 | ( 1.02, 7.28 ) | |
| Outside hospital | 1.19 | ( 0.51, 2.80 ) | |
| Sepsis or septic shock | 0.43 | ( 0.30, 0.62 ) | <0.001 |
| Pneumonia | 0.55 | ( 0.38, 0.80 ) | 0.002 |
| Cardiac arrest | 0.28 | ( 0.16, 0.49 ) | <0.001 |
| COPD exacerbation | 1.26 | ( 0.63, 2.53 ) | 0.51 |
| Asthma exacerbation | 1.68 | ( 0.35, 8.06 ) | 0.52 |
| Vasopressor receipt | 0.55 | ( 0.38, 0.79 ) | 0.002 |
| SOFA score at enrollment (10.75:5) | 0.69 | ( 0.54, 0.90 ) | 0.01 |

*Definitions of abbreviations:* ED=Emergency department; ICU= Intensive care unit; OR=Operating room; PACU= Post-anesthesia care unit

**Table E18. Sensitivity analyses: full cohort including patients enrolled in washout periods**

| **Primary analysis with all enrolled patients**: including patients admitted during washout periods | | | |
|---|---|---|---|
| **Variable** | **Odds Ratio** | **95% CI** | **P-value** |
| Study group | | | 0.50 |
| Volume control | 0.75 | 0.41 – 1.39 | |
| Pressure control | 0.75 | 0.46 – 1.21 | |
| Adaptive pressure control | referent | | |
| Time – days | 1.66 | 0.89 – 3.11 | 0.21 |
