## Supplementary material for "Effect of Ventilator Mode on Ventilator-Free Days in Critically Ill Adults: A Randomized Trial": Protocol and Analysis Plan

Supplementary Appendix

Study Protocol and Statistical Analysis Plan

| | |
|---|---|
| **Trial:** | Mode of Ventilation in Critical Illness (MODE) trial |
| **Manuscript:** | Effect of Ventilator Mode on Ventilator-free Days in Critically Ill Adults: A Randomized Clinical Trial |
| **Clinicaltrials.gov:** | NCT05563779 |
| **Authors:** | Kevin P. Seitz; Bradley D. Lloyd; Li Wang; Matthew S. Shotwell; Edward T. Qian; Amelia L. Muhs; Roger K. Richardson, J. Craig Rooks; Vanessa Hennings-Williams; Claire E. Sandoval; Whitney D. Richardson; Tracy L. Morgan; Amber N. Thompson; Pamela G. Hastings; Terry P. Ring; Joanna L. Stollings; Erica M. Talbot; David J. Krasinski; Bailey R. DeCoursey; Tanya K. Marvi; Stephanie C. DeMasi; Kevin W. Gibbs; Wesley H. Self; Amanda S. Mixon; Todd W. Rice; Matthew W. Semler; Jonathan D. Casey for the Pragmatic Critical Care Research Group |

This Supplementary Appendix contains the following items:

1) Initial Protocol (dated 9/26/2022)
2) Final Protocol (dated 7/21/2023)
3) Summary of changes to Study Protocol
4) Citations for Statistical Analysis Plan and summary of changes

### Mode Of Ventilation During Critical IllnEss (MODE) Pilot Trial

**Clinical Trial Protocol**

**Version 1.0**

**Principal Investigator**

Kevin Seitz, MD, MSc
*Division of Allergy, Pulmonary, and Critical Care Medicine*
*Department of Medicine*
*Vanderbilt University School of Medicine*

**Co-Investigators**

Jonathan Casey, MD, MSc
Matthew Semler, MD, MSc
Todd Rice, MD, MSc
Edward Qian, MD
Bradley Lloyd, RT-ACCS
*Division of Allergy, Pulmonary, and Critical Care Medicine*
*Department of Medicine*
*Vanderbilt University School of Medicine*

Wesley Self, MD, MPH
*Department of Emergency Medicine*
*Vanderbilt University School of Medicine*

#### Table of Contents

**1.0 Trial Summary 4**

**2.0 Background 6**

**3.0 Aims and Hypotheses 7**

3.1 Study Aims: 7

3.2 Hypotheses: 7

**4.0 Study Description 7**

4.1 Rationale for Cluster-Level Allocation 8

**5.0 Inclusion and Exclusion Criteria 9**

5.1 Inclusion Criteria: 9

5.2 Exclusion Criteria: 9

**6.0 Enrollment / Randomization 9**

6.1 Study Site: 9

6.2 Study Population: 9

6.3 Enrollment: 9

6.4 Consent: 9

6.5 Randomization: 11

**7.0 Study Procedures 11**

7.1 Study Interventions: 11

7.2 Study Co-interventions: 13

7.3 Monitoring Ventilator Mode: 14

7.4 Blinding: 14

7.5 Data Collection: 14

7.6 Outcome Measures: 15

**8.0 Risks and Benefits 16**

**9.0 Safety Monitoring and Adverse Events 16**

9.1 Adverse Event Definitions: 17

9.2 Monitoring for Adverse Events: 18

9.3 Recording and Reporting Adverse Events: 18

9.4 Clinical Outcomes that may be Exempt from Adverse Event Recording and Reporting: 19

9.5 Unanticipated Problems involving Risks to Subjects or Others: 19

**10.0 Statistical Considerations 20**

**11.0 Privacy / Confidentiality Issues 22**

**12.0 Follow-up and Record Retention 22**

**13.0 Data Safety and Monitoring Board (DSMB) 23**

**14.0 References 23**

**Appendix 1: Ventilator Guidelines 29**

**Appendix 2. Ventilator Guidelines Table 31**

**Appendix 3: Spontaneous Breathing Trials and Ventilator Discontinuation Guidelines 32**

#### 1.0 Trial Summary

| | |
|---|---|
| **Title** | Mode Of Ventilation During Critical IllnEss (MODE) Pilot Trial |
| **Background** | Landmark trials in critical care have demonstrated that, among critically ill adults receiving invasive mechanical ventilation, the use of low tidal volumes and low airway pressures prevents lung injury and improves patient outcomes. Limited evidence, however, informs the best method of mechanical ventilation to achieve these targets. To provide mechanical ventilation, clinicians must choose between modes of ventilation that directly control tidal volumes ("volume control"), modes that directly control the inspiratory airway pressure ("pressure control"), and modes that are hybrids ("adaptive pressure control"). Whether the choice of the mode used to target low tidal volumes and low inspiratory plateau pressures affects clinical outcomes for critically ill adults receiving mechanical ventilation is unknown. All three modes of mechanical ventilation are commonly used in clinical practice. A large, multicenter randomized trial comparing available modes of mechanical ventilation is needed to understand the effect of each mode on clinical outcomes. We will conduct a pilot trial evaluating the feasibility of comparing modes for mechanically ventilated ICU patients. |
| **Study Design** | Prospective, unblinded, cluster-randomized, multiple-crossover pilot trial |
| **Trial Groups** | 1. Volume Control group<br>2. Pressure Control group<br>3. Adaptive Pressure Control group |
| **Inclusion Criteria** | 1. Age ≥ 18 years<br>2. Receiving mechanical ventilation through an endotracheal tube or tracheostomy<br>3. Admitted to the study ICU |
| **Exclusion Criteria** | 1. Patient is pregnant<br>2. Patient is a prisoner<br>3. Patient was receiving invasive mechanical ventilation at place of residence prior to hospital admission<br>4. Patient is receiving extracorporeal membrane oxygenation at the time of admission to the study ICU |
| **Randomization** | In this cluster-randomized cluster-crossover trial, the entire study ICU will be assigned to a mode of ventilation (volume control, pressure control, or adaptive pressure control) and will switch between modes every month in an order determined by computer-generated randomization using permuted blocks of 3. |
| **Primary Outcome** | Ventilator-free days to day 28 after enrollment |
| **Exploratory Outcomes** | Feasibility Outcomes<br>• Exposure to assigned study mode in first 3 days: Proportion of time receiving invasive mechanical ventilation in the study ICU during the first 72 hours after enrollment in which the patient was receiving the assigned mode of ventilation |

| | |
|---|---|
| | • Adherence to study mode in first 3 days: Proportion of time receiving invasive mechanical ventilation in the study ICU during the first 72 hours after enrollment in which the patient was receiving the assigned mode of ventilation, excluding spontaneous breathing trials<br>• Time from enrollment to initiation of assigned mode of mechanical ventilation<br>• Number of patients with a "Mode Modification Sheet" completed by treating clinicians<br>Exploratory Efficacy and Safety Outcomes<br>• Median exhaled tidal volume on each study day<br>• Exhaled tidal volumes above target range: Proportion of recorded breaths with exhaled tidal volume values above the target range (>8mL/kg PBW) on each study day<br>• Hypoxemia during mechanical ventilation: Episodes of hypoxemia during mechanical ventilation: SpO2 < 85% for more than 5 minutes<br>• Severe acidemia during mechanical ventilation: Episodes of severe acidemia during mechanical ventilation: pH < 7.1 on blood gas<br>• Number of blood gas laboratory tests per day while receiving mechanical ventilation<br>• Pneumomediastinum or pneumothorax during course of mechanical ventilation<br>• SOFA score daily on the first 7 study days<br>• Delirium and coma-free days to day 28<br>Exploratory Clinical Outcomes:<br>• ICU-free days to study day 28<br>• Hospital-free days to study day 28<br>• In-hospital mortality to study day 28 |
| **Analysis** | The primary analysis of the primary outcome (ventilator-free days) will be conducted in an intention-to-treat fashion using a proportional odds model with independent covariates of group assignment (volume control, pressure control, or adaptive pressure control) and time since beginning enrollment (in days). |
| **Sample Size** | The trial will be conducted for a total of 9 months as this is the duration needed to evaluate the feasibility of the cluster-randomized multiple-crossover design. Based on data from a prior trial in the same ICU, we expect to enroll 675 patients, of whom 69 will be enrolled during washout periods and 606 patients will be included in the primary analysis. |
| **Duration** | 9 months |

#### 2.0 Background

Approximately 2-3 million critically ill adults receive invasive mechanical ventilation in the ICU each year.[1–3] Mechanical ventilation provides support for ventilation and gas exchange while patients recover from the underlying causes of their respiratory failure. Mechanical ventilation itself, however, may injure the lungs, diaphragm, and other organs,[4,5] and in-hospital mortality for mechanically ventilated ICU patients remains 25-35%.[6]

For patients on mechanical ventilation, tidal volume and inspiratory airway pressures are directly related. Increasing tidal volumes increases airway pressures and vice versa. The ventilator can be set by clinicians to deliver breaths in one of two ways: either by controlling the volume ("volume control") or the pressure ("pressure control") administered during the inspiratory phase of the breath. By controlling one factor, each mode provides indirect control over the other, as a dependent variable that varies proportionally based on the patient's lung physiology.[7] Modern ventilators also offer hybrid modes such as adaptive pressure control, allowing clinicians to set a target volume for which the ventilator titrates and delivers the pressure controlled breath that is expected to be adequate based on the results of previous breaths (e.g. Pressure Regulated Volume Control). The mode of mechanical ventilation must be set for each of 2-3 million ICU patients who receive invasive mechanical ventilation. Yet, whether choice of mode affects outcomes remains uncertain. [8–10]

Landmark clinical trials in critically ill patients have demonstrated that using smaller tidal volumes and plateau pressures (Low Tidal Volume Ventilation) reduces lung injury and improves patient outcomes, both for patients with ARDS[11] and for patients without ARDS.[12] Each of the commonly used modes of mechanical ventilation (volume control, pressure control, adaptive pressure control) can achieve these low tidal volume and low plateau pressure targets – but minimal data exist to guide the choice of which ventilator mode should be used to achieve these targets. Current guidelines recommend delivering low tidal volumes and low plateau pressures to all patients on mechanical ventilation and make no recommendations regarding the mode of mechanical ventilation used to achieve these targets. [13–18] Volume control, pressure control, and adaptive pressure control can each be used to accomplish adequate gas exchange in critically ill patients, but they differ in how they control tidal volumes and inspiratory pressures and synchronize with patient efforts to breathe.[19,20]

Only two clinical trials have directly compared volume control and pressure control as the modes of mechanical ventilation for critically ill adults. The trials were small and lacked adequate power to evaluate for clinically significant difference in patient-important outcomes.[21,22] Further, they were performed more than 20 years ago, prior to many modern practices of critical care including low tidal volume ventilation. Evidence comparing adaptive modes to volume or pressure control are similarly limited to small trials with mixed results.[23,24] Reviews and meta-analyses of small and heterogenous studies of physiology have also not demonstrated clear superiority of volume control, pressure control, or adaptive pressure control.[8,9,20]

All three modes are in common use in large, international cohorts of ARDS and non-ARDS patients, with a slight increase in the use of pressure modes over time.[12,25,26] Pressure control modes are the most common modes in international cohorts.[12,26–28] While ventilator settings for critically ill patients in the United States are not as well described, the most recent data on patients in the US receiving contemporary critical care, from a large multi-center cohort of moderate-severe ARDS patients, demonstrated that on the first day of mechanical ventilation with ARDS, volume control was used in approximately half of patients, with most of the remainder receiving pressure regulated modes in either pressure control or adaptive pressure control.[29] Among all patients receiving mechanical ventilation in the Vanderbilt Medical ICU in 2021, 35% of breaths were in volume control, 31% were in pressure control, and 35% were in adaptive pressure control.

Because millions of critically ill adults receive invasive mechanical ventilation each year, a mode of ventilation must be selected for every patient, three modes are used commonly in current practice, and no high-quality data inform the effect of mode of ventilation on patient outcomes, a large, randomized trial is needed to inform the optimal choice of ventilator mode for mechanically ventilated ICU patients. Before such a trial can be conducted, however, data are needed to demonstrate the feasibility of maintaining adherence to ventilator modes in a pragmatic trial.[30–32]

#### 3.0 Aims and Hypotheses

##### 3.1 Study Aims:

- **Feasibility Aim:** To assess the feasibility of a cluster-randomized, multiple-crossover trial comparing volume control, pressure control, and adaptive pressure control modes.

- **Clinical Aim:** To evaluate the effect of the same ventilator mode intervention on days alive and free of invasive mechanical ventilation in the same population of mechanically ventilated critically ill patients.

##### 3.2 Hypotheses:

- **Feasibility Hypothesis:** Comparing volume control, pressure control, and adaptive pressure control ventilation among mechanically ventilated ICU patients will be feasible.

- **Clinical Hypothesis:** Use of an adaptive pressure control mode for mechanically ventilated ICU patients will result in more days alive and free of invasive mechanical ventilation than use of either a volume control or pressure control mode.

#### 4.0 Study Description

To address the aims outlined above, we propose the Mode Of Ventilation During Critical IllnEss (MODE) Pilot Trial. This trial will be a prospective, single-center, unblinded, cluster-randomized, multiple-crossover trial conducted in the medical ICU at Vanderbilt University Medical Center. During the 9 months of enrollment in the MODE trial, the entire medical ICU will be assigned to a single ventilator mode with the assigned mode alternating between volume control, pressure control, and adaptive pressure control modes every month in a randomly generated sequence. All patients who receive invasive mechanical ventilation and meet inclusion criteria without meeting exclusion criteria will be enrolled at the initiation of mechanical ventilation. The trial will control only the ventilator mode during controlled mechanical ventilation. All other aspects of clinical care for patients will remain at the discretion of treating clinicians.

#### 4.1 Rationale for Cluster-Level Allocation:

Group assignment in the MODE trial occurs at the level of the ICU (cluster) for multiple reasons. In routine clinical care in the study ICU, 2 to 4 respiratory therapists set the ventilator mode and adjust settings to maintain gas exchange and patient comfort for all mechanically ventilated adults, with input from nurses and physicians. The management of mechanical ventilation (selection of fraction of inspired oxygen, titration of positive end-expiratory pressure, screening for and performance of spontaneous breathing trials) is governed by unit-wide protocols implemented by the 2 to 4 respiratory therapists for all patients in the unit. Assigning the entire unit to a single ventilator mode both emulates the way mechanical ventilation is managed during clinical care and limits contamination that might result from a respiratory therapist managing multiple patients assigned to different modes of mechanical ventilation.

Additionally, mechanical ventilation can damage lungs even after brief periods of ventilation, and the initial settings have a unique significance. For example, during the minutes-to-hours of temporary mechanical ventilation for surgical procedures, harmful ventilator settings can affect organ function and clinical outcomes.[34] In critically ill patients, the initial hours of invasive mechanical ventilation are also the period with the highest risk of worsening lung injury, dyssynchrony, increased work of breathing, and hemodynamic compromise. Prior research has demonstrated that [1] the very first ventilator settings at the time the patient is being initiated on invasive mechanical ventilation in the ICU are associated with differences in mortality[35,36] and [2] the association between ventilator settings and mortality is larger for earlier settings compared to later settings.[33] The ventilator protocol used to set the ventilator mode in the study ICU begins immediately at the time of initiation of mechanical ventilation. Enrollment immediately after initiation of invasive mechanical ventilation minimizes pre-study exposure to other modes and ventilator settings, and it facilitates on-study separation between groups.

Recognizing the importance of initial ventilator settings, three recent clinical trials have examined other ventilator settings (i.e. end-expiratory pressure, tidal volume, and $SpO_2$ target) by attempting to enroll patients soon after the initiation of mechanical ventilation in the ICU. [12,37,38] In these patient-level, parallel-group trials, however, the logistical challenges of performing screening, enrollment, randomization, and study group assignment resulted in a significant gap between initiation of invasive mechanical ventilation and delivery of the respective trial interventions while also leading to the exclusion of 60-67% of eligible patients – raising concern for systematic exclusion of important patient groups (e.g. patients with higher acuity of illness). In MODE, group assignment at the cluster level allows enrollment immediately on initiation of invasive mechanical ventilation in the ICU. This approach emulates the manner in which ventilator settings are managed in practice, precludes systematic exclusion of important patient groups, decreases pre-study exposure to harmful ventilator settings, and facilitates early separation in the receipt of ventilator modes between groups.

#### 5.0 Inclusion and Exclusion Criteria

##### 5.1 Inclusion Criteria:

1. Age ≥ 18 years
2. Receiving mechanical ventilation through an endotracheal tube or tracheostomy
3. Admitted to the study ICU.

##### 5.2 Exclusion Criteria:

1. Patient is pregnant
2. Patient is a prisoner
3. Patient receiving invasive mechanical ventilation at place of residence prior to hospital admission
4. Patient receiving extracorporeal membrane oxygenation at the time of admission to the study ICU.

#### 6.0 Enrollment / Randomization

##### 6.1 Study Site:

- Medical Intensive Care Unit at Vanderbilt University Medical Center

##### 6.2 Study Population:

All adults located in the study ICU for whom the treating clinicians have decided invasive mechanical ventilation is required will be enrolled unless meeting exclusion criteria. Patients will be included regardless of age, gender, race, or other clinical factors.

##### 6.3 Enrollment:

All adult patients who do not meet exclusion criteria will be enrolled immediately upon receipt of invasive mechanical ventilation in the study ICU.

##### 6.4 Consent:

Volume control, pressure control, and adaptive pressure control are all common approaches to controlled mechanical ventilation for critically ill adults. All represent standard-of-care treatments in current clinical practice. Results from prior clinical trials do not demonstrate superiority of one approach over the other. Current clinical guidelines do not recommend any specific mode of mechanical ventilation.[14,16–18] As a result, significant variation exists in the use of volume control, pressure control, and adaptive pressure control for patients in routine clinical care with ARDS[29] and those without ARDS.[26] In the study ICU, all three modes are currently in use in routine care. Of all patients on mechanical ventilation in the study ICU in 2021, 35% of mandatory breaths were in volume control mode, 31% were in pressure control mode, and 35% were in adaptive pressure control. This trial will only enroll patients who would already receive continuous mandatory ventilation with one of these three modes as part of their routine clinical care. When the primary medical team feels that the patient needs care with a specific ventilator mode different from what is assigned by the study, the team can alter the ventilator mode for the patient and use the mode that they believe is the correct mode for the patient. Only patients for whom the primary team feels that the ventilator mode assigned by the study is acceptable will continue with that assigned ventilator mode. We will request a waiver of informed consent because the study involves minimal risk and obtaining informed consent would be impracticable.

Participation in this study involves minimal risk because:

- The three interventions being compared are commonly used in routine clinical care in the study ICU;
- All are interventions to which the patient could be exposed even if not participating in the study (all patients receiving the initiation of mechanical ventilation receive one of these modes of continuous mandatory ventilation);
- No established differences in risk and benefits are known to exist between the studied approaches to mechanical ventilation based on the currently available data; and
- The trial only determines the mode of ventilation when treating clinicians feel all three modes would be consistent with optimal care for the individual patient – otherwise treating clinicians select the mode of ventilation via a Mode Modification Sheet.

The MODE trial is designed as a cluster-randomized multiple-crossover trial to ensure that the mode selected at the time of initiation of mechanical ventilation is consistent with trial group assignment, preventing contamination between groups and capturing the period of mechanical ventilation in which ventilator settings have the strongest association with outcomes.

Obtaining informed consent before initiation of mechanical ventilation in the study ICU would be impracticable because:

- **The expected medical condition of patients at the time of initiation invasive mechanical ventilation in the ICU is critical.** All patients will be critically ill and receiving continuous intravenous sedation at the time of enrollment. Thus, all patients eligible for MODE will not have the capacity to provide informed consent. Further, data from prior trials in the same patient population and setting demonstrate that prior to intubation and initiation of mechanical ventilation, approximately 70% of patients eligible for the MODE trial will be experiencing encephalopathy (altered mental status) due to their illness. The anticipated median Glasgow coma scale score will be 11 (equivalent to moderate brain injury). Among the minority of patients whose level of consciousness is not impaired, 45-55% will be experiencing acute delirium. Further, family members or legally authorized representatives (LAR) are frequently unavailable when critically ill patients undergo intubation in the ED or ICU.
- **The intervention is delivered by unit-level protocols**. Mechanical ventilator settings and titrations are performed by respiratory therapists using unit-level protocols. In this cluster-randomized trial, the entire unit will be assigned to the same mode of mechanical ventilation for each 1-month block. Obtaining informed consent from every eligible patient in the ICU prior to emergency tracheal intubation and initiation of mechanical ventilation would be impracticable.

Because the study involves minimal risk, the study would not adversely affect the welfare or privacy rights of the participant, and obtaining informed consent would be impracticable, we will request a waiver of informed consent. Numerous previous randomized trials comparing two standards of care for interventions such as emergency intubation and methods of respiratory support during critical illness have been completed with waiver of informed consent.[39–47]

##### 6.5 Randomization:

During each one-month block of the study, the ICU will be assigned to either volume control, pressure control, or adaptive pressure control. The order of the study group assignments will be generated by computerized randomization using permuted blocks of 3 to minimize the impact of seasonal variation. The last 3 days of each block will be a washout period during which the ICU will continue to use the assigned mode, but new patients will not be included in the primary analysis. The 3-day washout period will limit the number of patients who experience a 'crossover' from one assigned mode to another while receiving a mandatory mode of mechanical ventilation.

#### 7.0 Study Procedures

##### 7.1 Study Interventions:

**Ventilator Mode:**

**Volume Control Group:** Patients assigned to the volume control group will receive volume control as the initial mode of ventilation and whenever they are receiving continuous mandatory invasive mechanical ventilation.

**Pressure Control Group:** Patients assigned to the pressure control group will receive pressure control as the initial mode of ventilation and whenever they are receiving continuous mandatory invasive mechanical ventilation.

**Adaptive Pressure Control Group:** Patients assigned to the adaptive pressure control group will receive adaptive pressure control as the initial mode of ventilation and whenever they are receiving continuous mandatory invasive mechanical ventilation.

Characteristics of each of the three modes of ventilation are presented in Table 1.

**Table 1. Characteristics of the modes of mechanical ventilation.**

| Group Assignment | Volume Control | Pressure Control | Adaptive Pressure Control |
|---|---|---|---|
| Ventilator Mode Designation | *Volume Control* | *Pressure Control* | *Pressure Regulated Volume Control* |
| Control Variable | Volume | Pressure | Pressure |
| Breath Sequence | Continuous mandatory ventilation | Continuous mandatory ventilation | Continuous mandatory ventilation |
| Targeting Scheme | Set-point (clinician sets target volume) | Set-point (clinician sets target pressure) | Adaptive (clinician sets target volume and ventilator titrates pressure to achieve target volume) |

*Ventilator Settings and Titration*:
In the study ICU and the United States generally, respiratory therapists have primary responsibility for determining the initial settings for invasive mechanical ventilation and titrating these settings for clinical goals (e.g., $SaO_2$, $pCO_2$, target tidal volume, target plateau pressure, etc.). To set and titrate mechanical ventilator settings, respiratory therapists use clinical protocols jointly developed by respiratory therapy and physician leaders. For patients enrolled in the MODE trial, respiratory therapists will use preexisting clinical protocols for volume control, pressure control, and adaptive pressure control in use in the study unit (Appendix 1-3).

*Initiation of Mechanical Ventilation:* All mechanically ventilated patients being initiated on invasive mechanical ventilation in an ICU initially receive a “mandatory” mode of ventilation (e.g., volume control, pressure control, or adaptive pressure control) immediately following intubation. Mandatory modes are necessary for patients being initiated on invasive mechanical ventilation in an ICU, because patients nearly uniformly received sedation or neuromuscular blockade for placement of the endotracheal tube and do not have the spontaneous respiratory effort required to allow a “spontaneous” mode of ventilation. When respiratory therapists select the initial ventilator settings for a patient being initiated on invasive mechanical ventilation in the ICU, they will select the assigned mode of ventilation (volume control, pressure control, or adaptive pressure control). As prior studies have demonstrated that ventilator settings early in patients’ course of invasive mechanical ventilation may affect clinical outcomes,[33] ensuring the first set of ventilator settings includes the mode of ventilation assigned by the study will prevent contamination with non-assignment ventilator modes and ensure delivery of the study intervention during a key period of patients’ critical illness.

*Maintenance of Mechanical Ventilation:*
Study protocol will determine the ventilator mode from enrollment until the first of: (1) extubation from mechanical ventilation, (2) transfer out of the study ICU, (3) completion of a ventilator Mode Modification Sheet by treating clinicians, or (4) end of the one-month study block. Study protocol does not determine the ventilator mode during time-periods in which the patient is not receiving invasive mechanical ventilation, is not receiving a continuous mandatory mode of ventilation (e.g., when patients

who are no longer deeply sedated are weaned to pressure support ventilation), is not physically located in the study ICU (e.g., during transport), or is undergoing an invasive procedure (e.g., bronchoscopy).

*Modification of Ventilator Mode:* At any time, any member of the clinical team (respiratory therapist, nurse, nurse practitioner, or physician) may use any mode of mechanical ventilation if determined to be needed for the optimal treatment of any specific patient. If modifying the mode of ventilation is necessary, the date, time, and reason for modification of the mode will be prospectively recorded.

Details of each mode modification will be collected and monitored on a Mode Modification Sheet. In our prior cluster-crossover trial using the same approach to modification of therapy assignment, treating clinicians exercised the ability to modify the assigned therapy for approximately 5% of patients.[39]

Anticipated examples of conditions for which treating clinicians may elect to override the assigned mode include:
- Refractory hypoxemia
- Peak pressures persistently high (>40 cm $H_20$)
- Asynchrony or dyssynchrony not amenable to changes within the assigned mode of ventilation
- Excessive work of breathing
- Barotrauma (pneumothorax or pneumomediastinum)
- Inability to limit tidal volumes delivered
- Intrinsic PEEP
- Other

**7.2 Study Co-interventions:**

*Ventilator Adjustment:* For all patients regardless of group assignment, existing clinical ventilator protocols will be used to target a tidal volume of 6 mL/kg PBW and a plateau pressure <30 cm $H_20$, while maintaining a respiratory rate less than 35 and pH of at least greater than 7.15, with a target of 7.3 to 7.45. (Appendix 1-3) Respiratory therapists will titrate volume or pressure settings according to the patient's ventilator mode to achieve these parameters and manage asynchrony and patient discomfort if it occurs.

*Liberation from Mechanical Ventilation:* Each day of mechanical ventilation, all patients in the study ICU are assessed for safety of a spontaneous awakening trial (SAT) and spontaneous breathing trial (SBT)[48] using the SAT and SBT safety criteria from the Awakening and Breathing Controlled (ABC) trial.[49] These criteria relate to the non-respiratory clinical status of the patient and the PEEP and FiO2 settings which will be managed similarly across groups. As such, the SAT and SBT procedures will be handled in the same way for each patient, regardless of study group assignment. Definitions of SAT and SBT failure and the ventilator settings and duration of the SBT will be those used in the ABC trial and used in routine clinical care in the study ICU. For patients who have passed an SAT and SBT, the decision to discontinue invasive mechanical ventilation will be made by the treating clinicians.

*Fraction of Inspired Oxygen ($FiO_2$) and Positive End-Expiratory Pressure (PEEP)*: Institutional protocols in the study ICU will ensure that mechanically ventilated patients in MODE on each study mode receive $FiO_2$ and PEEP titration according to the ARDSNet Lower PEEP/higher $FiO_2$ table (except for patients with severe ARDS, for whom the higher PEEP/lower $FiO_2$ table is applied)[50,51]

*Sedation and Analgesics*: Institutional protocols in the study ICU will ensure that mechanically ventilated patients in MODE on each study mode receive similar management of pain, agitation, and delirium[52] targeting Critical Care Pain Observation Tool (CPOT)[53], Richmond Agitation-Sedation Scale (RASS)[54,55], and Confusion Assessment Method for the ICU (CAM-ICU) scores.[56,57]

#### 7.3 Monitoring Ventilator Mode:

For all mechanically ventilated patients in the study ICU, ventilators display settings such as mode and measurements such as observed tidal volume on a digital display at the patient's bedside in real time. For critical measurements such as respiratory rate, minute ventilation, and peak pressures, audible alarms from the ventilator alert clinicians to potentially dangerous situations. These settings and measurements are sampled and uploaded automatically into the EMR frequently and are stored in the data warehouse. This will allow for much more accurate assessment of the ventilator modes that patients receive over the course of the day compared to prior trials which documented ventilator mode once or twice daily.[58,59]

***Feedback on Ventilator Mode Adherence:*** During the study, study personnel will monitor compliance with the assigned ventilator mode. Study personnel will remotely monitor ventilator modes up to four times daily from 4AM through 10PM Monday through Friday and during a convenience sample of weekend hours to provide feedback to treating clinicians and explore reasons why patients are not receiving the assigned mode of mechanical ventilation,

Study personnel will attend respiratory therapy group meetings, nursing unit board meetings, and ICU physician leadership meetings to educate staff about the study, solicit safety concerns and adverse events, and identify and address barriers to ventilator mode assignment compliance. This approach to daily monitoring and intermittent feedback to clinical personnel successfully achieved 80% compliance with the assigned intervention in a prior cluster-crossover trial of ventilator settings in the same ICU.[39]

#### 7.4 Blinding:

Similar to prior ventilator mode studies among critically ill adults, patients and clinicians will not be blinded to study group assignment.[21,58,59] Observer bias will be minimized by use of objective endpoints collected and automated data extraction from the EHR (see *Data Collection* below).

#### 7.5 Data Collection:

The MODE trial will primarily use structured data collected in routine clinical care, exported daily from the institution's EHR into an Enterprise Data Warehouse, along with data from the patient registration, billing, and laboratory clinical information systems. We have previously validated the quality of this method of data collection against the reference standard of two-physician manual chart review,[60] and the planned approach to electronic dataset generation for the MODE trial has already been successfully employed for the conduct of four prior pragmatic trials.[39,61–63]

**Electronically extracted data elements will include:**
*Enrollment (Day 0):* date and time of clinical events (hospital admission, ICU admission, intubation); source of admission to study ICU; age; sex; race; ethnicity; height, weight;[64] SOFA score;[65] Glasgow Coma Scale score;[66] Elixhauser Comorbidity Index;[67] history of present illness; vital signs (temperature, heart rate, systolic blood pressure, diastolic blood pressure, $SpO_2$); mechanical ventilator settings (mode, set and exhaled tidal volumes, set and actual respiratory rate, minute ventilation, set and total positive end-expiratory pressure, peak pressure, plateau pressure, inspiratory time, flow pattern, $FiO_2$); serum laboratory values (white blood cell count, hemoglobin, platelet count, sodium, potassium, bicarbonate, creatinine, bilirubin, alanine aminotransferase, aspartate aminotransferase, lactate, arterial and venous pH, $PaCO_2$,$PaO_2$, $SaO_2$).

*Daily On-Study (Days 0-28):* Vital signs, ventilator settings, screening for and performance of spontaneous awakening trials and spontaneous breathing trials, and serum laboratory values (as above); Mean daily dose of sedative infusions (propofol, dexmedetomidine, morphine, fentanyl, and midazolam); CAM status; RASS score; Daily completion of each element from ICU Liberation Bundle (A-F); Beginning and end time of Spontaneous Breathing Trials; vasopressor and inotrope infusions (norepinephrine, epinephrine, vasopressin, phenylephrine, dobutamine, angiotensin-2, dopamine); "Mode Modification Sheet" (use, time, reason); Characteristics of treating clinicians (respiratory therapist, attending, pulmonary fellow, registered nurse) providing direct care; Use of therapies for refractory hypoxemia (prone positioning, ECMO, neuromuscular blockade, inhaled pulmonary dilators); SOFA score; chest radiograph with bilateral infiltrates consistent with ARDS;[68] Stage II or greater AKI by KDIGO criteria;[69] cardiac arrest, pneumothorax or pneumomediastinum.

Termination (Days 0-28): Vital status at 28 days; time of liberation from invasive mechanical ventilation; receipt and duration of vasopressors; receipt and duration of renal replacement therapy; tracheostomy; duration of ICU and hospital admission.

##### 7.6 Outcome Measures:

**Primary Outcome:**
The primary outcome is ventilator-free days (VFDs) to day 28 after enrollment.

VFDs will be defined as the number of calendar days alive and free of invasive mechanical ventilation from the final receipt of invasive mechanical ventilation through 28 days after enrollment. For the assessment of VFDs, the day of enrollment (defined as the day on which the patient first receives invasive mechanical ventilation in a participating study location), will be considered to be day 0. Outcome ascertainment will cease at the time of hospital discharge or 28 days after enrollment, whichever occurs first. Receipt of invasive mechanical ventilation will be considered to end when patients undergo the final tracheal extubation or disconnection of the ventilator from a tracheostomy tube between enrollment and 28 days after enrollment. Patients who continue to receive invasive mechanical ventilation at day 28 will receive zero VFDs. Patients who die prior to day 28 will receive zero VFDs. Patients who are discharged from the hospital prior to day 28 and are receiving invasive mechanical ventilation at the time of discharge will receive zero VFDs. Patients who are removed from invasive mechanical ventilation and are discharged from the hospital without invasive mechanical ventilation prior to 28 days will be assumed to remain free of invasive mechanical ventilation between hospital discharge and day 28. For patients who are removed from invasive mechanical ventilation, return to invasive mechanical ventilation, and are subsequently removed again from invasive mechanical ventilation prior to day 28, VFDs will be counted from the final receipt of invasive mechanical ventilation prior to day 28.

**Feasibility Outcomes**

- Exposure to assigned study mode in first 3 days: proportion of time receiving invasive mechanical ventilation in the study ICU during the first 72 hours after enrollment in which the patient was receiving the assigned mode of ventilation
- Adherence to study mode in first 3 days: proportion of time receiving invasive mechanical ventilation in the study ICU during the first 72 hours after enrollment in which the patient was receiving the assigned mode of ventilation, excluding spontaneous breathing trials
- Time from enrollment to initiation of assigned mode of mechanical ventilation
- Number of patients with a “Mode Modification Sheet” completed by treating clinicians

**Exploratory Efficacy and Safety Outcomes**

- Median exhaled tidal volume on each study day
- Exhaled tidal volumes above target range: proportion of recorded breaths with exhaled tidal volume values above the target range (>8mL/kg PBW) on each study day
- Hypoxemia during mechanical ventilation: episodes of hypoxemia during mechanical ventilation: $SpO_2 < 85\%$ for more than 5 minutes
- Severe acidemia during mechanical ventilation: Episodes of severe acidemia during mechanical ventilation: pH < 7.1 on blood gas
- Number of blood gas laboratory tests per day while receiving mechanical ventilation
- Pneumomediastinum or pneumothorax during course of mechanical ventilation
- SOFA score daily on the first 7 study days
- Delirium and coma-free days to day 28

**Exploratory Clinical Outcomes:**

- ICU-free days to study day 28
- Hospital-free days to study day 28
- In-hospital mortality to study day 28

#### 8.0 Risks and Benefits

Although no risks are currently known to differ between mechanical ventilation with volume control, pressure control, and adaptive pressure control (all standard-of-care approaches in currently clinical care), it is possible that the results of the MODE trial will ultimately demonstrate a difference between the approaches in the risk of ventilator-free days, hypoxemia, delirium and coma free days, or another outcome. The benefits of this research will include understanding the effect of choice of ventilator mode on outcomes for the 2-3 million adults per year who receive invasive ventilation.

#### 9.0 Safety Monitoring and Adverse Events

Assuring patient safety is an essential component of this protocol. Mechanical ventilation with volume control, pressure control, and adaptive pressure control are all standard-of-care interventions that have been use in clinical practice for decades with an established safety profile. This protocol further ensures safety of its participants through:

a. Exclusion criteria designed to prevent enrollment of patients likely to experience adverse events from pressure control, volume control, or adaptive pressure control settings;
b. Permitting treating clinicians to change the ventilator mode at any time for any patient in whom they feel a ventilator mode different from the assigned mode is required for the safe treatment of the patient;
c. Systematic collection of outcomes relevant to the safety of ventilator mode selection;
d. Structured monitoring, assessment, recording, and reporting of adverse events.

##### 9.1 Adverse Event Definitions:

**Adverse Event –** An adverse event will be defined as any untoward or unfavorable medical occurrence in a human subject temporally associated with the subject's participation in the research, whether or not considered related to the subject's participation in the research. Adverse events will be classified according to the following characteristics:

- **Seriousness** – An adverse event will be considered "serious" if it:
    - Results in death;
    - Is life-threatening (defined as placing the patient at immediate risk of death);
    - Results in inpatient hospitalization or prolongation of existing hospitalization;
    - Results in a persistent or significant disability or incapacity;
    - Results in a congenital anomaly or birth defect; or
    - Based upon appropriate medical judgment, may jeopardize the patient's health and may require medical or surgical intervention to prevent one of the other outcomes listed in this definition.

- **Unexpectedness** – An adverse event will be considered "unexpected" if the nature, severity, or frequency is neither consistent with:
    - The known or foreseeable risk of adverse events associated with the procedures involved in the research that are described in the protocol-related documents, such as the IRB-approved research protocol; nor
    - The expected natural progression of any underlying disease, disorder, or condition of the subject experiencing the adverse event and the subject's predisposing risk factor profile for the adverse event.

- **Relatedness** – The strength of the relationship of an adverse event to a study intervention or study procedure will be defined as follows:
    - Definitely Related: The adverse event follows (1) a reasonable, temporal sequence from a study procedure AND (2) cannot be explained by the known characteristics of the patient's clinical state or other therapies AND (3) evaluation of the patient's clinical state indicates to the investigator that the experience is definitely related to study procedures.
    - Probably or Possibly Related: The adverse event meets some but not all of the above

criteria for "Definitely Related".

- Probably Not Related: The adverse event occurred while the patient was on the study but can reasonably be explained by the known characteristics of the patient's clinical state or other therapies.
- Definitely Not Related: The adverse event is definitely produced by the patient's clinical state or by other modes of therapy administered to the patient.
- Uncertain Relationship: The adverse event does not fit in any of the above categories.

#### 9.2 Monitoring for Adverse Events:

For patients whose mode of mechanical ventilation is controlled by the trial, investigators will evaluate for the occurrence of adverse events daily as part of the assessment of ventilator modes. Investigators will assess any adverse events that occur for whether the adverse event meets the criteria for recording and reporting outlined below.

#### 9.3 Recording and Reporting Adverse Events:

The following types of adverse events will be recorded and reported:

- Adverse events that are Serious and Definitely Related, Probably or Possibly Related, or of Uncertain Relationship.
- Adverse events that are Unexpected and Definitely Related, Probably or Possibly Related, or of Uncertain Relationship.

Adverse events that do not meet the above criteria will not be recorded or reported. Adverse events that the investigator assesses to meet the above criteria for recording and reporting will be entered into the adverse event electronic case report form in the trial database. The investigator will record a preliminary assessment of each characteristic for the adverse event, including seriousness, unexpectedness, and relatedness. For any adverse event that is **serious AND unexpected**, and definitely related, probably or possibly related, or of uncertain relationship, the investigator will report the adverse event to the principal investigator **within 24 hours** of the investigator becoming aware of the adverse event. For any other adverse event requiring recording and reporting, the investigator will report the adverse event to the principal investigator **within 72 hours** of the investigator becoming aware of the adverse event. The principal investigator will make the final determination regarding each characteristic for the adverse event, including seriousness, expectedness, and relatedness.

For adverse events that meet the above criteria for recording and reporting, the coordinating center will notify the DSMB, the IRB, and the sponsor in accordance with the following reporting plan:

| **Characteristics of the Adverse Event** | **Reporting Period** |
|---|---|
| Fatal or life-threatening (and therefore serious), unexpected, and definitely related, probably or possibility related, or of uncertain relationship. | Report to DSMB and IRB within 7 days after notification of the event. |

| | |
|---|---|
| Serious but non-fatal and non-life-threatening, unexpected, and definitely related, probably or possibly related, or of uncertain relationship. | Report to DSMB and IRB within 7 days of notification of the event. |
| All other adverse events meeting criteria for recording and reporting. | Report to DSMB and IRB as part of continuing review. |

The investigator will distribute the written summary of the DSMB's periodic review of reported adverse events to the IRB in accordance with NIH guidelines (http://grants.nih.gov/grants/guide/notice-files/not99-107.html).

##### 9.4 Clinical Outcomes that may be Exempt from Adverse Event Recording and Reporting:

In this study of critically ill patients at high risk for death and other adverse outcomes due to their underlying critical illness, clinical outcomes, including death and organ dysfunction, will be systematically collected and analyzed for all patients. The primary and exploratory outcomes will be recorded and reported as clinical outcomes and not as adverse events unless treating clinicians or site investigators believe the event is Definitely Related or Probably or Possibly Related to the study intervention or study procedures. This approach – considering death and organ dysfunction as clinical outcomes rather than adverse events and systemically collecting these clinical outcomes for analysis – is common in ICU trials. This approach ensures comprehensive data on death and organ dysfunction for all patients, rather than relying on sporadic adverse event reporting to identify these important events. The following events are examples of study-specific clinical outcomes that would not be recorded and reported as adverse events unless treating clinicians or site investigators believe the event was Definitely Related or Probably or Possibly Related to the study intervention or study procedures:

- Death (all deaths occurring prior to hospital discharge or 28 days will be recorded);
- Organ dysfunction
    - Pulmonary – hypoxemia, hypercarbia, acute hypoxemic respiratory failure, pneumothorax;
    - Cardiac – hypotension, cardiac arrest or shock with or without receipt of vasopressors;
    - Delirium or coma;
- Duration of mechanical ventilation;
- Duration of ICU admission;
- Duration of hospitalization.

Note: A study-specific clinical outcome may also qualify as an adverse event meeting criteria for recording and reporting. For example, a pneumothorax that the investigator considers Definitely Related to ventilator mode would be both recorded as a study-specific clinical outcome and recorded and reported as a Serious and Definitely Related adverse event.

##### 9.5 Unanticipated Problems involving Risks to Subjects or Others:

Investigators must also report to the principal investigator Unanticipated Problems Involving Risks to Subjects or Others ("Unanticipated Problems"), regardless of severity, associated with study procedures

**within 24 hours** of the investigator becoming aware of the Unanticipated Problem. An Unanticipated Problem is defined as any incident, experience, or outcome that meets all of the following criteria:

- Unexpected (in terms of nature, severity, or frequency) given (a) the research procedures that are described in the protocol-related documents, such as the IRB-approved research protocol; and (b) the characteristics of the subject population being studied; AND
- Definitely Related or Probably or Possibly Related to participation in the research (as defined above in the section on characteristics of adverse events); AND
- Suggests that the research places subjects or others at a greater risk of harm (including physical, psychological, economic, or social harm) than was previously known or recognized.

Upon becoming aware of any event that may represent an Unanticipated Problem, the investigator will assess whether the event represents an Unanticipated Problem by applying the criteria described above. If the investigator determines that the event represents an Unanticipated Problem, the investigator will record the Unanticipated Problem in the Unanticipated Problem electronic case report form in the trial database. The investigators will obtain information about the Unanticipated Problem and report the Unanticipated Problem to the DSMB and the IRB within 15 days of becoming aware of the Unanticipated Problem.

#### 10.0 Statistical Considerations

*Sample size justification*.
The purpose of this trial is to assess feasibility of a cluster-randomized, cluster-crossover design. To assess whether compliance can be maintained after cross-over events, we will need to observe two cross-over events to each mode while maintaining an equal number of treatment blocks for each mode. Doing so will require a total of 9 months. Based on the experience of a prior trial in the same ICU,[70] we anticipate an average of 75 patients on mechanical ventilation will be enrolled per month. In 9 months, we expect to enroll 675 patients, of whom 69 will be enrolled during washout periods and excluded from the primary analysis. The resulting 606 patients will be included in the primary analysis.

While the sample size was determined to demonstrate feasibility of the cluster-crossover design, we anticipate this pilot will be the largest randomized trial of ventilator modes to date, and as such, we plan to compare ventilator free days to day 28 (primary outcome) between groups. In a prior cluster-randomized cluster-crossover trial in the same ICU,[70] mechanically ventilated patients experienced a median of 22 VFDs [IQR 0-25 VFDs] and an intra-cluster intra-period correlation of 0.01. With 606 (202 in each group) patients in the primary analysis, a standard deviation in the primary outcome of VFDs of 11 days, and a two-sided alpha of 0.05, the MODE trial will have 80 percent statistical power to detect an absolute difference between groups in the sole primary clinical endpoint of Ventilator Free Days of 3 days.

*Primary Analysis of the Primary Clinical Outcome*
The sole pre-specified primary outcome of ventilator free days will be compared between the three trial groups in an intention-to-treat fashion among all patients enrolled in the trial except those admitted during one of the 3-day washout periods. To estimate the conditional effect of a group assignment on an individual patient given the values of the covariates for that patient, we will use a proportional odds model with independent covariates of group assignment (volume control, pressure control, or adaptive

pressure control) and time. Time (in days) will be treated as a continuous variable with values ranging from 1 (first day of enrollment) to 271 (final day of enrollment) and will be analyzed using restricted cubic splines with multiple knots to allow for non-linearity resulting from seasonality or secular trends. In addition to assessing for an overall group effect within the model, we will estimate the differences between each pair of modes by extracting 95% confidence intervals from the model.

*Sensitivity Analyses of the Primary Clinical Outcomes:*
We will repeat the primary clinical analyses using alternative statistical approaches to comparing the VFDs outcome between groups.

We will repeat the primary clinical outcome analysis with adjustment for pre-specified baseline covariates.

We will repeat the analyses of the primary outcome and feasibility outcomes among all patients enrolled in the trial including those initiated on invasive mechanical ventilation in the study ICU during one of the pre-specified 3-day washout periods.

*Analysis of Effect Modification for the Primary Outcomes.*
We will examine whether pre-specified baseline variables modify the effect of study group on the primary clinical outcome using formal tests of statistical interaction in a proportional odds model. Independent variables will include study group assignment, the potential effect modifier of interest, and the interaction between the two (e.g., study group * history of COPD) and time. Significance will be determined by the P value for the interaction term, with values less than 0.10 considered to suggest a potential interaction and values less than 0.05 considered to confirm an interaction.

We will examine whether the following baseline variables modify the effect of study group on the primary outcome:

- Age (continuous variable);
- Source of admission to the ICU (ED, hospital ward, another ICU in the study hospital, operating room, outside hospital);
- Duration of invasive mechanical ventilation prior to enrollment (0 minutes; 1 to 360 minutes; >360 minutes);
- Indication for intubation (categories are not mutually exclusive)
    - Hypoxemic respiratory failure (yes, no);
    - Hypercarbic respiratory failure (yes, no);
    - Altered mental status or airway protection (yes, no);
- Receipt of supplemental oxygen at place of residence prior to hospital admission (yes, no);
- Chronic Obstructive Pulmonary Disease (yes, no);
- SOFA score at enrollment (continuous variable).

*Analysis of the Exploratory, Feasibility, and Clinical Outcomes*
Each of the exploratory outcomes will be compared between groups in an intention-to-treat fashion in the primary analysis population. Continuous outcomes will be compared between groups in a similar manner as the primary clinical outcome. A logistic model will be used for binary outcomes, a

multinomial model for categorical outcomes, and a proportional odds model for ordinal and continuous outcomes.

*Corrections for multiple testing*
We have pre-specified a single primary clinical outcome, which will have a test of statistical significance for difference between groups. Consistent with recommendations of the Food and Drug Administration and the European Medicines Association, the clinical outcome will be tested using a two-sided p-value with a significance level of 0.05. For all other analyses, emphasis will be placed on the estimate of effect size with 95% confidence intervals, as recommended by the International Committee of Medical Journal Editors, and no corrections for multiple comparisons will be performed.

*Handling of missing data*
The primary outcome is not anticipated to be missing for any patients. Missing data will not be imputed for the primary outcome or any secondary or exploratory outcomes. In adjusted analyses, missing data for covariates will be imputed using multiple imputations.

*Interim Analysis*
In addition to ongoing monitoring of safety throughout the trial, we will plan for the DSMB to conduct a single interim analysis for safety after the ICU has been assigned to each of the three study modes, that is, after enrollment of patients for 3 months. At the interim analysis, the DSMB will evaluate the incidence of the following outcomes across the study groups (volume control, pressure control, or adaptive pressure control):

- Pneumomediastinum or pneumothorax during the course of mechanical ventilation,
- Episodes of hypoxemia while receiving invasive mechanical ventilation: $SpO_2 < 85\%$ for more than 5 minutes,
- In-hospital mortality to study day 28.

The stopping boundary for safety will be met if the P value for the difference between groups is 0.05 or less. Given the design of the study is to demonstrate feasibility and pilot trial procedures rather than demonstrating efficacy, there will be no stopping boundary for efficacy or futility.

#### 11.0 Privacy / Confidentiality Issues

At no time during this study, its analysis, or its publication, will patient identities be revealed in any manner. The minimum necessary data containing patient or provider identities will be collected. Data collected from the medical record and the Mode Modification Sheets will be entered into the secure online database REDCap. Only key study personnel will have access to the REDCap database. Hard copies of the treating clinician Mode Modification Sheet will be stored in a locked room until after the completion of enrollment and data cleaning.

All data will be maintained in the secure online database REDCap until the time of study publication. At the time of publication, a de-identified version of the database will be generated.

#### 12.0 Follow-up and Record Retention

Patients will be followed after enrollment for 28 days or until hospital discharge, whichever occurs first. Data collected from the medical record will be entered into the secure online database REDCap. Hard copies of the treating clinician Mode Modification Sheet will be used for manual data entry into the REDCap database and stored in a locked room until after the completion of enrollment and data cleaning. All data will be maintained in the secure online database REDCap until the time of study publication. Once data are verified and the database is locked, all hard copies of Mode Modification Sheets will be destroyed. At the time of publication, a de-identified version of the database will be generated.

#### 13.0 Data Safety and Monitoring Board (DSMB)

The principal role of the DSMB is to assure the safety of patients in the trial. They will regularly monitor data from this trial, review and assess the performance of its operations, and make recommendations to the steering committee and sponsor with respect to:

- Participant safety and risk/benefit ratio of study procedures and interventions
- Protocol amendments (with specific attention to study population, intervention, and study procedures)
- Adherence to the protocol requirements
- Completeness, quality, and planned analysis of data
- Ancillary study burden on participants and main study
- Possible early termination of the trial because of new external information, safety concerns, or inadequate performance.

The DSMB will consist of members with expertise in critical care medicine, pulmonary medicine, biostatistics, and clinical trials. Appointment of all members is contingent upon the absence of any conflicts of interest. All the members of the DSMB are voting members. The Principal Investigator and unblinded study biostatistician will be responsible for the preparation of all DSMB and adverse event reports. The DSMB will develop a charter and review the protocol and patient notification forms during its first meeting. Subsequent DSMB meetings will be scheduled in accordance with the DSMB Charter with the assistance of the Principal Investigator. The DSMB will have the ability to recommend that the trial end, be modified, or continued unchanged.

**Tracking of Protocol Versions:**

Version 1.0 – Initial Protocol.

**Appendix 1: Ventilator Guidelines**

SUMMARY OF VUMC MICU VENTILATOR PROTOCOL GUIDELINES:

**Ventilator Set Up and Adjustment:**

1. Calculate Predicted Body Weight (PBW):
    a. In males = 50 - 2.3 (height (inches) - 60)
    b. In females = 45.5 + 2.3 (height (inches) - 60)

2. Select Mode and initial default settings:
    a. **Volume Control.** Set Tidal Volume (Vt) to target 6 mL/kg PBW. Set Flow Pattern: 100 or [square-wave icon]; Set T pause (s): 0.
    b. **Pressure Regulated Volume Control.** Set Vt to target 6 mL/kg PBW.
    c. **Pressure Control.** Set PC above PEEP to 15 cm$H_2O$, set inspiratory time to 0.9 s and confirm delivered Vt is appropriate for target 6 mL/kg PBW.

3. Set initial respiratory rate (RR) to approximate baseline Minute Ventilation (12-16 bpm, not > 35 bpm).

4. Set alarms:
    For **VC and PRVC**: High Peak Pressure alarm at 40 cm $H_20$
    For **PC**: Low Minute Ventilation alarm at 2 L/min less than the current Minute Ventilation

5. Adjust Vt and RR to achieve pH and plateau pressure goals below.

**Lung Protective Ventilation Goals – Vt** ≤ 6 mL/kg PBW**, Pplat** < 30 cm$H_20$

**For Pressure Control, where Vt cannot be directly adjusted, adjust PC over PEEP in increments of 2 cm$H_20$ and monitor resulting Vt to achieve the changes outlined below.*

If Vt > 6 mL/kg PBW:
    Reduce Vt by 1 mL/kg at intervals ≤ 2 hours until Vt = 6 mL/kg PBW.
If Pplat > 30 (or PIP > 35):
    Decrease Vt by 1 mL/kg steps to a minimum of 4mL/kg PBW.
If Pplat < 25 and Vt < 6 mL/kg:
    Increase Vt by 1 mL/kg until either Pplat > 25 or Vt = 6 mL/kg PBW.
If Pplat <3 0 and breath stacking occurs:
    May increase Vt in 1 mL/kg increments to maximum of 8 mL/kg PBW.

**Ventilation Goals - pH**: 7.30-7.45

If pH 7.15-7.30:
    Increase RR until pH > 7.30 or PaCO2 < 25 (Max RR = 35).
If RR = 35 and PaCO2 < 25:
    May give $NaHCO_3$, to be managed by primary team.
If pH < 7.15:
    Increase RR to 35.
If pH remains < 7 .15 and $NaHCO_3$ considered or infused:

Vt may be increased until pH >7.15 or Vt = 8 mL/kg PBW (Pplat target may be exceeded).

If pH >7.45:
Decrease RR rate.

**Oxygenation goals - $SpO_2$:** 88-95% or **$PaO_2$:** 55-80 mmHg.

Use incremental $FiO_2$/PEEP combinations below to provide minimum $FiO_2$ and PEEP necessary to achieve $PaO_2$ or $SpO_2$ goals.

| **FiO2** | 0.3-0.4 | 0.4 | 0.5 | 0.5 | 0.6 | 0.7 | 0.7 | 0.7 | 0.8 | 0.9 | 0.9 | 0.9 | 1 |
|---|---|---|---|---|---|---|---|---|---|---|---|---|---|
| **PEEP** | 5 | 8 | 8 | 10 | 10 | 10 | 12 | 14 | 14 | 14 | 16 | 18 | 18-25 |

**Other Settings**

**I:E ratio goal:** 1:1.0 – 1-3.0. Adjust Ti to achieve goal.
If $FiO_2$ = 1.0 and PEEP = 24 $cmH_20$, may adjust I:E to 1:1.
Avoid inverse-ratio ventilation.
**Trigger:** 1.6 L/min by default, can be increased or reduced to improve synchrony
**Tinsp rise (s):** 0.15 s by default, can be increased or decreased to improve ventilation

**Monitoring and Documentation:**

Measure & record Pplat with inspiratory pause of at least 0.5 s, $SpO_2$, Total RR, Vt and pH (if available) at least every 4 hours AND after each change in PEEP, Vt, or PC above PEEP settings.

Once the patient tolerates $FiO_2$ < 0.5 and PEEP < 8 $cmH_2O$, advance to Part II: Spontaneous Breathing Trial Readiness Assessment and Ventilator Discontinuation Guidelines.

**Appendix 2. Ventilator Guidelines Table**

| | Volume Control | Pressure Control | Adaptive Pressure Control |
|---|---|---|---|
| Mode option | Volume Control | Pressure Control | Pressure Regulated Volume Control (PRVC) |
| Tidal Volume target (mL/kg PBW) | 6 | 6 | 6 |
| Tidal Volume range (mL/kg PBW) | 4 - 8 | 4 - 8 | 4 - 8 |
| Initial breath settings | Tidal volume to 6 mL/kg PBW | PC over PEEP to 15 cm H20, Insp Time to 0.9 sec | Tidal volume to 6 mL/kg PBW |
| Achieving target Tidal Volume | Directly adjust tidal volume | Dependent on adjustment of PC over PEEP | Adjust tidal volume directly |
| Plat Pressure target (cm H20) | ≤ 30 | ≤ 30 | ≤ 30 |
| Achieving target Plateau Pressure | Dependent on adjustment of tidal volume | Directly adjust PC over PEEP | Dependent on adjustment of tidal volume |
| Flow | Set directly | Dependent ventilator and patient factors | Dependent ventilator and patient factors |
| Flow Pattern | Square waveform | Decelerating, dynamic | Decelerating, dynamic |
| Arterial pH goal | 7.3 - 7.45 | 7.3 - 7.45 | 7.3 - 7.45 |
| Ventilator rate (breaths/min) | ≤ 35 | ≤ 35 | ≤ 35 |
| Oxygenation goal by PaO2 (mmHg) | 55 - 80 | 55 - 80 | 55 - 80 |
| Oxygenation goal by SpO2 (%) | 88 - 95 | 88 - 95 | 88 - 95 |
| Positive end-expiratory pressure | Set with FiO2-PEEP table | Set with FiO2-PEEP table | Set with FiO2-PEEP table |
| Inspiratory: Expiratory ratio | 1:1 - 1:3 | 1:1 - 1:3 | 1:1 - 1:3 |
| Extubation evaluation | MICU SBT Readiness and Extubation Guidelines | MICU SBT Readiness and Extubation Guidelines | MICU SBT Readiness and Extubation Guidelines |

**Appendix 3: Spontaneous Breathing Trials and Ventilator Discontinuation Guidelines**

**Part I – Spontaneous Breathing Trial Readiness Assessment**

Every morning or at least once daily, patients will undergo screening and trial in a Spontaneous Awakening Trial (SAT) performed by bedside nursing. If the patient passes the SAT, then the patient should be promptly evaluated in a Spontaneous Breathing Trial (SBT) Readiness Assessment as a safety screening:

If all of the following criteria have been present for more than 2 hours, then evaluate the patient for progression to Spontaneous Breathing Trial:
a) $FiO_2 \leq 0.5$ & PEEP ≤ 8 $cmH_2O$
b) Values of both PEEP and $FiO_2 \leq$ values from previous 12 hours.
c) Not receiving neuromuscular blocking agents
d) Patient is exhibiting adequate inspiratory efforts (if not, decrease the ventilator rate to 50% of baseline level for up to 5 minutes to detect inspiratory effort).
e) Systolic arterial pressure > 90 mmHg without significant vasopressor support (< 10 mcg/min of norepinephrine or < 5 mcg/kg/min dopamine or dobutamine will not be considered a vasopressor).

**Part II – Spontaneous Breathing Trial (SBT)**

If criteria a-e above are met, then initiate a trial of at least 30 minutes of spontaneous breathing with FIO2 ≤ 0.5 using any of the following approaches:
1. Pressure support ≤ 5$cmH_2O$, PEEP ≤ 5$cmH_2O$
2. CPAP ≤ 5 $cmH_2O$
3. T-piece
4. Tracheostomy mask

**Monitor for tolerance using the following:**
1. $SpO_2 \geq 88\%$ and / or $PaO_2 \geq 60$ mmHg. $FiO_2$ may be increased to 0.5 if necessary to maintain oxygenation in the target range.
2. Mean spontaneous tidal volume ≥ 4 mL/kg PBW (if measured)
3. Total Respiratory Rate ≤ 35 / min (< 5 min at RR > 35 may be tolerated, example post suctioning)
4. pH ≥ 7.30 (if measured)
5. No respiratory distress (defined as 2 or more of the following):
    a. Heart rate ≥ 120% of the 06:00 rate (≤ 5 min at > 120% may be tolerated)
    b. Marked use of accessory muscles
    c. Abdominal paradoxical breathing
    d. Diaphoresis
    e. Marked subjective dyspnea.

If any of the goals 1-5 are NOT met, revert to previous ventilator settings with Positive End-expiratory Pressure and $FiO_2$ = previous settings and reassess for SBT the next morning.

The clinical team may decide to change mode of support during spontaneous breathing (PS = 5cm$H_2O$, CPAP, tracheostomy mask, or T-piece) at any time.

**Part III – Decision to Remove Ventilatory Support**

For intubated patients, if tolerance criteria for spontaneous breathing trial (1-5 above) are met for at least 30 minutes, continue with unassisted breathing and extubate if clinically indicated under the direction of team physicians. However, the spontaneous breathing trial can continue for up to 120 minutes if tolerance remains in question (in fragile patients).

If any of criteria 1-5 are NOT met during spontaneous breathing (or 120 minutes has passed without clear tolerance), then the ventilator settings that were in use before the attempt to wean will be restored (Positive End-expiratory Pressure and $FiO_2$ = previous settings) and the patient will be reassessed for an SBT the following day.

**Tracking of Protocol Versions:**

Version 1.0 – Initial Protocol (IRB approval 7/21/22)

### Mode Of Ventilation During Critical IllnEss (MODE) Pilot Trial

**Clinical Trial Protocol**

**Version 1.2**

**Principal Investigator**

Kevin Seitz, MD, MSc
*Division of Allergy, Pulmonary, and Critical Care Medicine*
*Department of Medicine*
*Vanderbilt University School of Medicine*

**Co-Investigators**

Jonathan Casey, MD, MSc
Matthew Semler, MD, MSc
Todd Rice, MD, MSc
Edward Qian, MD
Bradley Lloyd, RT-ACCS
*Division of Allergy, Pulmonary, and Critical Care Medicine*
*Department of Medicine*
*Vanderbilt University School of Medicine*

Wesley Self, MD, MPH
*Department of Emergency Medicine*
*Vanderbilt University School of Medicine*

**Table of Contents**

**1.0 Trial Summary 4**

**2.0 Background 6**

**3.0 Aims and Hypotheses 7**

3.1 Study Aims: 7

3.2 Hypotheses: 7

**4.0 Study Description 7**

4.1 Rationale for Cluster-Level Allocation 8

**5.0 Inclusion and Exclusion Criteria 9**

5.1 Inclusion Criteria: 9

5.2 Exclusion Criteria: 9

**6.0 Enrollment / Randomization 9**

6.1 Study Site: 9

6.2 Study Population: 9

6.3 Enrollment: 9

6.4 Consent: 9

6.5 Randomization: 11

**7.0 Study Procedures 11**

7.1 Study Interventions: 11

7.2 Study Co-interventions: 13

7.3 Monitoring Ventilator Mode: 14

7.4 Blinding: 14

7.5 Data Collection: 14

7.6 Outcome Measures: 15

**8.0 Risks and Benefits 16**

**9.0 Safety Monitoring and Adverse Events 16**

9.1 Adverse Event Definitions: 17

9.2 Monitoring for Adverse Events: 18

9.3 Recording and Reporting Adverse Events: 18

9.4 Clinical Outcomes that may be Exempt from Adverse Event Recording and Reporting: 19

9.5 Unanticipated Problems involving Risks to Subjects or Others: 19

**10.0 Statistical Considerations 20**

**11.0 Privacy / Confidentiality Issues 22**

**12.0 Follow-up and Record Retention 22**

**13.0 Data Safety and Monitoring Board (DSMB) 23**

**14.0 References 23**

**Appendix 1: Ventilator Guidelines 28**

**Appendix 2. Ventilator Guidelines Table 30**

**Appendix 3: Spontaneous Breathing Trials and Ventilator Discontinuation Guidelines 31**

#### 1.0 Trial Summary

| | |
|---|---|
| **Title** | Mode Of Ventilation During Critical IllnEss (MODE) Pilot Trial |
| **Background** | Landmark trials in critical care have demonstrated that, among critically ill adults receiving invasive mechanical ventilation, the use of low tidal volumes and low airway pressures prevents lung injury and improves patient outcomes. Limited evidence, however, informs the best method of mechanical ventilation to achieve these targets. To provide mechanical ventilation, clinicians must choose between modes of ventilation that directly control tidal volumes ("volume control"), modes that directly control the inspiratory airway pressure ("pressure control"), and modes that are hybrids ("adaptive pressure control"). Whether the choice of the mode used to target low tidal volumes and low inspiratory plateau pressures affects clinical outcomes for critically ill adults receiving mechanical ventilation is unknown. All three modes of mechanical ventilation are commonly used in clinical practice. A large, multicenter randomized trial comparing available modes of mechanical ventilation is needed to understand the effect of each mode on clinical outcomes. We will conduct a pilot trial evaluating the feasibility of comparing modes for mechanically ventilated ICU patients. |
| **Study Design** | Prospective, unblinded, cluster-randomized, multiple-crossover pilot trial |
| **Trial Groups** | 1. Volume Control group<br>2. Pressure Control group<br>3. Adaptive Pressure Control group |
| **Inclusion Criteria** | 1. Age ≥ 18 years<br>2. Receiving mechanical ventilation through an endotracheal tube or tracheostomy<br>3. Admitted to the study ICU |
| **Exclusion Criteria** | 1. Patient is pregnant<br>2. Patient is a prisoner<br>3. Patient was receiving invasive mechanical ventilation at place of residence prior to hospital admission<br>4. Patient is receiving extracorporeal membrane oxygenation at the time of admission to the study ICU |
| **Randomization** | In this cluster-randomized cluster-crossover trial, the entire study ICU will be assigned to a mode of ventilation (volume control, pressure control, or adaptive pressure control) and will switch between modes every month in an order determined by computer-generated randomization using permuted blocks of 3. |
| **Primary Outcome** | Ventilator-free days to day 28 after enrollment |
| **Exploratory Outcomes** | Feasibility Outcomes<br>• Exposure to assigned study mode in first 3 days: Proportion of time in the assigned mode while receiving invasive mechanical ventilation in the study ICU between enrollment and 72 hours after enrollment |

| | |
|---|---|
| | • Adherence to study mode in first 3 days: Proportion of time in the assigned mode while receiving invasive mechanical ventilation in the study ICU with a mandatory mode between enrollment and 72 hours after enrollment (excluding time spent in spontaneous modes)<br>• Time from enrollment to initiation of assigned mode of mechanical ventilation<br>• Receipt of a "Mode Modification Sheet" completed by treating clinicians<br>Exploratory Efficacy and Safety Outcomes<br>• Median exhaled tidal volume (mL/kg predicted body weight) on each study day<br>• Exhaled tidal volumes above target range: Proportion of recorded breaths with exhaled tidal volume values above the target range (>8mL/kg predicted body weight) on each study day<br>• Hypoxemia during mechanical ventilation: Episodes of hypoxemia during mechanical ventilation: SpO2 < 85% for more than 5 minutes<br>• Severe acidemia during mechanical ventilation: Episodes of severe acidemia during mechanical ventilation: pH < 7.1 on blood gas<br>• Number of blood gas laboratory tests per day while receiving mechanical ventilation<br>• Pneumomediastinum or pneumothorax during course of mechanical ventilation<br>• SOFA score daily on the first 7 study days<br>• Delirium and coma-free days to day 28<br>Exploratory Clinical Outcomes:<br>• ICU-free days to study day 28<br>• Hospital-free days to study day 28<br>• In-hospital mortality to study day 28 |
| **Analysis** | The primary analysis of the primary outcome (ventilator-free days) will be conducted in an intention-to-treat fashion using a proportional odds model with independent covariates of group assignment (volume control, pressure control, or adaptive pressure control) and time since beginning enrollment (in days). |
| **Sample Size** | The trial will be conducted for a total of 9 months as this is the duration needed to evaluate the feasibility of the cluster-randomized multiple-crossover design. Based on data from a prior trial in the same ICU, we expect to enroll 675 patients, of whom 69 will be enrolled during washout periods and 606 patients will be included in the primary analysis. |
| **Duration** | 9 months |

#### 2.0 Background

Approximately 2-3 million critically ill adults receive invasive mechanical ventilation in the ICU each year.[1–3] Mechanical ventilation provides support for ventilation and gas exchange while patients recover from the underlying causes of their respiratory failure. Mechanical ventilation itself, however, may injure the lungs, diaphragm, and other organs,[4,5] and in-hospital mortality for mechanically ventilated ICU patients remains 25-35%.[6]

For patients on mechanical ventilation, tidal volume and inspiratory airway pressures are directly related. Increasing tidal volumes increases airway pressures and vice versa. The ventilator can be set by clinicians to deliver breaths in one of two ways: either by controlling the volume ("volume control") or the pressure ("pressure control") administered during the inspiratory phase of the breath. By controlling one factor, each mode provides indirect control over the other, as a dependent variable that varies proportionally based on the patient's lung physiology.[7] Modern ventilators also offer hybrid modes such as adaptive pressure control, allowing clinicians to set a target volume for which the ventilator titrates and delivers the pressure controlled breath that is expected to be adequate based on the results of previous breaths (e.g. Pressure Regulated Volume Control). The mode of mechanical ventilation must be set for each of 2-3 million ICU patients who receive invasive mechanical ventilation. Yet, whether choice of mode affects outcomes remains uncertain. [8–10]

Landmark clinical trials in critically ill patients have demonstrated that using smaller tidal volumes and plateau pressures (Low Tidal Volume Ventilation) reduces lung injury and improves patient outcomes, both for patients with ARDS[11] and for patients without ARDS.[12] Each of the commonly used modes of mechanical ventilation (volume control, pressure control, adaptive pressure control) can achieve these low tidal volume and low plateau pressure targets – but minimal data exist to guide the choice of which ventilator mode should be used to achieve these targets. Current guidelines recommend delivering low tidal volumes and low plateau pressures to all patients on mechanical ventilation and make no recommendations regarding the mode of mechanical ventilation used to achieve these targets. [13–18] Volume control, pressure control, and adaptive pressure control can each be used to accomplish adequate gas exchange in critically ill patients, but they differ in how they control tidal volumes and inspiratory pressures and synchronize with patient efforts to breathe.[19,20]

Only two clinical trials have directly compared volume control and pressure control as the modes of mechanical ventilation for critically ill adults. The trials were small and lacked adequate power to evaluate for clinically significant difference in patient-important outcomes.[21,22] Further, they were performed more than 20 years ago, prior to many modern practices of critical care including low tidal volume ventilation. Evidence comparing adaptive modes to volume or pressure control are similarly limited to small trials with mixed results.[23,24] Reviews and meta-analyses of small and heterogenous studies of physiology have also not demonstrated clear superiority of volume control, pressure control, or adaptive pressure control.[8,9,20]

All three modes are in common use in large, international cohorts of ARDS and non-ARDS patients, with a slight increase in the use of pressure modes over time.[12,25,26] Pressure control modes are the most common modes in international cohorts.[12,26–28] While ventilator settings for critically ill patients in the United States are not as well described, the most recent data on patients in the US receiving contemporary critical care, from a large multi-center cohort of moderate-severe ARDS patients, demonstrated that on the first day of mechanical ventilation with ARDS, volume control was used in approximately half of patients, with most of the remainder receiving pressure regulated modes in either pressure control or adaptive pressure control.[29] Among all patients receiving mechanical ventilation in the Vanderbilt Medical ICU in 2021, 35% of breaths were in volume control, 31% were in pressure control, and 35% were in adaptive pressure control.

Because millions of critically ill adults receive invasive mechanical ventilation each year, a mode of ventilation must be selected for every patient, three modes are used commonly in current practice, and no high-quality data inform the effect of mode of ventilation on patient outcomes, a large, randomized trial is needed to inform the optimal choice of ventilator mode for mechanically ventilated ICU patients. Before such a trial can be conducted, however, data are needed to demonstrate the feasibility of maintaining adherence to ventilator modes in a pragmatic trial.[30–32]

#### 3.0 Aims and Hypotheses

##### 3.1 Study Aims:

- **Feasibility Aim:** To assess the feasibility of a cluster-randomized, multiple-crossover trial comparing volume control, pressure control, and adaptive pressure control modes.

- **Clinical Aim:** To evaluate the effect of the same ventilator mode intervention on days alive and free of invasive mechanical ventilation in the same population of mechanically ventilated critically ill patients.

##### 3.2 Hypotheses:

- **Feasibility Hypothesis:** Comparing volume control, pressure control, and adaptive pressure control ventilation among mechanically ventilated ICU patients will be feasible.

- **Clinical Hypothesis:** Use of an adaptive pressure control mode for mechanically ventilated ICU patients will result in more days alive and free of invasive mechanical ventilation than use of either a volume control or pressure control mode.

#### 4.0 Study Description

To address the aims outlined above, we propose the Mode Of Ventilation During Critical IllnEss (MODE) Pilot Trial. This trial will be a prospective, single-center, unblinded, cluster-randomized, multiple-crossover trial conducted in the medical ICU at Vanderbilt University Medical Center. During the 9 months of enrollment in the MODE trial, the entire medical ICU will be assigned to a single ventilator mode with the assigned mode alternating between volume control, pressure control, and adaptive pressure control modes every month in a randomly generated sequence. All patients who receive invasive mechanical ventilation and meet inclusion criteria without meeting exclusion criteria will be enrolled at the initiation of mechanical ventilation. The trial will control only the ventilator mode during controlled mechanical ventilation. All other aspects of clinical care for patients will remain at the discretion of treating clinicians.

#### 4.1 Rationale for Cluster-Level Allocation:

Group assignment in the MODE trial occurs at the level of the ICU (cluster) for multiple reasons. In routine clinical care in the study ICU, 2 to 4 respiratory therapists set the ventilator mode and adjust settings to maintain gas exchange and patient comfort for all mechanically ventilated adults, with input from nurses and physicians. The management of mechanical ventilation (selection of fraction of inspired oxygen, titration of positive end-expiratory pressure, screening for and performance of spontaneous breathing trials) is governed by unit-wide protocols implemented by the 2 to 4 respiratory therapists for all patients in the unit. Assigning the entire unit to a single ventilator mode both emulates the way mechanical ventilation is managed during clinical care and limits contamination that might result from a respiratory therapist managing multiple patients assigned to different modes of mechanical ventilation.

Additionally, mechanical ventilation can damage lungs even after brief periods of ventilation, and the initial settings have a unique significance. For example, during the minutes-to-hours of temporary mechanical ventilation for surgical procedures, harmful ventilator settings can affect organ function and clinical outcomes.[34] In critically ill patients, the initial hours of invasive mechanical ventilation are also the period with the highest risk of worsening lung injury, dyssynchrony, increased work of breathing, and hemodynamic compromise. Prior research has demonstrated that [1] the very first ventilator settings at the time the patient is being initiated on invasive mechanical ventilation in the ICU are associated with differences in mortality[35,36] and [2] the association between ventilator settings and mortality is larger for earlier settings compared to later settings.[33] The ventilator protocol used to set the ventilator mode in the study ICU begins immediately at the time of initiation of mechanical ventilation. Enrollment immediately after initiation of invasive mechanical ventilation minimizes pre-study exposure to other modes and ventilator settings, and it facilitates on-study separation between groups.

Recognizing the importance of initial ventilator settings, three recent clinical trials have examined other ventilator settings (i.e. end-expiratory pressure, tidal volume, and $SpO_2$ target) by attempting to enroll patients soon after the initiation of mechanical ventilation in the ICU. [12,37,38] In these patient-level, parallel-group trials, however, the logistical challenges of performing screening, enrollment, randomization, and study group assignment resulted in a significant gap between initiation of invasive mechanical ventilation and delivery of the respective trial interventions while also leading to the exclusion of 60-67% of eligible patients – raising concern for systematic exclusion of important patient groups (e.g. patients with higher acuity of illness). In MODE, group assignment at the cluster level allows enrollment immediately on initiation of invasive mechanical ventilation in the ICU. This approach emulates the manner in which ventilator settings are managed in practice, precludes systematic exclusion of important patient groups, decreases pre-study exposure to harmful ventilator settings, and facilitates early separation in the receipt of ventilator modes between groups.

#### 5.0 Inclusion and Exclusion Criteria

##### 5.1 Inclusion Criteria:

1. Age ≥ 18 years
2. Receiving mechanical ventilation through an endotracheal tube or tracheostomy
3. Admitted to the study ICU.

##### 5.2 Exclusion Criteria:

1. Patient is pregnant
2. Patient is a prisoner
3. Patient receiving invasive mechanical ventilation at place of residence prior to hospital admission
4. Patient receiving extracorporeal membrane oxygenation at the time of admission to the study ICU.

#### 6.0 Enrollment / Randomization

##### 6.1 Study Site:

- Medical Intensive Care Unit at Vanderbilt University Medical Center

##### 6.2 Study Population:

All adults located in the study ICU for whom the treating clinicians have decided invasive mechanical ventilation is required will be enrolled unless meeting exclusion criteria. Patients will be included regardless of age, gender, race, or other clinical factors.

##### 6.3 Enrollment:

All adult patients who do not meet exclusion criteria will be enrolled immediately upon receipt of invasive mechanical ventilation in the study ICU.

##### 6.4 Consent:

Volume control, pressure control, and adaptive pressure control are all common approaches to controlled mechanical ventilation for critically ill adults. All represent standard-of-care treatments in current clinical practice. Results from prior clinical trials do not demonstrate superiority of one approach over the other. Current clinical guidelines do not recommend any specific mode of mechanical ventilation.[14,16–18] As a result, significant variation exists in the use of volume control, pressure control, and adaptive pressure control for patients in routine clinical care with ARDS[29] and those without ARDS.[26] In the study ICU, all three modes are currently in use in routine care. Of all patients on mechanical ventilation in the study ICU in 2021, 35% of mandatory breaths were in volume control mode, 31% were in pressure control mode, and 35% were in adaptive pressure control. This trial will only enroll patients who would already receive continuous mandatory ventilation with one of these three modes as part of their routine clinical care. When the primary medical team feels that the patient needs care with a specific ventilator mode different from what is assigned by the study, the team can alter the ventilator mode for the patient and use the mode that they believe is the correct mode for the patient. Only patients for whom the primary team feels that the ventilator mode assigned by the study is acceptable will continue with that assigned ventilator mode. We will request a waiver of informed consent because the study involves minimal risk and obtaining informed consent would be impracticable.

Participation in this study involves minimal risk because:

- The three interventions being compared are commonly used in routine clinical care in the study ICU;
- All are interventions to which the patient could be exposed even if not participating in the study (all patients receiving the initiation of mechanical ventilation receive one of these modes of continuous mandatory ventilation);
- No established differences in risk and benefits are known to exist between the studied approaches to mechanical ventilation based on the currently available data; and
- The trial only determines the mode of ventilation when treating clinicians feel all three modes would be consistent with optimal care for the individual patient – otherwise treating clinicians select the mode of ventilation via a Mode Modification Sheet.

The MODE trial is designed as a cluster-randomized multiple-crossover trial to ensure that the mode selected at the time of initiation of mechanical ventilation is consistent with trial group assignment, preventing contamination between groups and capturing the period of mechanical ventilation in which ventilator settings have the strongest association with outcomes.

Obtaining informed consent before initiation of mechanical ventilation in the study ICU would be impracticable because:

- **The expected medical condition of patients at the time of initiation invasive mechanical ventilation in the ICU is critical.** All patients will be critically ill and receiving continuous intravenous sedation at the time of enrollment. Thus, all patients eligible for MODE will not have the capacity to provide informed consent. Further, data from prior trials in the same patient population and setting demonstrate that prior to intubation and initiation of mechanical ventilation, approximately 70% of patients eligible for the MODE trial will be experiencing encephalopathy (altered mental status) due to their illness. The anticipated median Glasgow coma scale score will be 11 (equivalent to moderate brain injury). Among the minority of patients whose level of consciousness is not impaired, 45-55% will be experiencing acute delirium. Further, family members or legally authorized representatives (LAR) are frequently unavailable when critically ill patients undergo intubation in the ED or ICU.
- **The intervention is delivered by unit-level protocols**. Mechanical ventilator settings and titrations are performed by respiratory therapists using unit-level protocols. In this cluster-randomized trial, the entire unit will be assigned to the same mode of mechanical ventilation for each 1-month block. Obtaining informed consent from every eligible patient in the ICU prior to emergency tracheal intubation and initiation of mechanical ventilation would be impracticable.

Because the study involves minimal risk, the study would not adversely affect the welfare or privacy rights of the participant, and obtaining informed consent would be impracticable, we will request a waiver of informed consent. Numerous previous randomized trials comparing two standards of care for interventions such as emergency intubation and methods of respiratory support during critical illness have been completed with waiver of informed consent.[39–47]

#### 6.5 Randomization:

During each one-month block of the study, the ICU will be assigned to either volume control, pressure control, or adaptive pressure control. The order of the study group assignments will be generated by computerized randomization using permuted blocks of 3 to minimize the impact of seasonal variation. The last 3 days of each block will be a washout period during which the ICU will continue to use the assigned mode, but new patients will not be included in the primary analysis. The 3-day washout period will limit the number of patients who experience a 'crossover' from one assigned mode to another while receiving a mandatory mode of mechanical ventilation.

#### 7.0 Study Procedures

#### 7.1 Study Interventions:

**Ventilator Mode:**

**Volume Control Group:** Patients assigned to the volume control group will receive volume control as the initial mode of ventilation and whenever they are receiving continuous mandatory invasive mechanical ventilation.

**Pressure Control Group:** Patients assigned to the pressure control group will receive pressure control as the initial mode of ventilation and whenever they are receiving continuous mandatory invasive mechanical ventilation.

**Adaptive Pressure Control Group:** Patients assigned to the adaptive pressure control group will receive adaptive pressure control as the initial mode of ventilation and whenever they are receiving continuous mandatory invasive mechanical ventilation.

Characteristics of each of the three modes of ventilation are presented in Table 1.

**Table 1. Characteristics of the modes of mechanical ventilation.**

| Group Assignment | Volume Control | Pressure Control | Adaptive Pressure Control |
|---|---|---|---|
| Ventilator Mode Designation | *Volume Control* | *Pressure Control* | *Pressure Regulated Volume Control* |
| Control Variable | Volume | Pressure | Pressure |
| Breath Sequence | Continuous mandatory ventilation | Continuous mandatory ventilation | Continuous mandatory ventilation |
| Targeting Scheme | Set-point (clinician sets target volume) | Set-point (clinician sets target pressure) | Adaptive (clinician sets target volume and ventilator titrates pressure to achieve target volume) |

*Ventilator Settings and Titration*:
In the study ICU and the United States generally, respiratory therapists have primary responsibility for determining the initial settings for invasive mechanical ventilation and titrating these settings for clinical goals (e.g., $SaO_2$, $pCO_2$, target tidal volume, target plateau pressure, etc.). To set and titrate mechanical ventilator settings, respiratory therapists use clinical protocols jointly developed by respiratory therapy and physician leaders. For patients enrolled in the MODE trial, respiratory therapists will use preexisting clinical protocols for volume control, pressure control, and adaptive pressure control in use in the study unit (Appendix 1-3).

*Initiation of Mechanical Ventilation:* All mechanically ventilated patients being initiated on invasive mechanical ventilation in an ICU initially receive a "mandatory" mode of ventilation (e.g., volume control, pressure control, or adaptive pressure control) immediately following intubation. Mandatory modes are necessary for patients being initiated on invasive mechanical ventilation in an ICU, because patients nearly uniformly received sedation or neuromuscular blockade for placement of the endotracheal tube and do not have the spontaneous respiratory effort required to allow a "spontaneous" mode of ventilation. When respiratory therapists select the initial ventilator settings for a patient being initiated on invasive mechanical ventilation in the ICU, they will select the assigned mode of ventilation (volume control, pressure control, or adaptive pressure control). As prior studies have demonstrated that ventilator settings early in patients' course of invasive mechanical ventilation may affect clinical outcomes,[33] ensuring the first set of ventilator settings includes the mode of ventilation assigned by the study will prevent contamination with non-assignment ventilator modes and ensure delivery of the study intervention during a key period of patients' critical illness.

*Maintenance of Mechanical Ventilation:*
Study protocol will determine the ventilator mode from enrollment until the first of: (1) extubation from mechanical ventilation, (2) transfer out of the study ICU, (3) completion of a ventilator Mode Modification Sheet by treating clinicians, or (4) end of the one-month study block. If a patient is enrolled, extubated, and re-intubated during the same study block, the study protocol will again determine the ventilator mode until they meet one of the aforementioned criteria. Study protocol does not determine the ventilator mode during time-periods in which the patient is not receiving invasive

mechanical ventilation, is not receiving a continuous mandatory mode of ventilation (e.g., when patients who are no longer deeply sedated are weaned to pressure support ventilation), is not physically located in the study ICU (e.g., during transport), or is undergoing an invasive procedure (e.g., bronchoscopy).

*Modification of Ventilator Mode:* At any time, any member of the clinical team (respiratory therapist, nurse, nurse practitioner, or physician) may use any mode of mechanical ventilation if determined to be needed for the optimal treatment of any specific patient. If modifying the mode of ventilation is necessary, the date, time, and reason for modification of the mode will be prospectively recorded.

Details of each mode modification will be collected and monitored on a Mode Modification Sheet. In our prior cluster-crossover trial using the same approach to modification of therapy assignment, treating clinicians exercised the ability to modify the assigned therapy for approximately 5% of patients.[39]

Anticipated examples of conditions for which treating clinicians may elect to override the assigned mode include:

- Refractory hypoxemia
- Peak pressures persistently high (>40 cm $H_20$)
- Asynchrony or dyssynchrony not amenable to changes within the assigned mode of ventilation
- Excessive work of breathing
- Barotrauma (pneumothorax or pneumomediastinum)
- Inability to limit tidal volumes delivered
- Intrinsic PEEP
- Other

##### 7.2 Study Co-interventions:

*Ventilator Adjustment:* For all patients regardless of group assignment, existing clinical ventilator protocols will be used to target a tidal volume of 6 mL/kg PBW and a plateau pressure <30 cm $H_20$, while maintaining a respiratory rate less than 35 and pH of at least greater than 7.15, with a target of 7.3 to 7.45. (Appendix 1-3) Respiratory therapists will titrate volume or pressure settings according to the patient's ventilator mode to achieve these parameters and manage asynchrony and patient discomfort if it occurs.

*Liberation from Mechanical Ventilation:* Each day of mechanical ventilation, all patients in the study ICU are assessed for safety of a spontaneous awakening trial (SAT) and spontaneous breathing trial (SBT)[48] using the SAT and SBT safety criteria from the Awakening and Breathing Controlled (ABC) trial.[49] These criteria relate to the non-respiratory clinical status of the patient and the PEEP and FiO2 settings which will be managed similarly across groups. As such, the SAT and SBT procedures will be handled in the same way for each patient, regardless of study group assignment. Definitions of SAT and SBT failure and the ventilator settings and duration of the SBT will be those used in the ABC trial and used in routine clinical care in the study ICU. For patients who have passed an SAT and SBT, the decision to discontinue invasive mechanical ventilation will be made by the treating clinicians.

*Fraction of Inspired Oxygen ($FiO_2$) and Positive End-Expiratory Pressure (PEEP)*: Institutional protocols in the study ICU will ensure that mechanically ventilated patients in MODE on each study mode receive $FiO_2$ and PEEP titration according to the ARDSNet Lower PEEP/higher $FiO_2$ table (except for patients with severe ARDS, for whom the higher PEEP/lower $FiO_2$ table is applied)[50,51]

_Sedation and Analgesics_: Institutional protocols in the study ICU will ensure that mechanically ventilated patients in MODE on each study mode receive similar management of pain, agitation, and delirium[52] targeting Critical Care Pain Observation Tool (CPOT)[53], Richmond Agitation-Sedation Scale (RASS)[54,55], and Confusion Assessment Method for the ICU (CAM-ICU) scores.[56,57]

##### 7.3 Monitoring Ventilator Mode:

For all mechanically ventilated patients in the study ICU, ventilators display settings such as mode and measurements such as observed tidal volume on a digital display at the patient's bedside in real time. For critical measurements such as respiratory rate, minute ventilation, and peak pressures, audible alarms from the ventilator alert clinicians to potentially dangerous situations. These settings and measurements are sampled and uploaded automatically into the EMR frequently and are stored in the data warehouse. This will allow for much more accurate assessment of the ventilator modes that patients receive over the course of the day compared to prior trials which documented ventilator mode once or twice daily.[58,59]

***Feedback on Ventilator Mode Adherence:*** During the study, study personnel will monitor compliance with the assigned ventilator mode. Study personnel will remotely monitor ventilator modes up to four times daily from 4AM through 10PM Monday through Friday and during a convenience sample of weekend hours to provide feedback to treating clinicians and explore reasons why patients are not receiving the assigned mode of mechanical ventilation,

Study personnel will attend respiratory therapy group meetings, nursing unit board meetings, and ICU physician leadership meetings to educate staff about the study, solicit safety concerns and adverse events, and identify and address barriers to ventilator mode assignment compliance. This approach to daily monitoring and intermittent feedback to clinical personnel successfully achieved 80% compliance with the assigned intervention in a prior cluster-crossover trial of ventilator settings in the same ICU.[39]

##### 7.4 Blinding:

Similar to prior ventilator mode studies among critically ill adults, patients and clinicians will not be blinded to study group assignment.[21,58,59] Observer bias will be minimized by use of objective endpoints collected and automated data extraction from the EHR (see *Data Collection* below).

##### 7.5 Data Collection:

The MODE trial will primarily use structured data collected in routine clinical care, exported daily from the institution's EHR into an Enterprise Data Warehouse, along with data from the patient registration, billing, and laboratory clinical information systems. We have previously validated the quality of this method of data collection against the reference standard of two-physician manual chart review,[60] and the planned approach to electronic dataset generation for the MODE trial has already been successfully employed for the conduct of four prior pragmatic trials.[39,61–63]

**Electronically extracted data elements will include:**
*Enrollment (Day 0):* date and time of clinical events (hospital admission, ICU admission, intubation); source of admission to study ICU; age; sex; race; ethnicity; height, weight;[64] SOFA score;[65] Glasgow Coma Scale score;[66] Elixhauser Comorbidity Index;[67] history of present illness; vital signs (temperature, heart rate, systolic blood pressure, diastolic blood pressure, $SpO_2$); mechanical ventilator settings (mode, set and exhaled tidal volumes, set and actual respiratory rate, minute ventilation, set and total positive end-expiratory pressure, peak pressure, plateau pressure, inspiratory time, flow pattern, $FiO_2$); serum laboratory values (white blood cell count, hemoglobin, platelet count, sodium, potassium, bicarbonate, creatinine, bilirubin, alanine aminotransferase, aspartate aminotransferase, lactate, arterial and venous pH, $PaCO_2$,$PaO_2$, $SaO_2$).

*Daily On-Study (Days 0-28):* Vital signs, ventilator settings, screening for and performance of spontaneous awakening trials and spontaneous breathing trials, and serum laboratory values (as above); Mean daily dose of sedative infusions (propofol, dexmedetomidine, morphine, fentanyl, and midazolam); CAM status; RASS score; Daily completion of each element from ICU Liberation Bundle (A-F); Beginning and end time of Spontaneous Breathing Trials; vasopressor and inotrope infusions (norepinephrine, epinephrine, vasopressin, phenylephrine, dobutamine, angiotensin-2, dopamine); "Mode Modification Sheet" (use, time, reason); Characteristics of treating clinicians (respiratory therapist, attending, pulmonary fellow, registered nurse) providing direct care; Use of therapies for refractory hypoxemia (prone positioning, ECMO, neuromuscular blockade, inhaled pulmonary dilators); SOFA score; ARDS criteria by the Berlin Definition;[68] Stage II or greater AKI by KDIGO criteria;[69] cardiac arrest, pneumothorax or pneumomediastinum.

Termination (Days 0-28): Vital status at 28 days; time of liberation from invasive mechanical ventilation; receipt and duration of vasopressors; receipt and duration of renal replacement therapy; tracheostomy; duration of ICU and hospital admission.

#### 7.6 Outcome Measures:

**Primary Outcome:**
The primary outcome is ventilator-free days (VFDs) to day 28 after enrollment.

VFDs will be defined as the number of calendar days alive and free of invasive mechanical ventilation from the final receipt of invasive mechanical ventilation through 28 days after enrollment. Outcome ascertainment will cease at the time of hospital discharge or 28 days after enrollment, whichever occurs first. Receipt of invasive mechanical ventilation will be considered to end when patients undergo the final tracheal extubation or disconnection of the ventilator from a tracheostomy tube between enrollment (day 1) and day 28 after enrollment. Patients who die prior to discharge on or before day 28 will receive zero VFDs. Patients whose final receipt of invasive mechanical ventilation occurs on the day of enrollment and survive to day 28 will receive 27 VFDs. Patients who continue to receive invasive mechanical ventilation at day 28 will receive zero VFDs. Patients who are discharged from the hospital prior to day 28 and are receiving invasive mechanical ventilation at the time of discharge will receive zero VFDs. Patients who are removed from invasive mechanical ventilation and are discharged from the hospital without invasive mechanical ventilation prior to 28 days will be assumed to remain alive and free of invasive mechanical ventilation between hospital discharge and day 28. For patients who are removed from invasive mechanical ventilation, return to invasive mechanical ventilation, and are subsequently removed again from invasive mechanical ventilation prior to day 28, VFDs will be counted from the final receipt of invasive mechanical ventilation prior to day 28.

**Feasibility Outcomes**

- Exposure to assigned study mode in first 3 days: proportion of time in the assigned mode while receiving invasive mechanical ventilation in the study ICU between enrollment and 72 hours after enrollment.
- Adherence to study mode in first 3 days: proportion of time in the assigned mode while receiving invasive mechanical ventilation in the study ICU with a mandatory mode between enrollment and 72 hours after enrollment (excluding time spent in spontaneous modes).
- Time from enrollment to initiation of assigned mode of mechanical ventilation
- Receipt of a "Mode Modification Sheet" completed by treating clinicians

**Exploratory Efficacy and Safety Outcomes**

- Median exhaled tidal volume (mL/kg predicted body weight (PBW)) on each study day
- Exhaled tidal volumes above target range: proportion of recorded breaths with exhaled tidal volume values above the target range (> 8mL/kg PBW) on each study day
- Hypoxemia during mechanical ventilation: episodes of hypoxemia during mechanical ventilation: $SpO_2$ < 85% for more than 5 minutes
- Severe acidemia during mechanical ventilation: Episodes of severe acidemia during mechanical ventilation: pH < 7.1 on blood gas
- Number of blood gas laboratory tests per day while receiving mechanical ventilation
- Pneumomediastinum or pneumothorax during course of mechanical ventilation
- SOFA score daily on the first 7 study days
- Delirium and coma-free days to day 28

**Exploratory Clinical Outcomes:**

- ICU-free days to study day 28
- Hospital-free days to study day 28
- In-hospital mortality to study day 28

**8.0 Risks and Benefits**

Although no risks are currently known to differ between mechanical ventilation with volume control, pressure control, and adaptive pressure control (all standard-of-care approaches in currently clinical care), it is possible that the results of the MODE trial will ultimately demonstrate a difference between the approaches in the risk of ventilator-free days, hypoxemia, delirium and coma free days, or another outcome. The benefits of this research will include understanding the effect of choice of ventilator mode on outcomes for the 2-3 million adults per year who receive invasive ventilation.

**9.0 Safety Monitoring and Adverse Events**

Assuring patient safety is an essential component of this protocol. Mechanical ventilation with volume control, pressure control, and adaptive pressure control are all standard-of-care interventions that have been use in clinical practice for decades with an established safety profile. This protocol further ensures safety of its participants through:

a. Exclusion criteria designed to prevent enrollment of patients likely to experience adverse events from pressure control, volume control, or adaptive pressure control settings;
b. Permitting treating clinicians to change the ventilator mode at any time for any patient in whom they feel a ventilator mode different from the assigned mode is required for the safe treatment of the patient;
c. Systematic collection of outcomes relevant to the safety of ventilator mode selection;
d. Structured monitoring, assessment, recording, and reporting of adverse events.

##### 9.1 Adverse Event Definitions:

**Adverse Event –** An adverse event will be defined as any untoward or unfavorable medical occurrence in a human subject temporally associated with the subject's participation in the research, whether or not considered related to the subject's participation in the research. Adverse events will be classified according to the following characteristics:

- **Seriousness** – An adverse event will be considered "serious" if it:
  - Results in death;
  - Is life-threatening (defined as placing the patient at immediate risk of death);
  - Results in inpatient hospitalization or prolongation of existing hospitalization;
  - Results in a persistent or significant disability or incapacity;
  - Results in a congenital anomaly or birth defect; or
  - Based upon appropriate medical judgment, may jeopardize the patient's health and may require medical or surgical intervention to prevent one of the other outcomes listed in this definition.

- **Unexpectedness** – An adverse event will be considered "unexpected" if the nature, severity, or frequency is neither consistent with:
  - The known or foreseeable risk of adverse events associated with the procedures involved in the research that are described in the protocol-related documents, such as the IRB-approved research protocol; nor
  - The expected natural progression of any underlying disease, disorder, or condition of the subject experiencing the adverse event and the subject's predisposing risk factor profile for the adverse event.

- **Relatedness** – The strength of the relationship of an adverse event to a study intervention or study procedure will be defined as follows:
  - Definitely Related: The adverse event follows (1) a reasonable, temporal sequence from a study procedure AND (2) cannot be explained by the known characteristics of the patient's clinical state or other therapies AND (3) evaluation of the patient's clinical state indicates to the investigator that the experience is definitely related to study procedures.
  - Probably or Possibly Related: The adverse event meets some but not all of the above

criteria for “Definitely Related”.
- Probably Not Related: The adverse event occurred while the patient was on the study but can reasonably be explained by the known characteristics of the patient’s clinical state or other therapies.
- Definitely Not Related: The adverse event is definitely produced by the patient’s clinical state or by other modes of therapy administered to the patient.
- Uncertain Relationship: The adverse event does not fit in any of the above categories.

#### 9.2 Monitoring for Adverse Events:

For patients whose mode of mechanical ventilation is controlled by the trial, investigators will evaluate for the occurrence of adverse events daily as part of the assessment of ventilator modes. Investigators will assess any adverse events that occur for whether the adverse event meets the criteria for recording and reporting outlined below.

#### 9.3 Recording and Reporting Adverse Events:

The following types of adverse events will be recorded and reported:
- Adverse events that are Serious and Definitely Related, Probably or Possibly Related, or of Uncertain Relationship.
- Adverse events that are Unexpected and Definitely Related, Probably or Possibly Related, or of Uncertain Relationship.

Adverse events that do not meet the above criteria will not be recorded or reported. Adverse events that the investigator assesses to meet the above criteria for recording and reporting will be entered into the adverse event electronic case report form in the trial database. The investigator will record a preliminary assessment of each characteristic for the adverse event, including seriousness, unexpectedness, and relatedness. For any adverse event that is **serious AND unexpected**, and definitely related, probably or possibly related, or of uncertain relationship, the investigator will report the adverse event to the principal investigator **within 24 hours** of the investigator becoming aware of the adverse event. For any other adverse event requiring recording and reporting, the investigator will report the adverse event to the principal investigator **within 72 hours** of the investigator becoming aware of the adverse event. The principal investigator will make the final determination regarding each characteristic for the adverse event, including seriousness, expectedness, and relatedness.

For adverse events that meet the above criteria for recording and reporting, the coordinating center will notify the DSMB, the IRB, and the sponsor in accordance with the following reporting plan:

| **Characteristics of the Adverse Event** | **Reporting Period** |
| --- | --- |
| Fatal or life-threatening (and therefore serious), unexpected, and definitely related, probably or possibility related, or of uncertain relationship. | Report to DSMB and IRB within 7 days after notification of the event. |

| | |
|---|---|
| Serious but non-fatal and non-life-threatening, unexpected, and definitely related, probably or possibly related, or of uncertain relationship. | Report to DSMB and IRB within 7 days of notification of the event. |
| All other adverse events meeting criteria for recording and reporting. | Report to DSMB and IRB as part of continuing review. |

The investigator will distribute the written summary of the DSMB's periodic review of reported adverse events to the IRB in accordance with NIH guidelines (http://grants.nih.gov/grants/guide/notice-files/not99-107.html).

##### 9.4 Clinical Outcomes that may be Exempt from Adverse Event Recording and Reporting:

In this study of critically ill patients at high risk for death and other adverse outcomes due to their underlying critical illness, clinical outcomes, including death and organ dysfunction, will be systematically collected and analyzed for all patients. The primary and exploratory outcomes will be recorded and reported as clinical outcomes and not as adverse events unless treating clinicians or site investigators believe the event is Definitely Related or Probably or Possibly Related to the study intervention or study procedures. This approach – considering death and organ dysfunction as clinical outcomes rather than adverse events and systemically collecting these clinical outcomes for analysis – is common in ICU trials. This approach ensures comprehensive data on death and organ dysfunction for all patients, rather than relying on sporadic adverse event reporting to identify these important events. The following events are examples of study-specific clinical outcomes that would not be recorded and reported as adverse events unless treating clinicians or site investigators believe the event was Definitely Related or Probably or Possibly Related to the study intervention or study procedures:

- Death (all deaths occurring prior to hospital discharge or 28 days will be recorded);
- Organ dysfunction
  - Pulmonary – hypoxemia, hypercarbia, acute hypoxemic respiratory failure, pneumothorax;
  - Cardiac – hypotension, cardiac arrest or shock with or without receipt of vasopressors;
  - Delirium or coma;
- Duration of mechanical ventilation;
- Duration of ICU admission;
- Duration of hospitalization.

Note: A study-specific clinical outcome may also qualify as an adverse event meeting criteria for recording and reporting. For example, a pneumothorax that the investigator considers Definitely Related to ventilator mode would be both recorded as a study-specific clinical outcome and recorded and reported as a Serious and Definitely Related adverse event.

##### 9.5 Unanticipated Problems involving Risks to Subjects or Others:

Investigators must also report to the principal investigator Unanticipated Problems Involving Risks to Subjects or Others ("Unanticipated Problems"), regardless of severity, associated with study procedures

**within 24 hours** of the investigator becoming aware of the Unanticipated Problem. An Unanticipated Problem is defined as any incident, experience, or outcome that meets all of the following criteria:

- Unexpected (in terms of nature, severity, or frequency) given (a) the research procedures that are described in the protocol-related documents, such as the IRB-approved research protocol; and (b) the characteristics of the subject population being studied; AND
- Definitely Related or Probably or Possibly Related to participation in the research (as defined above in the section on characteristics of adverse events); AND
- Suggests that the research places subjects or others at a greater risk of harm (including physical, psychological, economic, or social harm) than was previously known or recognized.

Upon becoming aware of any event that may represent an Unanticipated Problem, the investigator will assess whether the event represents an Unanticipated Problem by applying the criteria described above. If the investigator determines that the event represents an Unanticipated Problem, the investigator will record the Unanticipated Problem in the Unanticipated Problem electronic case report form in the trial database. The investigators will obtain information about the Unanticipated Problem and report the Unanticipated Problem to the DSMB and the IRB within 15 days of becoming aware of the Unanticipated Problem.

#### 10.0 Statistical Considerations

*Sample size justification*.

The purpose of this trial is to assess feasibility of a cluster-randomized, cluster-crossover design. To assess whether compliance can be maintained after cross-over events, we will need to observe two cross-over events to each mode while maintaining an equal number of treatment blocks for each mode. Doing so will require a total of 9 months. Based on the experience of a prior trial in the same ICU,[70] we anticipate an average of 75 patients on mechanical ventilation will be enrolled per month. In 9 months, we expect to enroll 675 patients, of whom 69 will be enrolled during washout periods and excluded from the primary analysis. The resulting 606 patients will be included in the primary analysis.

While the sample size was determined to demonstrate feasibility of the cluster-crossover design, we anticipate this pilot will be the largest randomized trial of ventilator modes to date, and as such, we plan to compare ventilator free days to day 28 (primary outcome) between groups. In a prior cluster-randomized cluster-crossover trial in the same ICU,[70] mechanically ventilated patients experienced a median of 22 VFDs [IQR 0-25 VFDs] and an intra-cluster intra-period correlation of 0.01. With 606 (202 in each group) patients in the primary analysis, a standard deviation in the primary outcome of VFDs of 11 days, and a two-sided alpha of 0.05, the MODE trial will have 80 percent statistical power to detect an absolute difference between groups in the sole primary clinical endpoint of Ventilator Free Days of 3 days.

*Primary Analysis of the Primary Clinical Outcome*

The sole pre-specified primary outcome of ventilator free days will be compared between the three trial groups in an intention-to-treat fashion among all patients enrolled in the trial except those admitted during one of the 3-day washout periods. To estimate the conditional effect of a group assignment on an individual patient given the values of the covariates for that patient, we will use a proportional odds model with independent covariates of group assignment (volume control, pressure control, or adaptive pressure control) and time. Time (in days) will be treated as a continuous variable with values ranging from 1 (first day of enrollment) to 272 (final day of enrollment) and will be analyzed using restricted cubic splines with multiple knots to allow for non-linearity resulting from seasonality or secular trends. In addition to assessing for an overall group effect within the model, we will estimate the differences between each pair of modes by extracting 95% confidence intervals from the model.

*Sensitivity Analyses of the Primary Clinical Outcomes:*
We will repeat the primary clinical outcome analysis with adjustment for pre-specified baseline covariates.

We will repeat the analyses of the primary outcome and feasibility outcomes among all patients enrolled in the trial including those initiated on invasive mechanical ventilation in the study ICU during one of the pre-specified 3-day washout periods.

*Analysis of Effect Modification for the Primary Outcomes.*
We will examine whether pre-specified baseline variables modify the effect of study group on the primary clinical outcome using formal tests of statistical interaction in a proportional odds model. Independent variables will include study group assignment, the potential effect modifier of interest, and the interaction between the two (e.g., study group * history of COPD) and time. Significance will be determined by the P value for the interaction term, with values less than 0.10 considered to suggest a potential interaction and values less than 0.05 considered to confirm an interaction.

We will examine whether the following baseline variables modify the effect of study group on the primary outcome:

- Age (continuous variable);
- Duration of invasive mechanical ventilation prior to enrollment (0 minutes; 1 to 360 minutes; >360 minutes);
- Pre-enrollment fraction of inspired oxygen ($FiO_2$)(continuous variable);
- SOFA score at enrollment (continuous variable);
- Shock receiving vasopressors (yes, no)
- Indication for intubation (categories are not mutually exclusive)
    - Hypoxemic respiratory failure (yes, no);
    - Hypercarbic respiratory failure (yes, no);
    - Altered mental status or airway protection (yes, no);
- Chronic Obstructive Pulmonary Disease (yes, no);

*Analysis of the Exploratory, Feasibility, and Clinical Outcomes*
Each of the exploratory outcomes will be compared between groups in an intention-to-treat fashion in the primary analysis population. Continuous outcomes will be compared between groups in a similar manner as the primary clinical outcome. A logistic model will be used for binary outcomes, a multinomial model for categorical outcomes, and a proportional odds model for ordinal and continuous outcomes.

*Corrections for multiple testing*

We have pre-specified a single primary clinical outcome, which will have a test of statistical significance for difference between groups. Consistent with recommendations of the Food and Drug Administration and the European Medicines Association, the clinical outcome will be tested using a two-sided p-value with a significance level of 0.05. For all other analyses, emphasis will be placed on the estimate of effect size with 95% confidence intervals, as recommended by the International Committee of Medical Journal Editors, and no corrections for multiple comparisons will be performed.

*Handling of missing data*
The primary outcome is not anticipated to be missing for any patients. Missing data will not be imputed for the primary outcome or any secondary or exploratory outcomes. In adjusted analyses, missing data for covariates will be imputed using multiple imputations.

*Interim Analysis*
In addition to ongoing monitoring of safety throughout the trial, we will plan for the DSMB to conduct a single interim analysis for safety after the ICU has been assigned to each of the three study modes, that is, after enrollment of patients for 3 months. At the interim analysis, the DSMB will evaluate the incidence of the following outcomes across the study groups (volume control, pressure control, or adaptive pressure control):

- Pneumomediastinum or pneumothorax during the course of mechanical ventilation,
- Episodes of hypoxemia while receiving invasive mechanical ventilation: $SpO_2$ < 85% for more than 5 minutes,
- In-hospital mortality to study day 28.

The stopping boundary for safety will be met if the P value for the difference between groups is 0.016 or less, which is an a priori threshold determined by adjusting the p-value threshold of 0.05 for multiple comparisons by dividing 0.05 by the 3 outcomes. The DSMB will consider whether it is necessary to stop the trial or whether randomization to any one study group should be stopped, while the other two groups continue enrolling patients. Given the design of the study is to demonstrate feasibility and pilot trial procedures rather than demonstrating efficacy, there will be no stopping boundary for efficacy or futility.

#### 11.0 Privacy / Confidentiality Issues

At no time during this study, its analysis, or its publication, will patient identities be revealed in any manner. The minimum necessary data containing patient or provider identities will be collected. Data collected from the medical record and the Mode Modification Sheets will be entered into the secure online database REDCap. Only key study personnel will have access to the REDCap database. Hard copies of the treating clinician Mode Modification Sheet will be stored in a locked room until after the completion of enrollment and data cleaning.

All data will be maintained in the secure online database REDCap until the time of study publication. At the time of publication, a de-identified version of the database will be generated.

#### 12.0 Follow-up and Record Retention

Patients will be followed after enrollment for 28 days or until hospital discharge, whichever occurs first. Data collected from the medical record will be entered into the secure online database REDCap. Hard copies of the treating clinician Mode Modification Sheet will be used for manual data entry into the REDCap database and stored in a locked room until after the completion of enrollment and data cleaning. All data will be maintained in the secure online database REDCap until the time of study publication. Once data are verified and the database is locked, all hard copies of Mode Modification Sheets will be destroyed. At the time of publication, a de-identified version of the database will be generated.

#### 13.0 Data Safety and Monitoring Board (DSMB)

The principal role of the DSMB is to assure the safety of patients in the trial. They will regularly monitor data from this trial, review and assess the performance of its operations, and make recommendations to the steering committee and sponsor with respect to:

- Participant safety and risk/benefit ratio of study procedures and interventions
- Protocol amendments (with specific attention to study population, intervention, and study procedures)
- Adherence to the protocol requirements
- Completeness, quality, and planned analysis of data
- Ancillary study burden on participants and main study
- Possible early termination of the trial because of new external information, safety concerns, or inadequate performance.

The DSMB will consist of members with expertise in critical care medicine, pulmonary medicine, biostatistics, and clinical trials. Appointment of all members is contingent upon the absence of any conflicts of interest. All the members of the DSMB are voting members. The Principal Investigator and unblinded study biostatistician will be responsible for the preparation of all DSMB and adverse event reports. The DSMB will develop a charter and review the protocol and patient notification forms during its first meeting. Subsequent DSMB meetings will be scheduled in accordance with the DSMB Charter with the assistance of the Principal Investigator. The DSMB will have the ability to recommend that the trial end, be modified, or continued unchanged.

**Appendix 1: Ventilator Guidelines**

SUMMARY OF VUMC MICU VENTILATOR PROTOCOL GUIDELINES:

**Ventilator Set Up and Adjustment:**

1. Calculate Predicted Body Weight (PBW):
    a. In males = 50 - 2.3 (height (inches) - 60)
    b. In females = 45.5 + 2.3 (height (inches) - 60)

2. Select Mode and initial default settings:
    a. **Volume Control.** Set Tidal Volume (Vt) to target 6 mL/kg PBW. Set Flow Pattern: 100 or [square-wave icon]; Set T pause (s): 0.
    b. **Pressure Regulated Volume Control.** Set Vt to target 6 mL/kg PBW.
    c. **Pressure Control.** Set PC above PEEP to 15 $cmH_2O$, set inspiratory time to 0.9 s and confirm delivered Vt is appropriate for target 6 mL/kg PBW.

3. Set initial respiratory rate (RR) to approximate baseline Minute Ventilation (12-16 bpm, not > 35 bpm).

4. Set alarms:
    For **VC and PRVC**: High Peak Pressure alarm at 40 cm $H_20$
    For **PC**: Low Minute Ventilation alarm at 2 L/min less than the current Minute Ventilation

5. Adjust Vt and RR to achieve pH and plateau pressure goals below.

**Lung Protective Ventilation Goals – Vt** ≤ 6 mL/kg PBW**, Pplat** < 30 $cmH_20$

**For Pressure Control, where Vt cannot be directly adjusted, adjust PC over PEEP in increments of 2 $cmH_20$ and monitor resulting Vt to achieve the changes outlined below.*

If Vt > 6 mL/kg PBW:
    Reduce Vt by 1 mL/kg at intervals ≤ 2 hours until Vt = 6 mL/kg PBW.
If Pplat > 30 (or PIP > 35):
    Decrease Vt by 1 mL/kg steps to a minimum of 4mL/kg PBW.
If Pplat < 25 and Vt < 6 mL/kg:
    Increase Vt by 1 mL/kg until either Pplat > 25 or Vt = 6 mL/kg PBW.
If Pplat <3 0 and breath stacking occurs:
    May increase Vt in 1 mL/kg increments to maximum of 8 mL/kg PBW.

**Ventilation Goals - pH**: 7.30-7.45

If pH 7.15-7.30:
    Increase RR until pH > 7.30 or PaCO2 < 25 (Max RR = 35).
If RR = 35 and PaCO2 < 25:
    May give $NaHCO_3$, to be managed by primary team.
If pH < 7.15:
    Increase RR to 35.
If pH remains < 7 .15 and $NaHCO_3$ considered or infused:

Vt may be increased until pH >7.15 or Vt = 8 mL/kg PBW (Pplat target may be exceeded).

If pH >7.45:
Decrease RR rate.

**Oxygenation goals - $SpO_2$:** 88-95% or **$PaO_2$:** 55-80 mmHg.

Use incremental $FiO_2$/PEEP combinations below to provide minimum $FiO_2$ and PEEP necessary to achieve $PaO_2$ or $SpO_2$ goals.

| **FiO2** | 0.3-0.4 | 0.4 | 0.5 | 0.5 | 0.6 | 0.7 | 0.7 | 0.7 | 0.8 | 0.9 | 0.9 | 0.9 | 1 |
|---|---|---|---|---|---|---|---|---|---|---|---|---|---|
| **PEEP** | 5 | 8 | 8 | 10 | 10 | 10 | 12 | 14 | 14 | 14 | 16 | 18 | 18-25 |

**Other Settings**

**I:E ratio goal:** 1:1.0 – 1-3.0. Adjust Ti to achieve goal.
If $FiO_2$ = 1.0 and PEEP = 24 $cmH_20$, may adjust I:E to 1:1.
Avoid inverse-ratio ventilation.
**Trigger:** 1.6 L/min by default, can be increased or reduced to improve synchrony
**Tinsp rise (s):** 0.15 s by default, can be increased or decreased to improve ventilation

**Monitoring and Documentation:**

Measure & record Pplat with inspiratory pause of at least 0.5 s, $SpO_2$, Total RR, Vt and pH (if available) at least every 4 hours AND after each change in PEEP, Vt, or PC above PEEP settings.

Once the patient tolerates $FiO_2$ < 0.5 and PEEP < 8 $cmH_2O$, advance to Part II: Spontaneous Breathing Trial Readiness Assessment and Ventilator Discontinuation Guidelines.

**Appendix 2. Ventilator Guidelines Table**

| | Volume Control | Pressure Control | Adaptive Pressure Control |
|---|---|---|---|
| Mode option | Volume Control | Pressure Control | Pressure Regulated Volume Control (PRVC) |
| Tidal Volume target (mL/kg PBW) | 6 | 6 | 6 |
| Tidal Volume range (mL/kg PBW) | 4 - 8 | 4 - 8 | 4 - 8 |
| Initial breath settings | Tidal volume to 6 mL/kg PBW | PC over PEEP to 15 cm H20, Insp Time to 0.9 sec | Tidal volume to 6 mL/kg PBW |
| Achieving target Tidal Volume | Directly adjust tidal volume | Dependent on adjustment of PC over PEEP | Adjust tidal volume directly |
| Plat Pressure target (cm H20) | ≤ 30 | ≤ 30 | ≤ 30 |
| Achieving target Plateau Pressure | Dependent on adjustment of tidal volume | Directly adjust PC over PEEP | Dependent on adjustment of tidal volume |
| Flow | Set directly | Dependent ventilator and patient factors | Dependent ventilator and patient factors |
| Flow Pattern | Square waveform | Decelerating, dynamic | Decelerating, dynamic |
| Arterial pH goal | 7.3 - 7.45 | 7.3 - 7.45 | 7.3 - 7.45 |
| Ventilator rate (breaths/min) | ≤ 35 | ≤ 35 | ≤ 35 |
| Oxygenation goal by PaO2 (mmHg) | 55 - 80 | 55 - 80 | 55 - 80 |
| Oxygenation goal by SpO2 (%) | 88 - 95 | 88 - 95 | 88 - 95 |
| Positive end-expiratory pressure | Set with FiO2-PEEP table | Set with FiO2-PEEP table | Set with FiO2-PEEP table |
| Inspiratory: Expiratory ratio | 1:1 - 1:3 | 1:1 - 1:3 | 1:1 - 1:3 |
| Extubation evaluation | MICU SBT Readiness and Extubation Guidelines | MICU SBT Readiness and Extubation Guidelines | MICU SBT Readiness and Extubation Guidelines |

**Appendix 3: Spontaneous Breathing Trials and Ventilator Discontinuation Guidelines**

**Part I – Spontaneous Breathing Trial Readiness Assessment**

Every morning or at least once daily, patients will undergo screening and trial in a Spontaneous Awakening Trial (SAT) performed by bedside nursing. If the patient passes the SAT, then the patient should be promptly evaluated in a Spontaneous Breathing Trial (SBT) Readiness Assessment as a safety screening:

If all of the following criteria have been present for more than 2 hours, then evaluate the patient for progression to Spontaneous Breathing Trial:
a) $FiO_2 \leq 0.5$ & PEEP $\leq 8$ $cmH_2O$
b) Values of both PEEP and $FiO_2 \leq$ values from previous 12 hours.
c) Not receiving neuromuscular blocking agents
d) Patient is exhibiting adequate inspiratory efforts (if not, decrease the ventilator rate to 50% of baseline level for up to 5 minutes to detect inspiratory effort).
e) Systolic arterial pressure > 90 mmHg without significant vasopressor support (< 10 mcg/min of norepinephrine or < 5 mcg/kg/min dopamine or dobutamine will not be considered a vasopressor).

**Part II – Spontaneous Breathing Trial (SBT)**

If criteria a-e above are met, then initiate a trial of at least 30 minutes of spontaneous breathing with FIO2 ≤ 0.5 using any of the following approaches:
1. Pressure support ≤ $5cmH_2O$, PEEP ≤ $5cmH_2O$
2. CPAP ≤ 5 $cmH_2O$
3. T-piece
4. Tracheostomy mask

**Monitor for tolerance using the following:**
1. $SpO_2 \geq 88\%$ and / or $PaO_2 \geq 60$ mmHg. $FiO_2$ may be increased to 0.5 if necessary to maintain oxygenation in the target range.
2. Mean spontaneous tidal volume ≥ 4 mL/kg PBW (if measured)
3. Total Respiratory Rate ≤ 35 / min (< 5 min at RR > 35 may be tolerated, example post suctioning)
4. pH ≥ 7.30 (if measured)
5. No respiratory distress (defined as 2 or more of the following):
    a. Heart rate ≥ 120% of the 06:00 rate (≤ 5 min at > 120% may be tolerated)
    b. Marked use of accessory muscles
    c. Abdominal paradoxical breathing
    d. Diaphoresis
    e. Marked subjective dyspnea.

If any of the goals 1-5 are NOT met, revert to previous ventilator settings with Positive End-expiratory Pressure and $FiO_2$ = previous settings and reassess for SBT the next morning.

The clinical team may decide to change mode of support during spontaneous breathing (PS = 5cm$H_2O$, CPAP, tracheostomy mask, or T-piece) at any time.

**Part III – Decision to Remove Ventilatory Support**

For intubated patients, if tolerance criteria for spontaneous breathing trial (1-5 above) are met for at least 30 minutes, continue with unassisted breathing and extubate if clinically indicated under the direction of team physicians. However, the spontaneous breathing trial can continue for up to 120 minutes if tolerance remains in question (in fragile patients).

If any of criteria 1-5 are NOT met during spontaneous breathing (or 120 minutes has passed without clear tolerance), then the ventilator settings that were in use before the attempt to wean will be restored (Positive End-expiratory Pressure and $FiO_2$ = previous settings) and the patient will be reassessed for an SBT the following day.

**Tracking of Protocol Versions:**

Version 1.0 – Initial Protocol (IRB approval on 10/18/22)

Version 1.1 – Protocol revisions after initial meeting with DSMB (10/21/22, IRB approval on 12/6/22)

- Adjusted for multiple testing at the interim analysis by dividing p-value threshold used to stop for safety outcomes being formally evaluated, as follows:
  *"The stopping boundary for safety will be met if the P value for the difference between groups is 0.016 or less, which is an a priori threshold determined by adjusting the p-value threshold of 0.05 for multiple comparisons by dividing 0.05 by the 3 outcomes."*

- Clarified that the DSMB is empowered, in addition to making recommendations regarding stopping the trial, to make a recommendation to drop one arm of the three-armed trial if one arm appears significantly worse than the remaining two arms. The following has been added:
  *"The DSMB will consider whether it is necessary to stop the trial or whether randomization to any one study group should be stopped, while the other two groups continue enrolling patients."*

- Clarified what mode of ventilation is recommended for patients who are extubated and reintubated during the study block by adding:
  *"If a patient is enrolled, extubated, and re-intubated during the same study block, the study protocol will again determine the ventilator mode until they meet one of the aforementioned criteria."*

Version 1.2 – Minor protocol revisions to align with Statistical Analysis Plan (7/21/23, IRB approval pending):

- Clarified definitions of exploratory outcome variables
- Clarified definition of primary outcome
- Modified choices of variables to be analyzed as potential effect modifiers
- Modified definition of the variable for Acute Respiratory Distress Syndrome
- Removed the plan for additional statistical approaches to the analysis of the primary outcome

**Mode of Ventilation in Critical Illness (MODE) trial**

Summary of Changes to Study Protocol

| | |
|---|---|
| **9/26/2022** | **Original Protocol**, version 1.0 |
| **10/21/2022** | Amendment to Study Protocol, version 1.1<br><br>At the request of the DSMB, the protocol was updated with minor changes to (1) the safety stopping criteria for the planned interim analysis, (2) clarify that in addition to having the power to recommend stopping the trial at the interim analysis, the DSMB had the power to recommend stopping one arm and allowing the other two arms to continue, and (3) clarifying how the mode of mechanical ventilation would be managed for patients who are enrolled, extubated, and reintubated during the same study block. |
| **11/1/2022** | First patient enrolled |
| **7/21/2023** | Amendment to Study Protocol, version 1.2 **(Final Protocol)**<br><br>Minor changes were made to update protocol to match published statistical analysis plan by: (1) refining outcome and on-study variable definitions, (2) updating prespecified effect modifiers, (3) finalizing plans for secondary/sensitivity analyses. |
| **7/31/2023** | Enrollment complete |

**Mode of Ventilation in Critical Illness (MODE) trial**

Summary of changes to the Statistical Analysis Plan

| | |
|---|---|
| **7/24/2023** | **Original Statistical Analysis Plan** complete and posted to m*edRxiv* server and submitted for review at *CHEST Critical Care*<br><br>Seitz KP, Lloyd BD, Wang L, et al. Protocol and Statistical Analysis Plan for the Mode of Ventilation During Critical Illness (MODE) Trial. *medRxiv.* 2023.07.21.23292998; doi: https://doi.org/10.1101/2023.07.21.23292998 |
| **7/31/2023** | Enrollment complete |
| **11/25/2023** | Statistical Analysis Plan, initially submitted on Jul 21, 2023 is published online by *CHEST Critical Care*<br><br>Changes made in response to peer review were limited to additional description/rationale for: the chosen ventilator modes, the exclusion criteria, the management of patients who continued to receive mechanical ventilation following a cluster cross-over, and the management following a mode modification sheet.<br><br>No changes were made in peer review to the trial procedures, outcomes, or statistical analysis plan. |
| **2/22/2024** | **Final Statistical Analysis Plan version of record published** by *CHEST Critical Care*<br><br>No changes were made from the *CHEST Critical Care* online version.<br><br>Seitz KP, Lloyd BD, Wang L, et al. Protocol and Statistical Analysis Plan for the Mode of Ventilation During Critical Illness (MODE) Trial. *CHEST Critical Care*. Volume 2, Issue 1, March 2024, 100033, doi: 10.1016/j.chstcc.2023.100033 |
